# Safety, effectiveness, and skin immune response in a controlled human infection model of sand fly transmitted cutaneous leishmaniasis

**DOI:** 10.1101/2024.04.12.24305492

**Authors:** Vivak Parkash, Helen Ashwin, Shoumit Dey, Jovana Sadlova, Barbora Vojtkova, Katrien Van Bocxlaer, Rebecca Wiggins, David Thompson, Nidhi Sharma Dey, Charles L. Jaffe, Eli Schwartz, Petr Volf, Charles J. N. Lacey, Alison M. Layton, Paul M. Kaye

## Abstract

The leishmaniases are globally important parasitic diseases for which no human vaccines are currently available. To facilitate vaccine development, we conducted an open label observational study to establish a controlled human infection model of sand fly-transmitted cutaneous leishmaniasis caused by *L. major*. Between 24^th^ January and 12^th^ August 2022, we exposed 14 (8F, 6M) participants to infected *Phlebotomus duboscqi*. The primary objective was to demonstrate effectiveness (take rate) and safety (absence of CL lesion at 12 months), whereas secondary and exploratory objectives included rate of lesion development, parasite load and analysis of local immune responses by immunohistology and spatial transcriptomics. We estimated an overall take rate for CL development of 64% (9/14), or 82% (9/11) if calculated using only participants having confirmed bites following exposure. Lesion development was terminated by therapeutic biopsy in 10 participants with confirmed bites. 2/10 had one and 1/10 had two lesion recurrences 4-8 months after biopsy that were treated successfully with cryotherapy. No severe or serious adverse events were recorded, but scarring was evident as expected. All participants were lesion-free at >12 month follow up. We provide the first comprehensive map of immune cell distribution and cytokine/chemokine expression in human CL lesions, revealing discrete immune niches. This controlled human infection model offers opportunities for rapid vaccine candidate selection and a greater understanding of immune-mediated protection and pathology.

## Introduction

The leishmaniases are vector-borne diseases transmitted by phlebotomine sand flies ^1^ and have a global impact on health and well-being ^2^. Several species of *Leishmania* infect humans, causing a diverse spectrum of tegumentary and systemic diseases ^1^. Cutaneous leishmaniasis (CL) is endemic in 89 countries reporting to the WHO and over 200,000 new autochthonous cases were reported in 2020, though this is widely regarded as a significant under-estimate of true disease burden ^3^. CL presents as an inflammatory lesion at the site of transmission, but clinical outcome is typically dependent on parasite species. Lesions due to *L. major* commonly develop and then self-resolve over a period of months and / or respond well to topical therapy. The resulting scar may, however, have lifelong impact on well-being ^4–6^. Lesions due to *L. tropica* in the Old World and *L. mexicana* in the New World are more chronic, persist often for years and can be relatively refractory to treatment. Some species have metastatic potential (e.g. *L. braziliensis*, *L. guyanensis*) leading to mucocutaneous leishmaniasis, whereas others may spread within the skin causing disseminated or diffuse leishmaniasis (e.g. *L. aethiopica*). Two species, *L. donovani* and *L. infantum*, typically disseminate from the primary inoculation site to cause the systemic and life-threatening disease visceral leishmaniasis (VL), endemic in 79 countries. 12,739 cases of VL were reported to WHO in 2020, a significant decline on past estimates and reflective of the impact of a regional elimination campaign in South Asia. VL case fatality rates (2-3%) have, however, changed little over the past decade and poorer outcomes are reported in some countries (e.g. Brazil at 7.1%) ^3^. Since vector control alone is likely to be insufficient, controlling the leishmaniases by vaccination is an attractive option.

The potential for a vaccine to reduce the public health burden has been well-recognised ^7,8^. A recent report by the WHO Product Development for Vaccines Advisory Committee (PDVAC) ranked *Leishmania* as the highest priority parasitic target for new vaccine development after *Plasmodium falciparum* malaria ^9^. Recent studies have also provided estimates of the global demand for ^10^ and affordability of ^11^ a successful leishmaniasis vaccine. However, despite several decades of effort and numerous studies in animal models ^12,13^, few *Leishmania* vaccine candidates have progressed to clinical trial ^8^ and only two are currently in clinical development, an adenovirus-vectored vaccine encoding two *Leishmania* antigens (ChAd63-KH ^14^) and a live genetically attenuated vaccine ( *L. major* cen^-/-^ ^15^). Whilst multiple factors adversely impact the vaccine pipeline ^12^, new approaches are needed to identify and validate candidate vaccines, improve understanding of natural- and vaccine-induced protective immunity in humans, and ultimately shorten the pathway to registration. Experimental human infection, so-called challenge studies or controlled human infection models (CHIMs) can address these issues directly.

CHIM studies are now well-embedded in the vaccine development pathway for numerous infectious diseases including the parasitic diseases malaria ^19,20^, schistosomiasis ^21^ and hookworm ^22^. Deliberate human infection with *Leishmania* is not new ^23^ and “leishmanization”, the inoculation of lesion scapings into cosmetically hidden areas, has been practiced for centuries in CL endemic countries to induce immunity and minimise visible scarring and stigma ^24^. Leishmanization was proposed for vaccine development in the early 2000’s providing evidence of feasibility ^25^, but not pursued. Subsequently, the importance of vector-associated modulation of immunity has become more fully appreciated ^26^ and evidence suggests that vaccines effective against needle challenge may not protect against natural transmission ^27^.

Here, we provide the first report detailing a CL CHIM that incorporates natural sand fly transmission. The study was designed with the primary objectives of determining take rate (defined as lesion development) after exposure to infected sand flies and safety (absence of lesions at 12 month follow up). We report that this model is safe and well-tolerated by study participants. In addition, analysis of lesion biopsies provided new insights into the immune landscape associated with early CL development in humans.

## Results

### Study design

We previously reported on prior enabling studies, including development of a cGMP challenge strain of *L. major* (MHOM/IL/2019/MRC-02)^28^, development of a sand fly biting protocol ^29^ and focus group assessments of public perceptions of the project ^30^. For this first study involving human challenge (LEISH_Challenge; ClinicalTrials.gov *Identifier: NCT04512742*), we enrolled 14 healthy *Leishmania*-naïve volunteers aged 18-50 at the University of York Translational Research Facility (**Fig. 1 and Extended Data 1 and 2)**.

**Figure 1.**
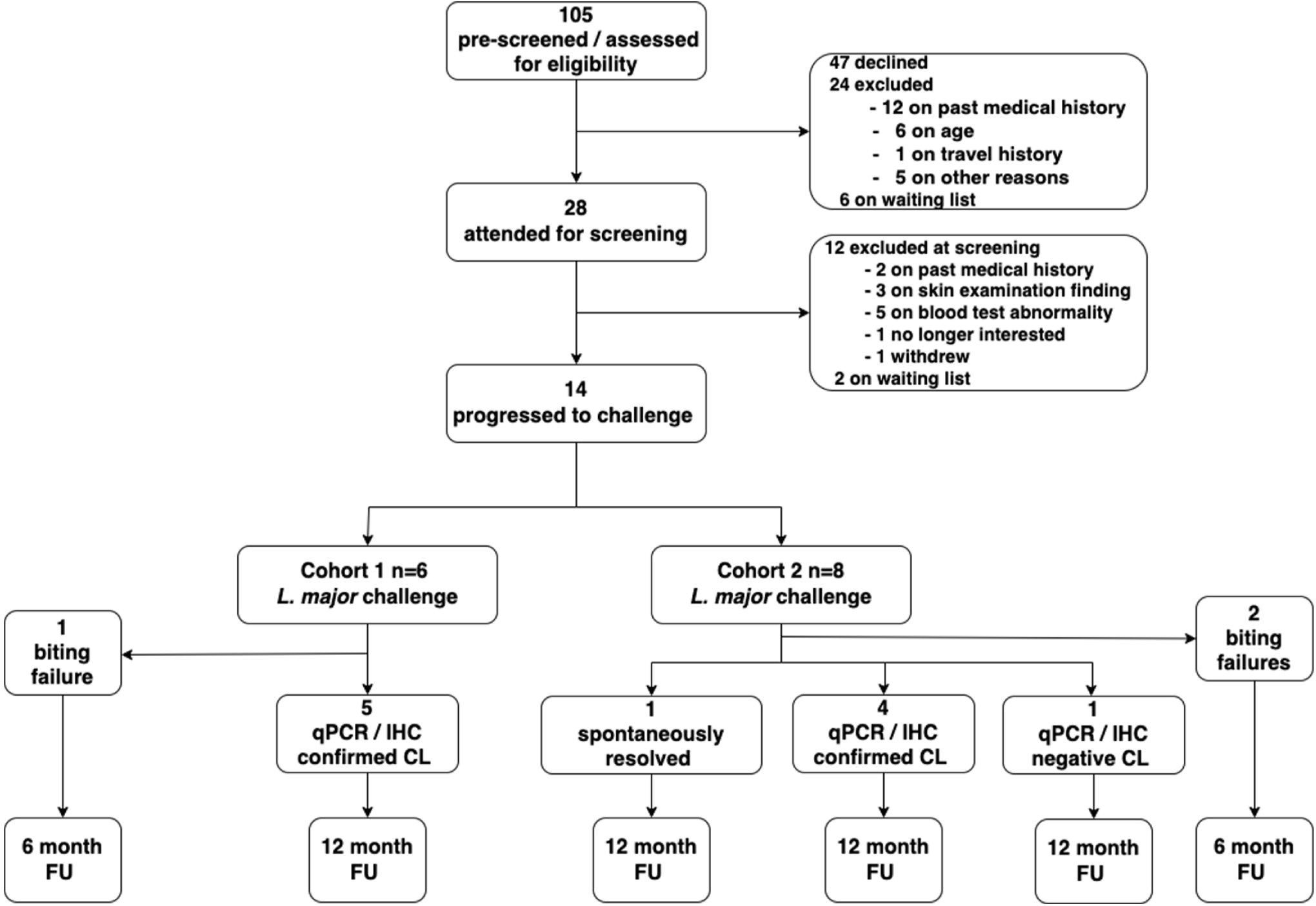
CONSORT diagram summarising the LEISH_Challenge study. A CONSORT checklist in included as **Extended Data 1**, details of participant demographics as **Extended Data 2** and study protocol and participant information sheet as **Extended Data 3**. CL, cutaneous leishmaniasis lesion; FU, follow up; IHC, immunohistochemistry.

First and last participants were exposed to infected sand flies between January and August 2022 respectively. There were 8 female and 6 male participants, median 32 years old, all white ethnicity reflecting our local resident population (98% white ethnicity). Sex was not explicitly factored into the experimental design. Sex was assigned and gender was not recorded. Exclusion and inclusion criteria were as per study protocol **(Extended Data 3**) and included absence of *Leishmania* exposure history, willingness to refrain from travel to *Leishmania major*-endemic regions during the study and absence of significant atopy or active skin disease. All participants had negative HIV, hepatitis B and hepatitis C serology and gave written informed consent. A pragmatic adaptive design was chosen to expose the least number of participants to infection and provide flexibility based on initial outcomes.

Five *Phlebotomus duboscqi* sand flies infected with *L. major* MHOM/IL/2019/MRC-02 were allowed access for 30 min to the volar aspect of the proximal forearm, approximately 2-3 centimetres distal to the antecubital fossa, using a bespoke biting chamber with a variable aperture ^29^. Participants were considered for therapeutic biopsy when a clinically apparent lesion of ≥3mm diameter was observed. However, biopsy could not always be performed on the day of evaluation and was often postponed until another study visit could be scheduled.

### Development of CL after sand fly exposure

For the first 6 participants, we used a biting chamber aperture of 6mm diameter. 1/6 participants (17%; LC012) was deemed a bite failure (see Methods) but developed a small 2mm diameter papule at the exposure site 4 weeks later. This persisted for 3 weeks and was removed by punch biopsy but showed no evidence of CL or clinically significant histological abnormality. The remaining 5/6 participants had 1-6 confirmed bites (**Fig. 2a-b**) received from between 1-3 sand flies (**Fig. 2c**) and all developed a clinically compatible CL lesion (median area at day 13-16 of 18.9 mm^2^; range 12.6 mm^2^ – 23.6 mm^2^; **Fig 2d and e**).

**Figure 2.**
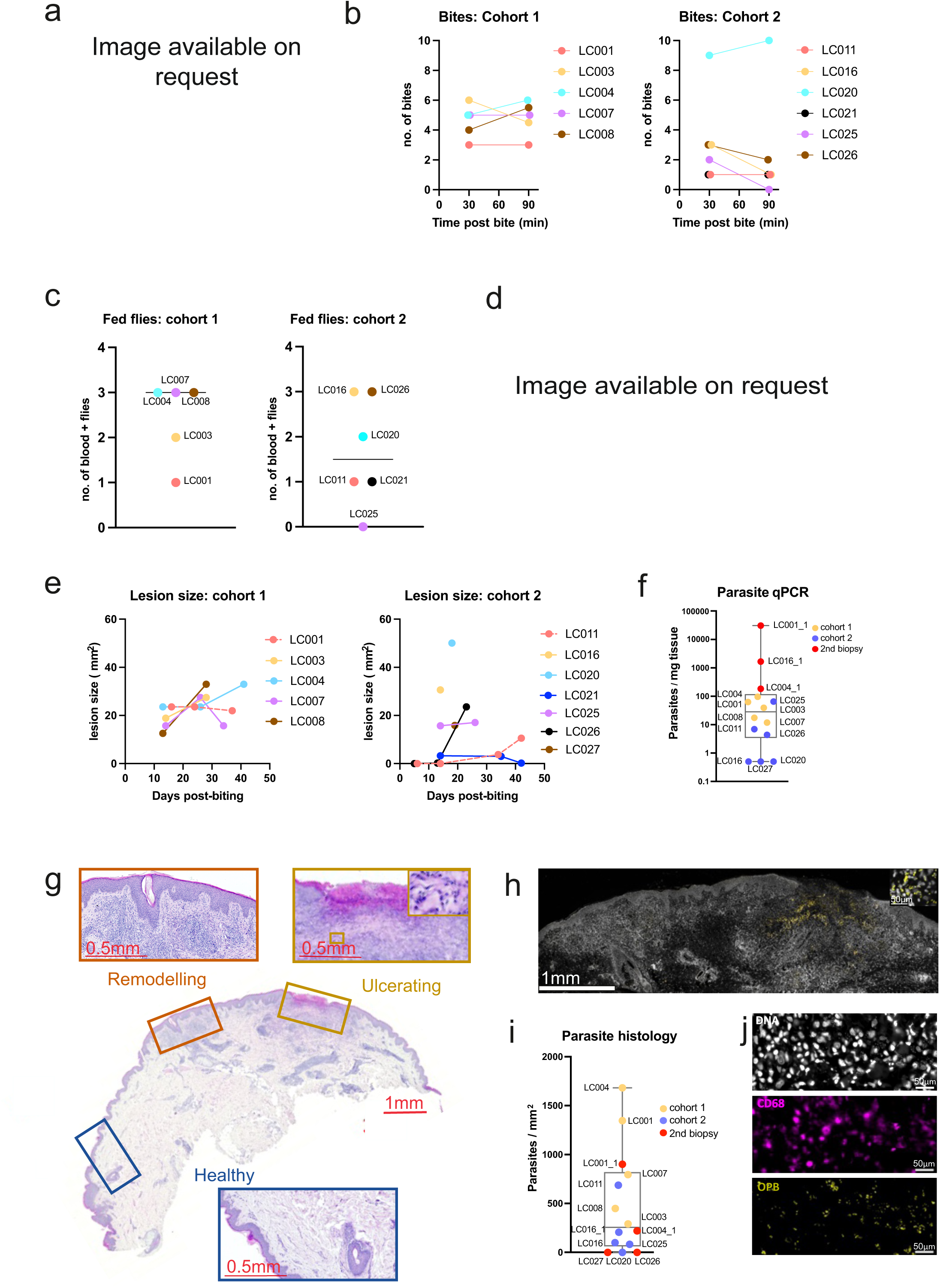
Parasitological outcomes in the LEISH_Challenge study. **a,** Dermoscopy image showing bite sites; **b,** number of recorded bites per participant per cohort at 30 and 90 minutes; No significant differences were noted between cohorts at either 30min (p=0.104) or 90 min (p=0.08; Mann-Whitney test). **c,** Number of partially and/or fully blood fed sand flies post biting per participant per cohort (p=0.36; Mann Whitney test) Bar represents median; **d,** CL lesion development and associated dermoscopy image (LC004, 13 days p.b.). **e,** Lesion areas (mm^2^) at varying times post bite per participant per cohort; **f,** Parasite load per mg biopsy tissue. Individual symbols reflect a single participant; box and whisker plot with median and max / min values. **g,** H&E-stained biopsy tissue from LC001, highlighting histologically normal tissue, a potential bite site with epidermal remodelling and an ulcer; Box in upper right image indicates higher magnification view of area of parasitism. H&E-stained sections from all other participants are shown in **Extended Data 4.** Scale bar = 1mm. **h,** IHC for *Leishmania* Oligopeptidase B (OPB, yellow) counterstained for nuclei (YOYO-1; white). Area shown represents remodelling and ulcerated regions shown in (g). Box shows higher magnification of area of parasitism. **i,** Parasites / mm2 of tissue determined by quantitative morphometry. Symbols show each participant and respective cohort; box and whisker plot with median and max / min values. **j,** IHC for OPB (yellow) and CD68 (purple) to show intracellular parasitism. Parasites are also evident by nuclear staining (YOYO1; white) with characteristic nucleus / kinetoplast. Scale bar = 20μm. Images are available on request as per medRxiv requirements.

Dermoscopy supported a diagnosis of leishmaniasis, showing characteristic erythema, teardrop-like structures, hyperkeratosis and vascular structures which included linear, dotted and hairpin-like vessels ^31^. Therapeutic excision biopsy was performed between day 28 and 41 post bite (median 34 days) allowing for confirmation of CL by qPCR and / or immunohistochemistry. 5/5 biopsies were positive by qPCR, albeit with high degree of variance in parasite load (median 1218 parasites/mg tissue, range 255/mg - 27547/mg; **Fig.2f**). Dermal cell infiltration of varying intensity was evident in all cases and immunostaining for *Leishmania* Oligopeptidase B (OPB) along with characteristic DAPI staining confirmed parasites in all 5 volunteers (**Fig. 2g-j** and **Extended Data 4**). In some sections, discrete foci of cellular infiltration were observed consistent with multiple bites. Responses at these different sites appeared heterogeneous. For example, in LC001 we observed one area of focal infiltration accompanied by epidermal remodelling and with scant parasites adjacent to an ulcer with extensive underlying parasitism (**Fig 2g and h**). By our per protocol definition (see Methods), we calculated the take rate for this cohort as 83% [95% CI, 0.44, 0.97] rising to 100% [95% CI, 0.57, 1] for participants with at least one confirmed bite.

For the second cohort (n=8) we made two procedural changes intended to minimise lesion size and / or subsequent scarring. We explored reducing the biting aperture to 3 mm (LC016), 4 mm (LC011, LC021, LC023, LC026) or 5 mm (LC020, LC025, LC027). Although this had no significant effect on the number of bites received (**Fig. 2b**) or number of fed flies (**Fig. 2c**), there was a trend towards a reduction in both indices of transmission compared to cohort 1. We also performed lesion biopsy earlier (median 18 days) and used 6-8 mm punch biopsy rather than excision biopsy. 2/8 volunteers (25%; LC023, LC027) were deemed bite failures. LC027 developed a minor localised reaction near the site of sand fly exposure and was biopsied at day 19 for further investigation. Parasite qPCR and IHC were negative, no histological abnormalities were observed, and the participant remained lesion free (**Extended Data 4**). Of the remaining 6 volunteers with confirmed bites, LC021 had a small palpable lesion (maximum lesion diameter of 3.3 mm^2^) that spontaneously resolved by d42, and no biopsy or qPCR was performed. LC011, LC016, LC020, LC025 and LC026 developed a clinically compatible lesion (median area at day 14 – 19 study visits of 9.5 mm^2^, range 0 mm^2^ -50.1 mm^2^; **Fig. 2e**). Early lesion areas were more variable than in cohort 1 but not significantly different (p=0.82, Mann-Whitney) and parasite load determined by qPCR or IHC was again highly variable (**Fig. 2f and i**). LC020 was negative by qPCR and IHC, and histology lacked focal dermal infiltration and therefore did not meet our per protocol lesion definition, despite being clinically compatible. For LC026, qPCR was positive with a pronounced dermal infiltration, albeit parasites were not observed by IHC. LC016 was qPCR negative but IHC positive with typical CL histology. We estimated take rate for cohort 2 as 50% [95% CI, 0.22, 0.78], or 67% [95% CI, 0.3, 0.90] for those with confirmed bite. Across both cohorts in this study, we determined a take rate of 64% [95% CI, 0.39, 0.84] for all participants (9/14) or 82% [95% CI, 0.52, 0.95] for those with confirmed bite (9/11).

### Recurrence following therapeutic biopsy

Of the 10 participants having therapeutic biopsy, 7 (70%) required no further treatment and remain lesion free (**Fig. 3a-h**). In three cases (LC001, LC004, LC016), a lesion subsequently developed at the biopsy site 4-8 months after biopsy (**Fig. 3i-l**). A second biopsy (punch) was performed in each participant for parasitological confirmation (30800, 184 and 1658 parasites /mg tissue in biopsies taken at 255 days, 282 days, and 126 days post bite, respectively). Cryotherapy was initiated using a standard cryotherapy delivery device (0.75mm nozzle and repeated 10 second freeze/thaw cycle) as per protocol with all three volunteers receiving 3 treatments spaced over 6-8 weeks. All responded well with apparent healing of their CL lesion. LC004 had a second recurrence 14 months post original biopsy. This later lesion resolved with an additional cycle of cryotherapy.

**Figure 3.**
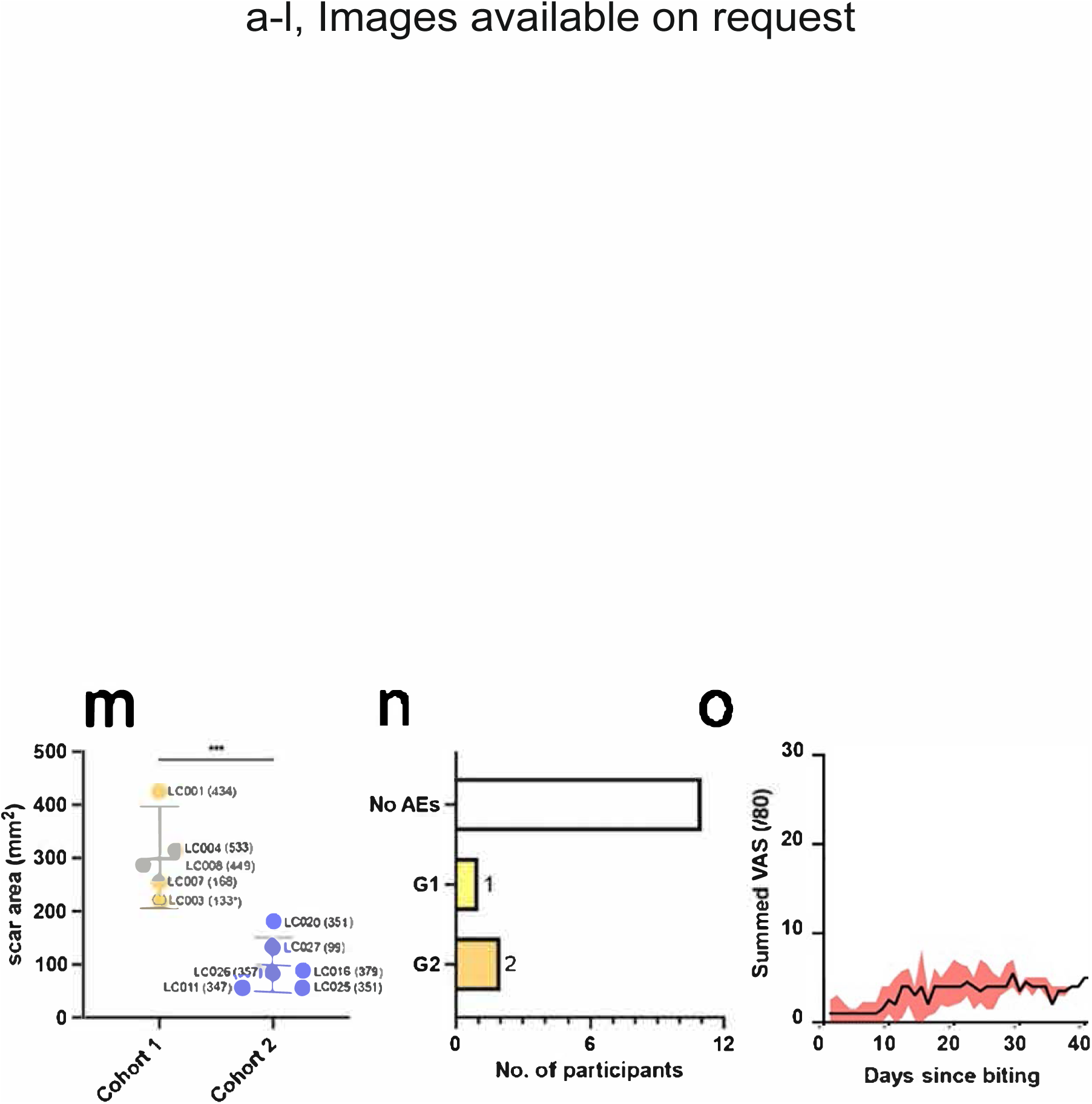
Clinical features of LEISH_Challenge. **a-d,** Participant LC008 images taken 28 days post bite (p.b.) and prior to 6mm excision biopsy (a, b) and 223 days (c) and 476 days (d) p.b. **e-h,** Participant LC011 images taken 42 days p.b. and prior to 4 mm punch biopsy (a, b) and at 140 days (c) and 350 days (d) p.b. Dermoscopy images are shown in b and f. **i-l,** Participant LC001 had a primary lesion (excised 37 days p.b;) followed by a secondary lesion (4mm punch biopsy and cryotherapy at 255 days p.b.). Images showing secondary lesion adjacent to healing primary lesion at 161 days (a) 251 days (b), 309 days (c) and 470 days (d) p.b. **m,** Quantitation of scar area at final measured follow up. Data points are labelled by participant ID and time in days from initial biopsy. ***, p<0.001 (Student’s t test). Further details are in **Extended Data 2**. **n,** Number of adverse events by grade assigned as definitely, probably, or possibly related to the study. Further details are in **Extended Data 5**. **o,** Summed participant-recorded visual analogue score across 8 parameters (itch, pain, erythema, swelling, malaise, myalgia, fever, nausea). Maximum score available = 80. Data are shown as median and IQR for all participants. Individual participant scores and summed scores for each parameter are provided in **Extended Data 7**. Images are available on request as per medRxiv requirements.

We considered potential factors influencing recurrence. For LC001, further evaluation of dermoscopy images suggested that one bite site was located beyond the perimeter of the original biopsy and may have been a slow to evolve primary lesion rather than a recurrence (**Fig. 3i-l)**. Participant LC004 confirmed that trauma had occurred at the biopsy site prior to the development of the second CL lesion. This participant remained concerned about the residual hypertrophic scarring post cryotherapy and Adcortyl (0.25 ml of 10mg/ml) was administered to the scar tissue. A new lesion developed 2 months later questioning whether the intralesional steroid was a contributory factor. For LC016, there were no precipitating factors identified. All these participants remain lesion free at the time of writing.

### Scarring in CHIM participants

Scarring was evident in all subjects who had a biopsy. Seven of the participants (LC007, LC008, LC011, LC016, LC020, LC025, LC026) were left with mild atrophic scarring in keeping with the excision biopsy plus the effect of CL. Two participants who developed atypical lesions where punch biopsy did not confirm CL (LC021, LC027) also developed mild atrophic scarring. One participant (LC003) had moderate scarring. Two participants who developed a second CL lesion (LC001, LC004) had more moderate to severe scarring, potentially as a combined result of CL, the surgical procedure(s) and cryotherapy. LC016 had mild atrophic scarring, perhaps reflecting earlier biopsy, smaller lesion size and use of punch vs. elliptical excision biopsy. Two participants (LC003, LC004) also had wound infection post biopsy which may have contributed to the resultant scarring. Measurement of the scar area at final follow up confirmed reduced scarring in cohort 2 (punch biopsy) compared to cohort 1 (excision biopsy) (**Fig3m**). Scar size was comparable between females and males across both cohorts (females: median, 200 days, range 56-425; males: median 88 days, range 56-315). The degree of scarring and potential contributory factors are summarised in **Extended Data 2**.

### Safety and adverse events

The study recorded no grade 3 or Serious Adverse Events. No AEs were reported during the biting phase of the study. 1 participant had a grade 1 AE (exudate from scar), and 2 participants had a grade 2 AE (both wound site infections; **Fig. 3n** and **Extended Data 2**).

Wound infections were associated with itch and scratching but showed no evidence of cellulitis. For all volunteers, full blood count, liver function tests, urea and electrolytes and C-reactive protein (CRP) were taken at baseline and follow-up, with no significant differences noted (**Extended Data 5**). A minimal inflammatory response was noted, and any changes observed in leucocyte count, CRP, biochemistry and liver and renal function tests remained within normal range and were deemed not clinically relevant. All volunteers remained seronegative (rK39) throughout follow up. Lymphadenopathy of the epitrochlear and axillary lymph nodes was absent in all volunteers. As previously used in our FLYBITE study ^29^, additional safety outcomes were collated using an electronic participant-submitted visual analogue score diary card that recorded on a 1-10 scale participants subjective perceptions of the level of itch, pain, erythema, swelling, malaise, myalgia, fever and nausea (**Fig. 3o and Extended Data 6**). Summed VAS per participant were not dissimilar to those reported after the bite of uninfected sand flies ^29^, suggesting that *L. major* infection per se has minimal impact over the period VAS measurements were recorded. Participants also completed validated quality of life questionnaires throughout the study, with the Dermatology Life Quality Index (DLQI) used to measure the impact of skin changes and the Generalized Anxiety Disorder 7 score (GAD-7) used to measure any mood disturbances. The mean change in score for participants who sustained a successful bite for DLQI (30-point scale), and GAD-7 (21-point scale) were 1.92±2.54 and -0.17 ±1.94 respectively. These changes are below the minimal clinically important difference (MCID) indicating that this CHIM was generally well tolerated ^32,33^.

### Immune landscape of CL lesions

Histologically, lesions showed one or more characteristic features, including a dense lympho-histiocytic infiltration extending from the papillary into the reticular dermis, acanthosis with elongation of rete ridges (e.g. LC007, LC016), patchy hyperkeratosis (e.g. LC008), and occasional unorganised granulomas (e.g. LC025). Compact organised granulomas with epithelioid cells and / or Langhans giant cells were not seen. Extensive collagen fibres were observed in most cases (**Extended Data 4**). In recurrence biopsies, dermal infiltration was enhanced and reached the hypodermis. CD4^+^ and CD8^+^ cells were readily detected in all biopsies at variable ratios and overall CD8:CD4 ratio positively correlated with lesion duration (**Fig. 4a, b, e, and f** and **Extended Data 7**). CD4^+^CD8^+^ cells were observed (**Fig. 4b**), consistent with other reports ^34,35^. CD14^+^ monocytes and CD68^+^ CD14^+^ monocyte-derived macrophages ^36^ generally out-numbered CD68^+^ dermal macrophages, but this was not consistent across all participants and there were no significant correlations with lesion duration (**Fig. 4a, c and e** and **Extended Data 7)**. Parasitism was largely but not exclusively confined to CD14^+^, CD68^+^ and CD14^+^CD68^+^ cells, with more variability observed in cohort 2 (**Fig. 4g**). There was no correlation between infected cell type and duration of infection (Spearman’s test). CD66b^+^ neutrophils were infrequent and mostly confined to sites of active ulceration and epidermal breakdown (**Fig 4a and d** and **Extended Data 7**). CD66b^+^ cells were rarely parasitised but were observed near infected CD68^+^ cells (**Fig. 4d**). CD20^+^ B cells were scarce and sparsely dispersed but more abundant in recurrent lesions (**Fig. 4a and e** and **Extended Data 7**).

**Figure 4.**
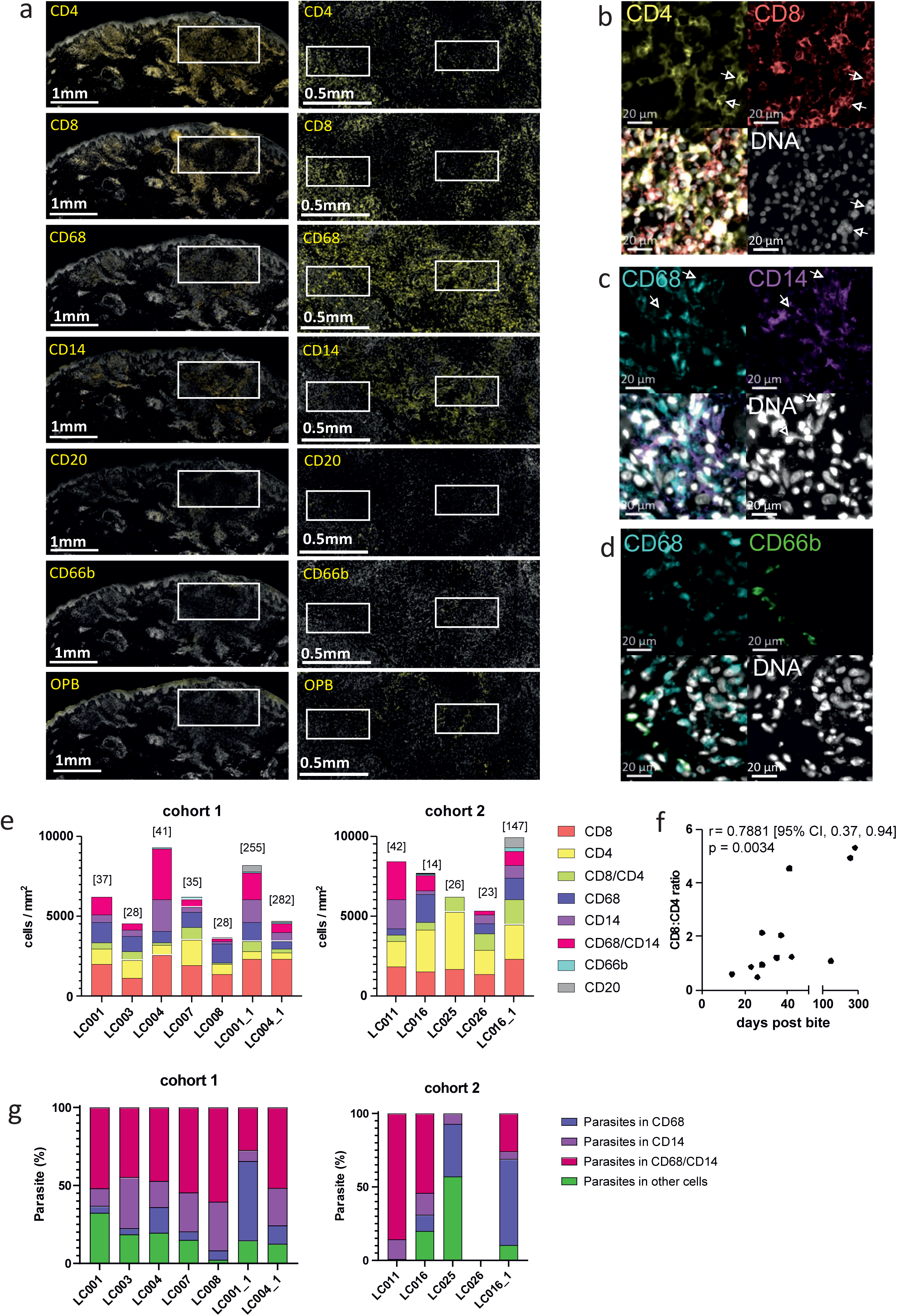
Inflammatory response during following *L. major* challenge. **a,** Immuno-histological detection (yellow) of CD4, CD8, CD68, CD14, CD20, CD66b and parasites (OPB). Sections counterstained for nuclei (YOYO1, white). Scale bar = 1mm (left images) and 0.5mm (white box; right images). Higher magnification images for white boxes shown in right panel are provided in **Extended Data 4**. Representative sections from LC001 are shown. **b**, Representative images of CD4 (yellow), CD8 (red), DNA (white) and merged image. CD4^+^CD8^+^ cells are indicated by arrowheads. **c**, Representative images of CD68 (blue), CD14 (red), DNA (white) and merged image. Infected CD14^+^CD68^+^ cells are indicated by arrowheads. **d**, Representative images of CD68 (blue) and CD66b (green), DNA (white) and merged image. Uninfected CD66b^+^ cells are seen adjacent to heavily parasitised CD68^+^ cells. **e,** Quantitation of cellular infiltrate across all participants based on IHC. Second biopsies are denoted participant number followed by _1. Data are shown in stacked bar format with time of biopsy (days post bite) shown in parenthesis above bar. **f,** Correlation between time post biopsy and CD8:CD4 ratio. Data were analysed using non-parametric Spearman two tailed test. **g**, Proportion of total parasites found in CD14^+^, CD68^+^, CD14^+^CD68^+^ or in other cell types. Total number of parasites counted ranged from 327 to 38693, except for LC025 where only 14 parasites were detected. Data are shown in stacked bar format.

We used Visium spatial transcriptomics (10X Genomics), a skin scRNAseq dataset^37^ and the Cell2Location^38^ prediction tool to further interrogate the immune landscape in three volunteers (LC001, LC003 and LC008) whose biopsies included histologically normal skin. We collected data from 20,241 55 μm-diameter Visium spots from across 12 FFPE sections (four per individual) with a median gene content of 2000 genes per spot. We used t-SNE to visualise spots coloured by Louvain clustering (**Fig 5a** and **Extended Data 8**) and generated a spatially resolved transcriptomic map of each lesion and adjacent tissue based on the 13 clusters identified (**Fig. 5a-c** and **Extended Data 8**). Cell deconvolution and transcript abundance identified key immune and stromal cell subsets associated with each cluster (**Figure 5d and e** and **Extended Data 9 and 10**). Cluster 2 (herein the lesion “core”) had abundant myeloid DC2, MigDC, LC1, Macro1, Macro2, and monocytes, as well as Tc, Th and Tregs, was enriched for interferon-inducible genes (*CXCL9*, *GBP5*), *LYZ* and Ig transcripts, effector and regulatory cytokines (*IFNG*, *TNF*, *IL-10*, *IL1B*), and was proportionally over-represented in lesion compared to healthy tissue (**Fig 5c and Extended Data 9a**). Within cluster 2, we also observed spatial co-occurrence of cell types suggestive of further spatial heterogeneity (**Fig. 5f**). For example, pericytes, MigDC, Th, ILC1_3, ILC1_NK, LC1, Tc, Treg, and vascular endothelial cells (VE1) were highly correlated, suggesting common pathways for recruitment, but were strongly anti-correlated with podoplanin-expressing F1 fibroblasts and Mono. Clusters 9 and 10 were enriched for keratinocytes, melanocytes, Langerhans cells (LC1, LC2, LC3) and ILC2, and together with cluster 5 and 12 (*CST6*) spatially defined the intact epidermis and epidermal/dermal border. Cluster 7 mapped to the “ulcer” and comprised a mixture of myeloid cells and lymphocytes, with differentiated KC, DC2 and Tregs, and a mixed gene signature including keratins, S100 proteins and collagens. Epidermal disruption was evident, with reduced expression of the basal epidermal marker *KRT5* and *LOR* (loricin, a major component of terminally differentiated epidermal cells). Of note, *LOR* mRNA was less abundant in the epidermis overlying the secondary lesion core in LC001, suggesting a less well differentiated epidermis at this site and consistent with a thickening of the stratum spinosum relative to adjacent tissue (**Extended Data 9a**). Cluster 3, mapped to the deep dermis / hypodermis, contained Macro2 and was notable for genes associated with lipid metabolism (*FABP4*, *SCD*, *PLIN1*, *GOS2*) consistent with the presence of adipose tissue, whereas dermal cluster 4 had smooth muscle cell markers (*MYL9*, *TAGLN*, *ACTA2*) suggestive of proximity to hair follicles. Dermal cluster 0 was rich in mRNAs for extracellular matrix genes (*COL1A1*, *COL1A2*, *COL3A1*, *DCN*) and *MMP2* with F2 fibroblasts the dominant stromal population. Cluster 1 was notable by an absence of key defining genes and cluster 11 (*CCL21*, *LYVE1* and various myeloid and T cells) comprised relatively few spots.

**Figure 5.**
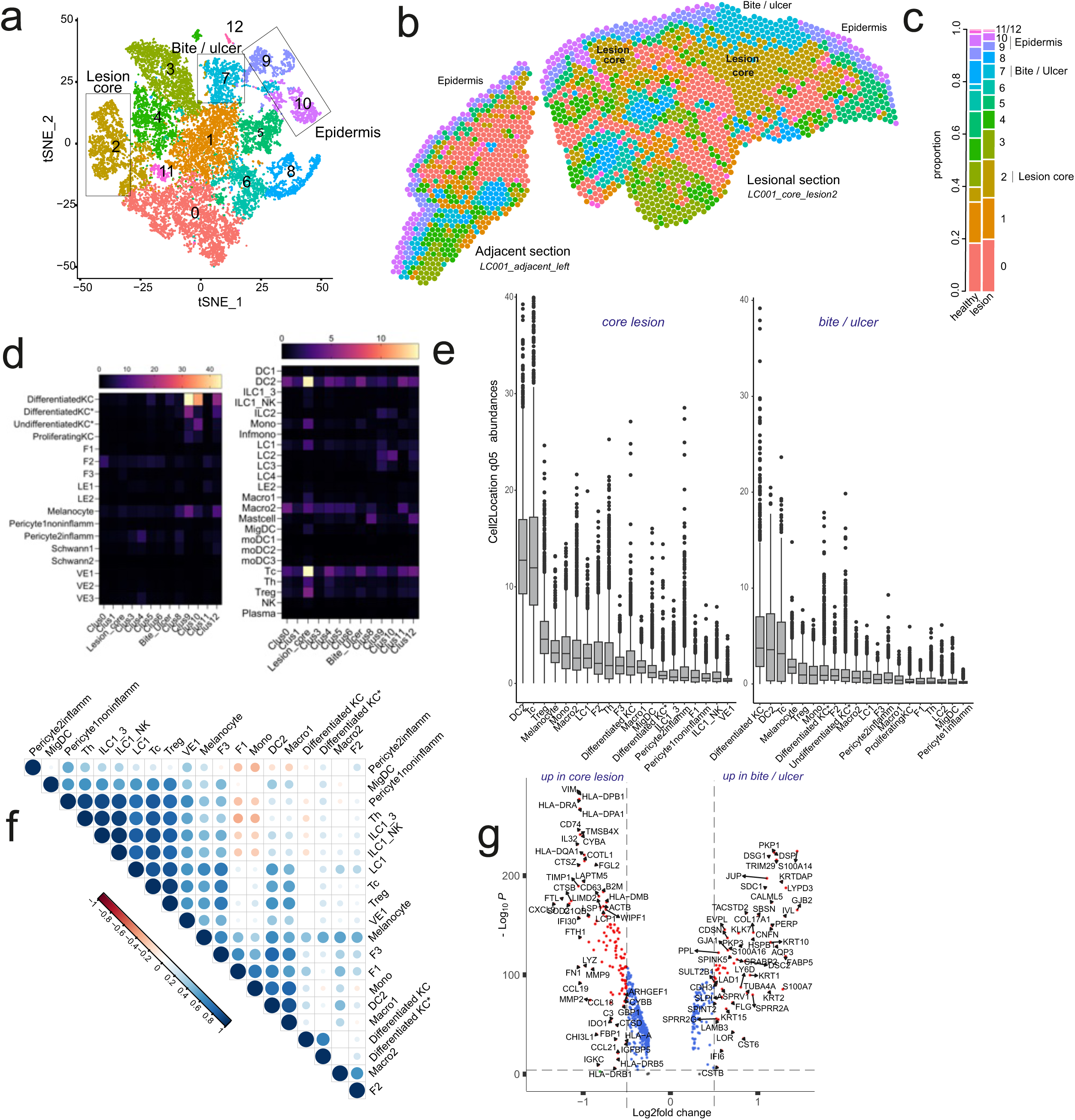
Transcriptomic landscape of early *L. major* lesions. FFPE sections from LC001, LC003 and LC008 were processed for Visium spatial transcriptomics with cell deconvolution performed using cell2location based on skin cell types identified by Reynolds et al.^37^ **a,b,** Clustering of spots reveals 13 clusters in UMAP space (a) and with discrete spatial locations (b). Cluster locations are mapped to lesion and adjacent sections from LC001. Mapping to LC003 and LC008 and additional sections from LC001 are shown in **Extended Data 8**. **c,** Proportion of spots attributed to each cluster in healthy and lesion tissue. Data are pooled across all participants / sections. **d**, Heat map representation of cellular abundances by cluster as determined by cell2location using the Reynolds et al reference dataset. Scale represents predicted 5% quantile abundances **e**, Box and whiskers plots representing cellular abundances / spot as in d for lesion core (cluster 2) and ulcer (cluster 7). Data are shown for top 20 most abundant cell types. f, Pair-wise Pearson’s correlations are represented as a correlation plot between cell types to infer spatial co-localization **g,** Volcano plot of differentially expressed genes (log2FC>1.5 and FDR=0.05) comparing lesion core with ulcer.

Given their leucocyte rich composition, we compared the lesion core and ulcer in more detail. We identified 134 genes that were differentially expressed (5% FDR; >1.5FC; **Figure 5g and Extended Data 10**) between lesion core (80 UP) and ulcer (54 UP). mRNAs with greater abundance in the lesion were related to antigen processing and presentation (e.g. *HLA-DRA*, *HLA-DPA1*, *CD74*), metalloproteinase activity (*MMP2*, *MMP9*, *TIMP1*) and multiple cytokines and chemokines (*CCL5*, *CCL18*, *CCL19*, and *CCL21*), the metabolic checkpoint enzyme *IL-4l1* linked to the induction of regulatory T cells and reduced proliferation and function of effector T cells ^39^, and *IL-32* recently shown to be abundant in late CL lesions and associated with IDO1 and PD-L1 expression ^40^. Pathway analysis^41^ identified multiple immune-related pathways in the lesion whereas pathways in the ulcer were related to epidermal remodelling (**Extended Data 10)**. The enrichment of a neutrophil degranulation pathway in the lesion core was discordant with the number of neutrophils identified by CD66b staining (**Figure 3**) and may reflect the expression of pathway-associated genes by monocytes / macrophages in inflammatory settings (e.g. *CTSG*, *MPO*, *CD63*, *MMP9*).

To further characterise the lesion core, we identified four spatially distinct sub-clusters or niches (**Fig. 6a and b**). Lesion core sub-cluster 0 was interspersed with sub-cluster 3, with abundant mRNA for the antimicrobial and monocyte / T cell chemoattractant chemokines *CCL22* and *CXCL9* ^42,43^ (**Figure 6b and c**). mRNAs whose abundance correlated with *CXCL9* formed a STRING network enriched for GO terms related to IFNγ response, antigen presentation, neutrophil activation, and leucocyte adhesion (**Extended Data 10**). Sub-cluster 1 had abundant mRNA for *CCL19* (**Fig. 6b and d**) and B cells (*IGKC*) and was located at the core periphery, as well as deeper in the dermis. Sub-cluster 2 contained presumptive fibroblasts (*GREM1*, *MMP2* and *PI16*) and was localised to the periphery (**Fig. 6b and e**). Sub-cluster 3 was characterised by *CHI3L1* (a chitinase-like protein with broad ranging activity in inflammation, tissue repair and macrophage polarisation ^44^; **Fig. 6b and f**), NUPR1 (strongly expressed by basophils and neutrophils), FBP1 (fructose-1,6-bisphosphatase 1, recently identified as a marker of human M1 polarisation ^45^) and MT1G (metallothionein 1G, associated with cancer associated TREM^hi^ macrophages ^46^ and a pleiotropic regulator of myeloid cell function ^47^).

**Figure 6.**
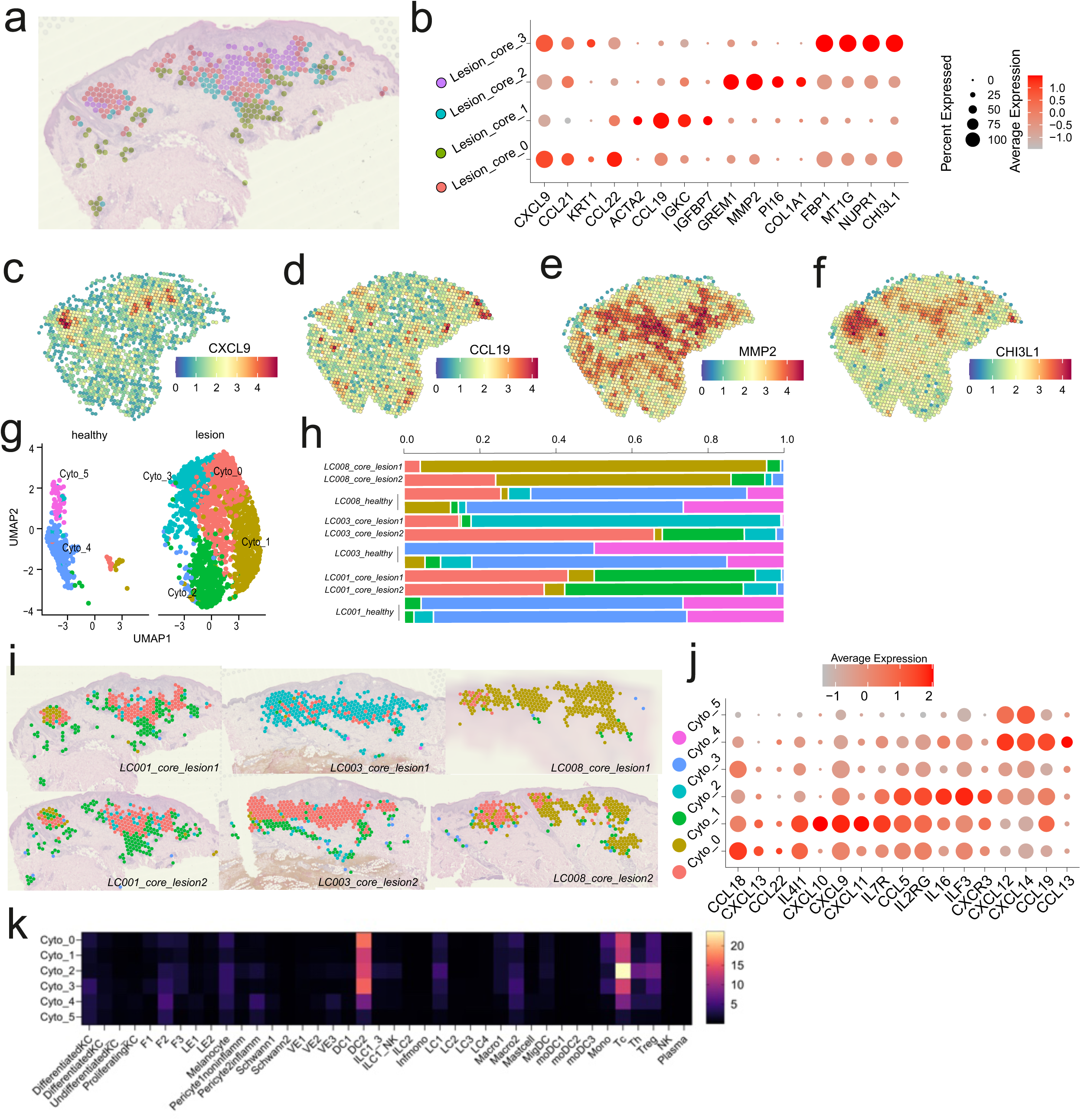
Discrete spatial niches in the lesion core. **a,** Sub-division of the lesion core cluster (Fig 5 a and b) into 4 sub-clusters. **b,** Bubble plot showing top genes associated with lesion core subclusters. **c-f,** Expression of CXCL9 (c), CCL19 (d), MMP2 (e) and CHI3L1 (f) mapped to LC001 for visualisation. **g-h,** Re-clustered lesion core (Lesion_core_0/1/2/3 as in a,b) based on chemokine and cytokine family genes visualised in UMAP space as Cyto_0/1/2/3/4/5 shown separately for healthy vs lesional tissue (g) with proportion of spots for each healthy and lesion shown individually for each section / participant (h). **i**, Cyto_0/1/2/3 clusters mapped to each participant for visualisation. **j,** Bubble plot showing key genes associated with each cytokine / chemokine-based cluster. Scale bar shows average expression. **k**, Correlation matrix (Pearson’s) for predicted cell abundances and Cyto_0/1,2/3 clusters, based on data from all sections / participants.

Given the importance of chemokine and cytokine signalling in anti-leishmanial immunity, we re-clustered these data (i.e. Lesion_core_0-3) based on expression of these molecules alone (**Fig. 6g-j**). 6 clusters were visualised in UMAP space (**Fig. 6g**), with Cyto_0/1/2/3 largely confined to the lesion, albeit with some variability between participants and across serial sections (**Fig. 6h and i**). Cyto_0 (*CCL18*) and Cyto_1 (*CXCL9*, *CXCL10* and CXCL11) formed the central region of the lesion core, whereas Cyto_2 (*CCL5, CCL19, IL-16, IL2RG, CXCR3*) was largely confined to the borders (**Fig 6i**). Although Tc were most abundant in Cyto_2, Th were selectively associated with this cluster in keeping with the chemoattractant role of CCL5 and IL-16 (**Fig 6k**). Cyto_3 generally showed lower mRNA abundance for most cytokines and chemokines and was mainly derived from one section (**Fig6i**). Cyto_4 and Cyto_5 (*CXCL12*, *CXCL14*, *CCL13*) mapped mainly to healthy tissue and comprised F2 fibroblasts and various myeloid and lymphocyte populations in low abundance (**Fig 6g**). Hence the lesion core, whether sub-clustered in an unbiased manner (**Fig 6a and b**) or using only cytokines/chemokines genes (**Fig 6g-k**) displays clear and hitherto unrecognised functional compartmentalisation and cellular heterogeneity.

## Discussion

We report clinical, parasitological, and preliminary immunological data from the first CHIM of sand fly transmitted CL caused by *Leishmania major*. We demonstrate that this model has a take rate comparable to that of other challenge models and is safe and well-tolerated by participants, suggesting its suitability for evaluating new *Leishmania* vaccines, pre- or post-exposure therapies, and the immunopathogenesis of CL in humans.

Previous human infection studies with *Leishmania* using needle challenge allowed lesion progression through to self-cure, with most participants developing ulcerated lesions by 60 days post inoculation ^25^. In contrast, our study was designed to estimate take rate using a newly identified and cGMP manufactured *L. major* strain and a laboratory-reared colony of *P. duboscqi* and to utilize therapeutic biopsy ^48^ to excise the lesion early in development. The latter approach was guided by a public involvement (PI) exercise ^30^ and a desire to avoid extensive lesion development. Increased understanding of disease mechanisms facilitated by lesion biopsy was also recognised by our PI group as a desirable outcome ^30^. The results from the first cohort demonstrated a take rate of 83% for all participants (100% for those receiving a bite). Recruitment was extended to increase confidence in the take rate and to incorporate changes aimed at minimising lesion size and scarring. These contributed to reduced scaring, but the smaller lesions and earlier time points of biopsy reduced overall take rate. Nevertheless, this aligns with other CHIMs and the common belief that it is wise to avoid an overwhelming force of infection in CHIM-based vaccine trials ^49^. However, if higher take rates were desired (e.g. for discovery research on immune mechanisms or for drug evaluation) this might be achievable by increasing the number of infected sand flies or extending the exposure time.

Target Product Profiles for CL vaccines propose an efficacy of 70% ^8^. Based on a dichotomous endpoint (lesion vs no lesion), that we view as the gold standard for an effective CL vaccine, and our minimal estimated take rate, the current CHIM protocol could provide efficacy data with ∼50 participants. This sample size, coupled with the rapidity of lesion development likely facilitated by sand fly transmission, would make vaccine evaluation highly cost effective compared to field trials in endemic countries. Relaxing the endpoint definition e.g. by excluding parasitological confirmation would increase take rate and reduce sample size further. Other measures of vaccine efficacy can be considered, but with likely consequences for sample size, cost, and burden on CHIM participants. For example, lesion parasite load was highly variable, reflecting i) the early time points studied, ii) sampling method, iii) the nature of vector transmission (where quantity of inoculated parasites ^50,51^ and the number of bites are hard to control), and iv) inter-individual variation in immune response. qPCR of sequential microbiopsies ^52^ might mitigate against variation in parasite load but could impact on lesion progression. Similarly, assessment of vaccine-induced reduction in CL scar would require omission of the therapeutic biopsy and participant consent to allow CL to take its full course.

CL is known to result in scarring and as expected some degree of scarring was evident in all participants who developed a lesion, underwent biopsy and / or received cryotherapy. Reducing the area of exposure and early punch biopsy resulted in better cosmesis based on final scar size but the study was not designed to evaluate this formally. Conversely, wound infection, later excision biopsy and cryotherapy all appeared to contribute to the resultant scarring. The low frequency of wound infections was within acceptable limits and could potentially have been mitigated by use of antimicrobial washes (e.g. Dermol 500) and antihistamines. All suspected wound infections responded quickly to treatment. Regarding mechanisms of scar formation, our preliminary analysis demonstrates significant epidermal remodelling and abundant mRNA for mediators of fibrosis such as MMP9, previously associated with clinical outcome in New World CL and VL ^54,55^. However, in the absence of a suitable control arm it is difficult to formally distinguish between scarring attributable to CL and that resulting from the therapeutic intervention. This could be considered in future, further adapting this CHIM to generate new insights into CL scar formation.

The study has some additional limitations. In two of the three participants we saw recurrence of lesion development after therapeutic biopsy, but each responded well to cryotherapy. In one case, steroid administration may have been a precipitating factor, suggesting that intra-lesional steroid injection should be contraindicated. Although the possibility of future recurrence cannot be completely excluded, the risk appears low. Relapse of *L. major* due to HIV-associated or elective immunosuppression has not been reported and unlike in mice, *L. major* does not appear to persist in the scars of CL patients ^56^. All participants in the study were white and future CHIM studies should consider the potential impact of race, ethnicity, and environment in relation to the desired endpoint. Previously identified constraints to implementing CHIM studies in lower- and middle-income settings ^57^ apply equally to this CHIM, and additional factors related to vector diversity and disease heterogeneity have been discussed extensively elsewhere ^58,59^. Finally, we preformed therapeutic biopsy in a narrow time window, consistent with the study objectives, but biopsy timings could be readily altered to accommodate different study objectives e.g. to study innate immune responses or more advanced immunopathology.

*L. major* infection in mice was instrumental for establishing the Th1 / Th2 paradigm of cellular immunity ^60^, understanding vector contributions to pathogenesis ^26,61^ and the contribution of diverse myeloid cell populations to CL chronicity ^62^. Studies on human immunity to *L. major* are, however, less comprehensive. Cure is associated with a strong Th1-mediated IFNγ response that promotes self-healing ^63,64^. Immuno-histochemical studies have described changes in adhesion molecule and MHC expression associated with infiltration by CD4^+^ and CD8^+^ T cells ^65^. Other studies ^66,67^ report detection of mRNA for *IFNG*, *TNF* and *IL6*, less frequent detection of *IL4* and suggest that IL-10 might support an immunosuppressive milieu ^66^. However, these and similar studies provide an incomplete picture of the immune landscape in CL lesions and were limited to patients with established lesions presenting for treatment. CHIM biopsies provide a unique opportunity to examine early lesion progression after natural infection. In the initial analysis presented here, we highlight how immune responses differ between the lesion core and ulcer, indicative of the independence of anti-parasitic and wound healing responses. We observed an unexpectedly high frequency of CD8^+^ T cells particularly in recurrent or late lesions, the predominant parasitism of macrophages and monocyte derived macrophages and that DC2 are predicted as the dominant myeloid cell type in the lesion core. We identified diverse immune niches with selective chemokine and / or cytokine expression linked to cellular composition. Similarly, we observed various stages of epidermal remodelling associated with ulceration or underlying inflammation. Collectively, these data provide a blueprint for future studies seeking to determine how microenvironment shapes the outcome of infection over time, to identify potential correlates of protection and pathology, and to inform the development of vaccines, drugs, and host-directed therapies though mechanistic understanding of immunity.

In conclusion, we have developed a safe and highly effective CHIM that recapitulates natural transmission of *L. major.* Notwithstanding the heterogeneity of the leishmaniases ^1^, epidemiological and experimental evidence supports at least some degree of natural or vaccine-induced cross-species protection ^12,68^. Hence, this CHIM will have broad utility for assessing vaccines designed to target many forms of leishmaniasis, including visceral leishmaniasis. Our analyses also highlight functional compartmentalisation of immune responses at the site of infection and provide a resource to comprehensively map the immune landscape of CL lesions in humans.

## Online methods

### Ethics and Inclusion statement

The study was approved by the UK Health Research Agency South Central - Hampshire A Research Ethics Committee (IRAS Project ID:286420; 20/SC/0348) and the Hull York Medical School Ethical Review Committee (Approval No. 2073). The study sponsor was University of York. The study was prospectively registered at ClinicalTrials.gov Identifier: NCT04512742. Participants in this study were both male and female sex.

### Vectors and parasites

*Phlebotomus duboscqi* originating in Senegal were maintained in the insectary of the Department of Parasitology, Charles University in Prague, under standard conditions (26°C on 50% sucrose solution, humidity in the insectary 60-70% and 14 hour light/10 hour dark photoperiod)(15). Colonies were negative by PCR for sand fly-associated phleboviruses (including Sandfly Fever Sicilian Virus group, Massilia virus and Toscana Virus) and Flaviviruses (targeting a conserved region of the NS5 gene). As required, batches of approximately 200 sand flies were shipped at 3 to 5 days of adult development to the University of York in a humidity and temperature-controlled sealed unit.

After arrival, the sand flies were maintained on a sugar solution for 24 hours, and subsequently starved to encourage later blood feeding. Twelve to fifteen days prior to a scheduled biting day, sand flies were infected using a membrane feeder (Hemotek) containing rabbit blood mixed with 10^6^ promastigotes per ml of a recently described strain of *L. major,* isolated in Israel and manufactured to cGMP (MHOM/IL/2019/MRC-02; ^28^). Three to five days before a scheduled biting day, a sample of engorged sand flies was dissected to ensure infection rates above 90% by standard methods. On the day of the biting study, a sand fly biting chamber (Precision Plastics Inc, Maryland, USA) was loaded with 5 female sand flies and placed on the participant’s arm for 30 minutes. Biting failure was defined by absence of i) participant-reported biting sensation during and immediately after biting, ii) sand fly biting activity as noted by clinical investigators (including inspection of video and photography during biting), iii) bite compatible lesions by dermoscopy or photography immediately after biting, and iv) any macroscopic evidence of blood in sand fly abdomen at end of biting period. No discrimination was made between partially and fully fed flies. Volunteers with suspected biting failure were followed up until day 28 and then replaced in the study (with a final follow-up at 6 months).

### Clinical procedures

All clinical procedures and SOPs are provided in Extended Data 3 (Study protocol).

### Histology and qPCR

Biopsies were obtained using either a standard elliptical excision biopsy or punch biopsy. Immediately following biopsy, the tissue was cut into 3 pieces (50% for histology, 25% for qPCR and 25% for immunological analysis). Extraction of total DNA was performed using DNeasy tissue isolation kit (Qiagen) according to the manufacturer’s instruction. Parasite quantification by quantitative PCR (qPCR) was performed in a Bio-Rad iCycler & iQ Real-Time PCR Systems using the SYBR Green detection method (SsoAdvanced™ Universal SYBR® Green Supermix, Bio-Rad, Hercules, CA). Primers targeting 116 bp long kinetoplast minicircle DNA sequence (forward primer (13A): 5′-GTGGGGGAGGGGCGTTCT-3′ and reverse primer (13B): 5′-ATTTTACACCAACCCCCAGTT-3′) were used. ^69^ One microlitre of DNA was used per individual reaction. PCR amplifications were performed in triplicates using the following conditions: 3 min at 98 °C followed by 40 repetitive cycles: 10 s at 98 °C and 25 s at 61 °C. PCR water was used as a negative control. A series of 10-fold dilutions of *L. major* promastigote DNA, ranging from 1 × 10^6^ to 1 × 10 parasites per PCR reaction was used to prepare a standard curve. Quantitative results were expressed by interpolation with a standard curve. To monitor non-specific products or primer dimers, a melting analysis was performed from 70 to 95 °C at the end of each run, with a slope of 0.5 °C/c, and 5 s at each temperature.

Samples for FFPE were placed in 4% formaldehyde (Thermo Scientific; 28908) for 24 hours at 4°C. They were then paraffin embedded in histosette I tissue processing/embedding cassettes (Simport; M490-5) on the Leica ASP300S Fully Enclosed Tissue Processor (Leica Biosystems) and embedded on the Leica EG1150 H Modular Tissue Embedding Center (Leica Biosystems). Blocks were chilled prior to sectioning. 7μM sections were cut on a Leica Wax Microtome and placed into a water bath set to 45°C for 15 seconds. Sections were then collected onto Superfrost slides (ThermoScientific; J1800AMNZ) and allowed to dry overnight at RT. Slides heat fixed at 60°C for 2 hours in a sterilising oven (Leader Engineering; GP/30/SS/250/HYD, 08H028). Slides were allowed to cool down and then deparaffinised with Histoclear II (SLS; NAT1334) for 5 minutes. Slides were equilibrated in 95% Ethanol for 3 minutes, 70% Ethanol for 3 minutes and distilled water for 3 minutes.

### Haematoxylin and Eosin

Slides were then stained in Harris Haematoxylin (ThermoScientific; 6765001) for 3 minutes and then rinsed in tepid water for 5 minutes. Slides were dipped once in 1% acid-alcohol (HCl-EtOH; Sigma; 30721-2.5L-M; Fisher Scientific; E/0650DF/C17) and then equilibrated in distilled water for 3 minutes. Slides were then stained with 1% Eosin (Sigma-Aldrich; E4382-25G) for 3 minutes and then dipped in 50% Ethanol 10 times. Slides were then equilibrated in 70% Ethanol for 3 minutes, 95% Ethanol for 3 minutes and 100% Ethanol for 3 minutes. Slides were then cleared in Histoclear II (SLS; NAT1334) for 9 minutes. Slides were then mounted with Dibutylphthalate Polystyrene Xylene (DPX; Sigma-Aldrich; 06522-500ML) and coverslipped with 22 x 50 mm cover slips (SLS; MIC3226). Slides were dried overnight before scanned on the Zeiss Axioscan Z1 (Zeiss).

### Immunohistochemistry

Slides were subjected to heat mediated antigen retrieval in 10mmol/l sodium citrate buffer (pH6). Sections were incubated with 1% BSA, 0.1% cold fish gelatin, 0.1% triton x100 in PBS for 1 hour at room temperature to block non-specific immunoglobulin binding. Sections were stained with primary antibodies overnight at 4^0^C, mouse anti-human CD3 (1:100, OriGene, UM500048CF), rabbit anti-CD4 (1:50, Abcam USA, Ab133616); mouse anti-CD8 (1:100, Biolegend 372902), rabbit anti-human CD68 (1:800, Abcam USA, ab213363), mouse anti-CD14 (1:200, Abcam USA, ab181470), *Leishmania* Oligopeptidase B (10µg/ml, provided by Jeremy Mottram, University of York, UK), rabbit IgG isotype control (concentration same as the primary, Abcam USA, ab172730, and mouse IgG1 isotype control (BioLegend USA, 401401). Primary antibodies were detected by F(ab’)2-goat anti-mouse IgG (H+L) cross-adsorbed secondary antibody, Alexa Fluor 555 (Thermo Fisher Scientific, USA, and A21425), donkey anti-sheep IgG (H+L) cross-adsorbed secondary antibody, Alexa Fluor 647 (Thermo Fisher Scientific, USA, and A21448) and donkey anti-rabbit IgG (H+L) highly cross-adsorbed secondary antibody, CF750 (Biotium, 20298) incubated for 30 minutes at room temperature. Subsequently sections were stained with conjugated antibodies, mouse anti-CD20 Alexa Fluor 647 (1:100, Novus, NBP-47840C), mouse anti-CD66b Alexa Fluor 647 (1:50, Biolegend, 392912), Mouse IgG1 Alexa Fluor 647 (concentration same as the conjugated primary, Biolegend 400130) and YOYO-1 (ThermoFisher, Y3601) for 1 hour at room temperature. Sections were mounted in ProLong ™ Gold antifade mountant (Invitrogen P36930). Images were acquired using Zeiss AxioScan.Z1 slide scanner. Identical exposure times and threshold settings were used for each channel on all sections of similar experiments. Quantification was performed using StrataQuest Analysis Software (TissueGnostics).

### Visium whole transcriptome spatial transcriptomics and processing

FFPE sections were cut onto 10X Genomics Visium slides with large sections being split into “lesion” and “adjacent” tissue to fit within Visium fiducial markers. Slides were processed according to the Visium Spatial Gene Expression Reagent Kits for FFPE recommended protocol v1 (10X Genomics). Briefly, slides were stained with haematoxylin and eosin, imaged, and de-crosslinked. Human probes were added overnight and then extended and released. Libraries were prepared according to the manufacturer’s instructions and sequenced using the NovaSeq 6000 platform. Raw fastq files were aligned to the human genome GRCh38 (GENCODE v32/Ensembl 98) using Space Ranger software (10x Genomics).

Associated image files were aligned onto slide specific fiducials using Loupe browser software (10X Genomics). Tissue regions were manually selected and a tissue x,y co-ordinate json file was created. Json files and image files were provided as input to the Space Ranger count() function to generate counts and align them to spatial spots. Raw counts were normalised and analysed further.

### Normalization and data integration

Seurat (v4.3.0) was used to find variable features, normalize, and scale the data using the SCTransform() function and nCount_Spatial and nFeature_Spatial was used to regress the counts. Next, spatial data for 4 sections per volunteer (3 volunteers:LC001, LC003 and LC008) were integrated into one single Seurat object containing 12 images by first selecting features for integration, SelectIntegrationFeatures() and then identifying anchors, FindIntegrationAnchors(), and finally integrated using IntegrateData(). Finally, the first 15 principal components and a resolution of 0.3 was used to obtain cluster memberships per spot. Additionally, underlying histology and Clustree ^70^ was used to visualise and choose the resolution of clustering. To exclude borderline areas between “lesion” and “adjacent” tissue, H&E images were used to exclude spots that were underlying morphologically altered or disrupted epithelium as these likely reflected the edge of the lesion. Analysis of spots underlying morphologically normal epithelium were taken to reflect “healthy” tissue for the purposes of comparative analysis. Differential gene expression was calculated first by using the minimum of the median UMI of individual objects to reverse individual SCT models as a covariate for sequencing depth using the function PrepSCTFindMarkers(). Next, Wilcoxon Rank Sum test was employed to find the features that were differentially expressed using an adjusted (Bonferroni correction) p-value threshold of 0.05. Differentially expressed gene names were submitted to StringDB (https://string-db.org/). The full STRING network (the edges indicate both functional and physical protein associations) was selected for the analysis. K-means clustering was performed within STRING to generate 3 clusters. Pathway analysis was conducted using gprofiler ^41^.

### Cell type deconvolution of Visium spots

We used Reynolds et al ^37^as a source of single-cell RNA cells from healthy and inflamed skin to model cell abundance per Visium spot using cell2location ^38^. Cell2location was used as per its recommended instructions. Briefly, fifty-thousand single-cell transcriptomes (retaining cell type annotation as per Reynolds et al) were used to model reference cell-type gene expression using cell2location’s negative binomial regression for one thousand epochs.

Spatial gene expression was then ascribed to cellular abundances based by training the cell2location model for thirty-thousand epochs. Hyperparameters N_cells_per_location and detection_alpha was selected as thirty and twenty respectively. Finally, predicted abundances (5% quantile values of the posterior distribution) per Visium spot were imported as metadata onto the Seurat object. Predicted abundances were further analysed by calculating Pearson’s correlation between cell-types to suggest colocalization.

### Quantification and statistical analysis

The study was an observational exploratory clinical study and not powered to detect differences in outcome measures between cohorts or between sex, age, or other demographic variables. Sample size was chosen on a pragmatic basis to confidently assess attack rate (lower 95% CI of ∼60%) with the minimum number of participants. Where quantitative measures were analysed, data was tested using Prism (v10.0.3; GraphPad) for normality (D’Agostino and Pearson or Shapiro-Wilks tests). Non-parametric data was analysed for correlation using Spearman’s test. Transcriptomic data was analysed using appropriate R packages (see above). No blinding was performed, but all downstream analysis of tissue samples were conducted using automated quantitative pipelines (see above).

## Data availability

The additional datasets generated, analysed, and supporting the conclusions of this study are available upon reasonable request (including a detailed proposal for its use) from VP, HA, SD, CJNL, AML, and PMK, and with agreement from the study sponsor.

Processed spatial transcriptomics data is available at 10.5281/zenodo.10018477 with instructions and code at https://github.com/jipsi/chim.

Raw transcriptomic data has been deposited in GEO (GSE263298) available from: https://www.ncbi.nlm.nih.gov/geo/query/acc.cgi?acc=GSE263298

## Reporting Guidelines

See CONSORT diagram (**Fig. 1**) and CONSORT checklist (**Extended Data 1**).

## Consent

Written informed consent for publication of pseudo-anonymised details and images was obtained from all participants.

## Acknowledgements

The authors wish to thank the clinical research nurses (Nicola Marshall, Siobhan Sutton, and Beverley Proctor), the Research and Development team at York Teaching Hospitals NHS Foundation Trust and Liz Greensted for their support and valued input. We are also grateful to Drs Tom Darton, Steve Walker and Amila Wickramasinghe for independent advice on clinical management. The participation of our volunteers is greatly appreciated.

## Author contributions

Conceptualisation by PMK, AML, CJNL, ES, CLJ and PV. Clinical and experimental studies were conducted by VP, AM, JS, BV, KVB, RW, DT, NSD, CLJ, CJNL, and AML. Study data collation and analysis was conducted by VP, HA, SD, AML, CJNL, and PMK. Transcriptomic analysis was conducted by ND. Statistical analysis was conducted by VP, HA and PMK. Writing original draft by PMK. Review & editing draft by VP, HA, SD CJNL, and AML. Writing final draft by PMK. All authors read and approved the final version of the manuscript. Funding acquisition by PMK, CJNL, AML, CLJ and PV.

## Extended Data files

**Extended Data 1. CONSORT checklist**

**Extended Data 2. Demographics, scarring and adverse events Tab 1,** Demographics. **Tab 2**, Scarring. **Tab 3,** Adverse events

**Extended Data 3. Protocol and PIS**

**Extended Data 4. H&E-stained FFPE sections from each biopsy.**

**a-n.** H&E stained FFPE sections from all biopsied participants. Boxed areas are shown at higher magnification. All scale bars represent 1mm.

**Extended Data 5. Blood biochemistry and CBC data**

Data are shown for all participants at each study visit where blood was drawn.

**Extended Data 6. Visual Analogue Scores by feature and participant**

**a-h.** VAS scores out of 10 for itch, pain, erythema, swelling, malaise, myalgia, fever and nausea. Data shown as mean and range for all participants with positive bite.

**i-s.** Summed VAS scores for each participant. Data shown as total score out of 80.

**Extended Data 7. IHC staining for immune cells populations**

**a-g**. IHC (yellow) for CD4 (a), CD8 (b), CD68 (c), CD14 (d), CD20 (e), CD66b (f) and *Leishmania* OPB (g). Left and right images correspond to boxed areas of images shown in Figure 4a right. Scale bar in all images represents 0.2mm.

**Extended Data 8. Visium spatial maps of CL lesions**

**a,** heat map representation of top 5 genes representing clusters 0-12 in Figure 5 and b. **b,** tSNE representation of clusters by lesion vs adjacent tissue. **c,** tSNE representation showing overlays for all samples studied. **d-f,** spatial mapping of clusters for all sections studied for participant LC003 (d), LC001 (e) and LC008 (f).

**Extended Data 9. Localisation of select mRNA and predicted cell types**

**a,** feature maps showing location of mRNA *LOR*, *KRT5*, *LYZ*, *S100A2*, *TNF*, *IFNG*, *IL10* and *IL1B*. **b,** Feature maps showing cell2location-predicted cells types by abundance for monocytes (Mono), myeloid dendritic cells 2 (DC2), macrophages 2 (Macro2), fibroblasts 1 (F1), cytotoxic T cells (Tc), helper T cells (Th) and regulatory T cells (Treg).

**Extended Data 10. Visium deconvolution and pathway analysis**

Tab 1, Data from Reynolds et al (Ref 37) used for cell2location deconvolution. Remaining tabs provide data underpinning Figures 5 and 6.

