## Supplementary figures and images for "Safety, effectiveness, and skin immune response in a controlled human infection model of sand fly transmitted cutaneous leishmaniasis"

### Extended Data 4

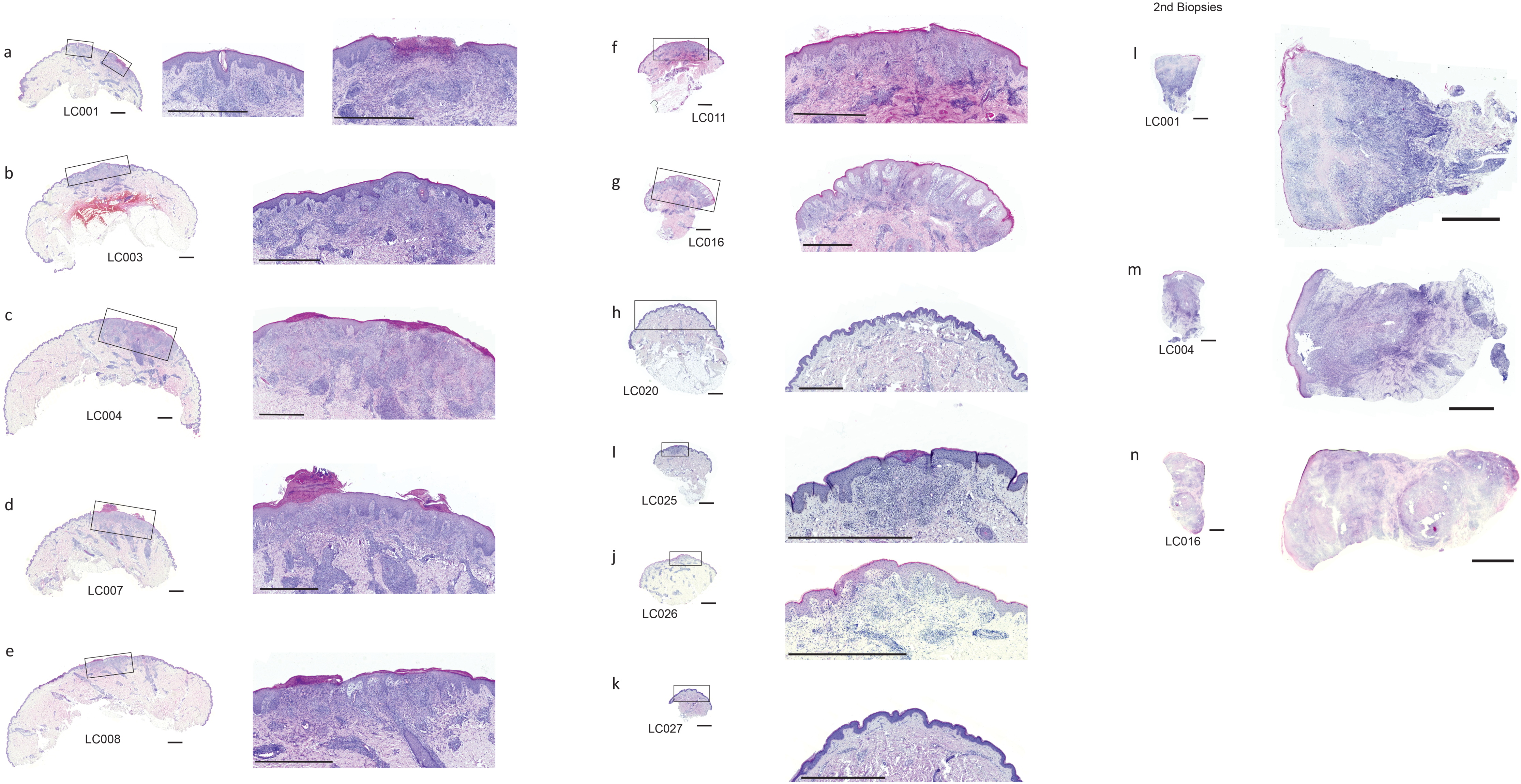

### Extended Data 7

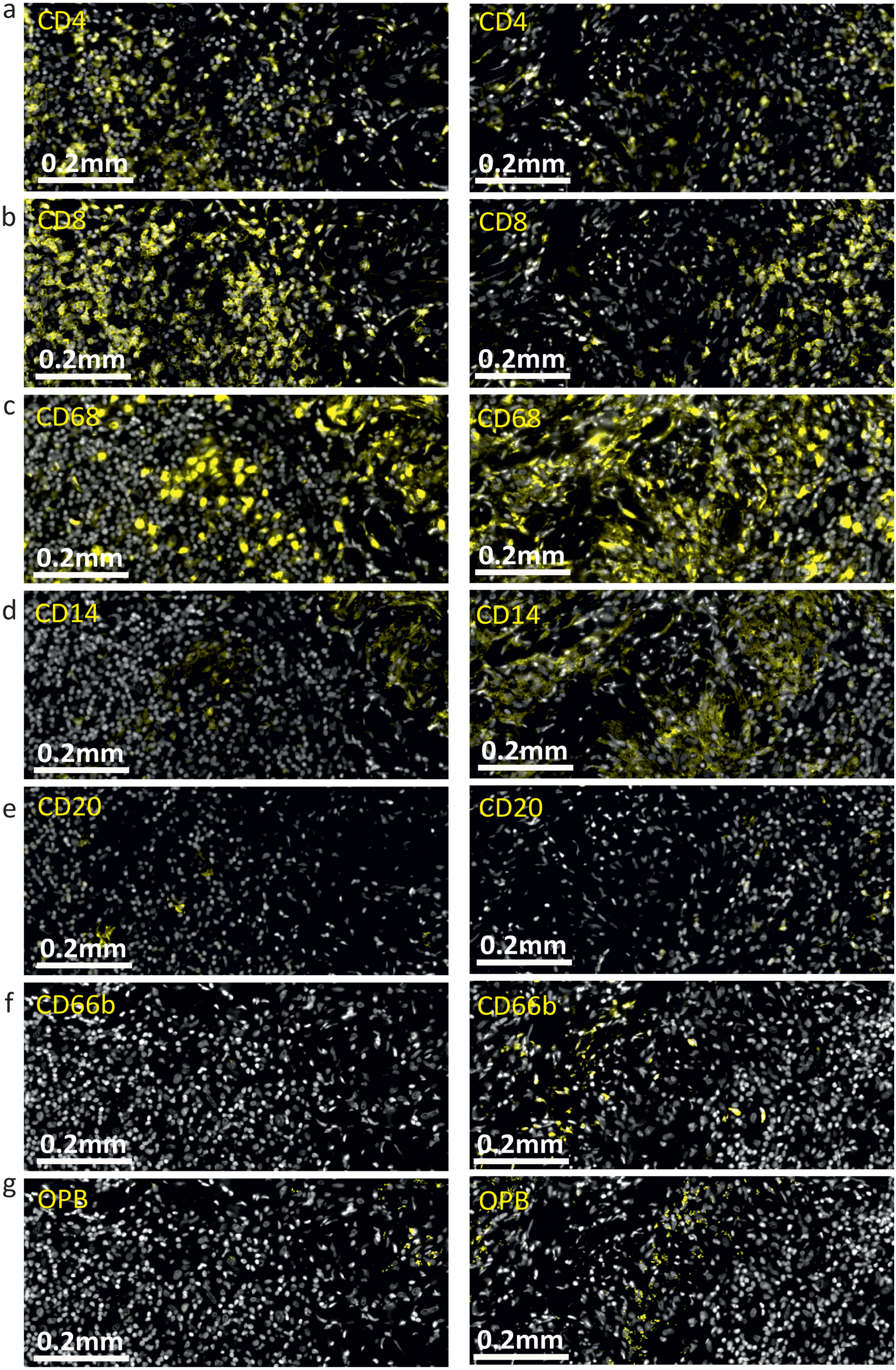

### Extended Data 8

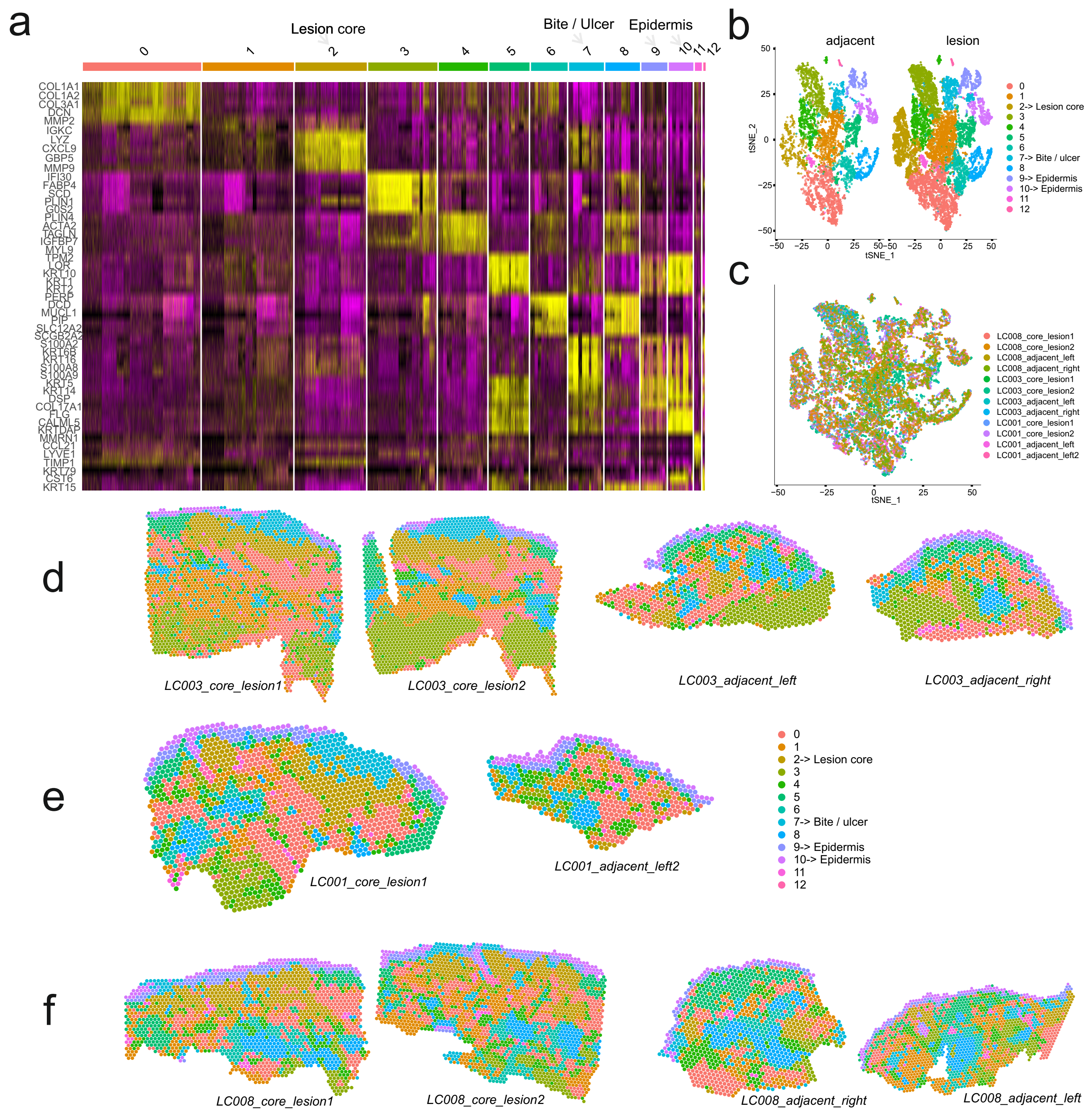

### Extended Data 9

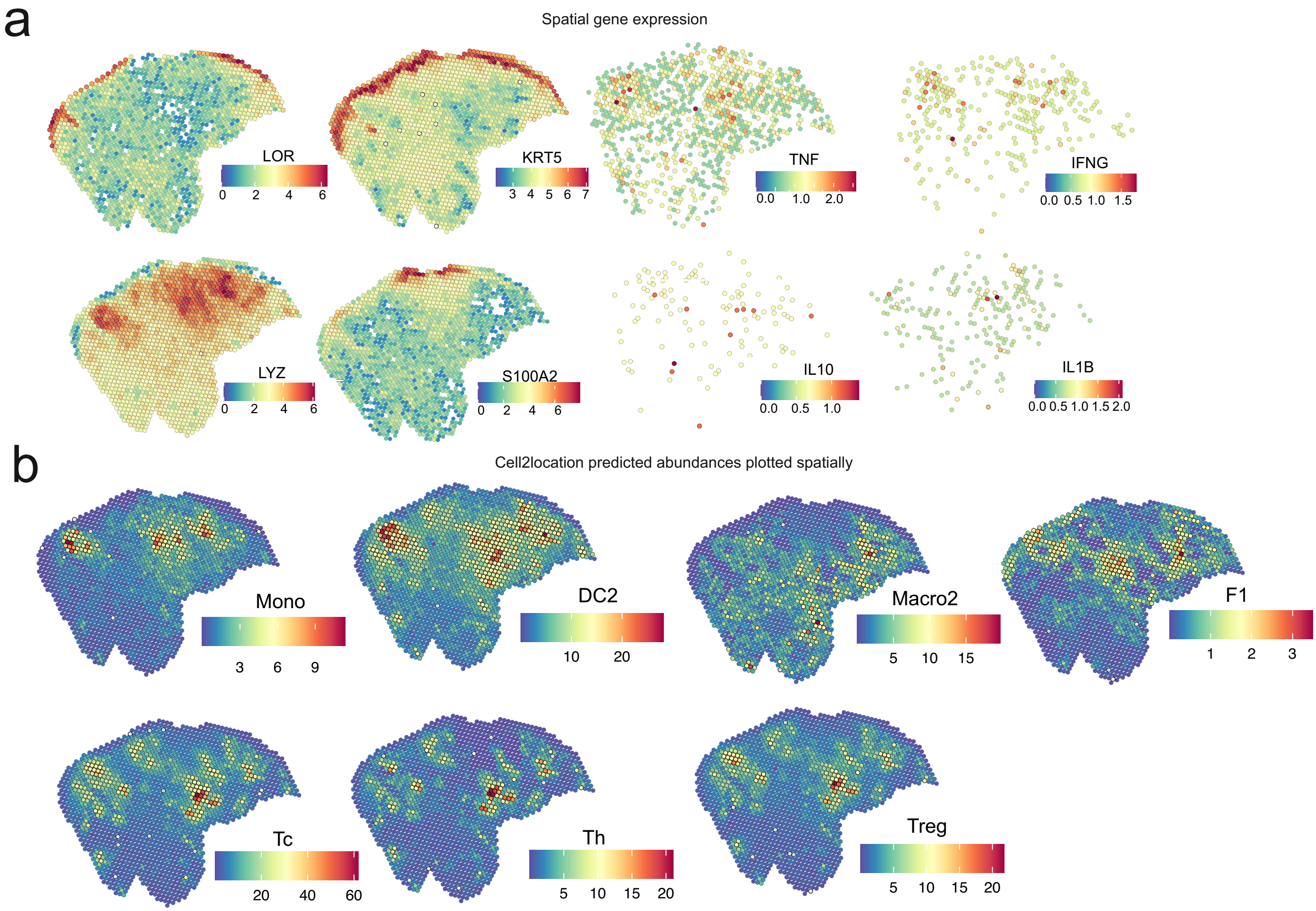
