## Extended Data 6 for "Safety, effectiveness, and skin immune response in a controlled human infection model of sand fly transmitted cutaneous leishmaniasis"

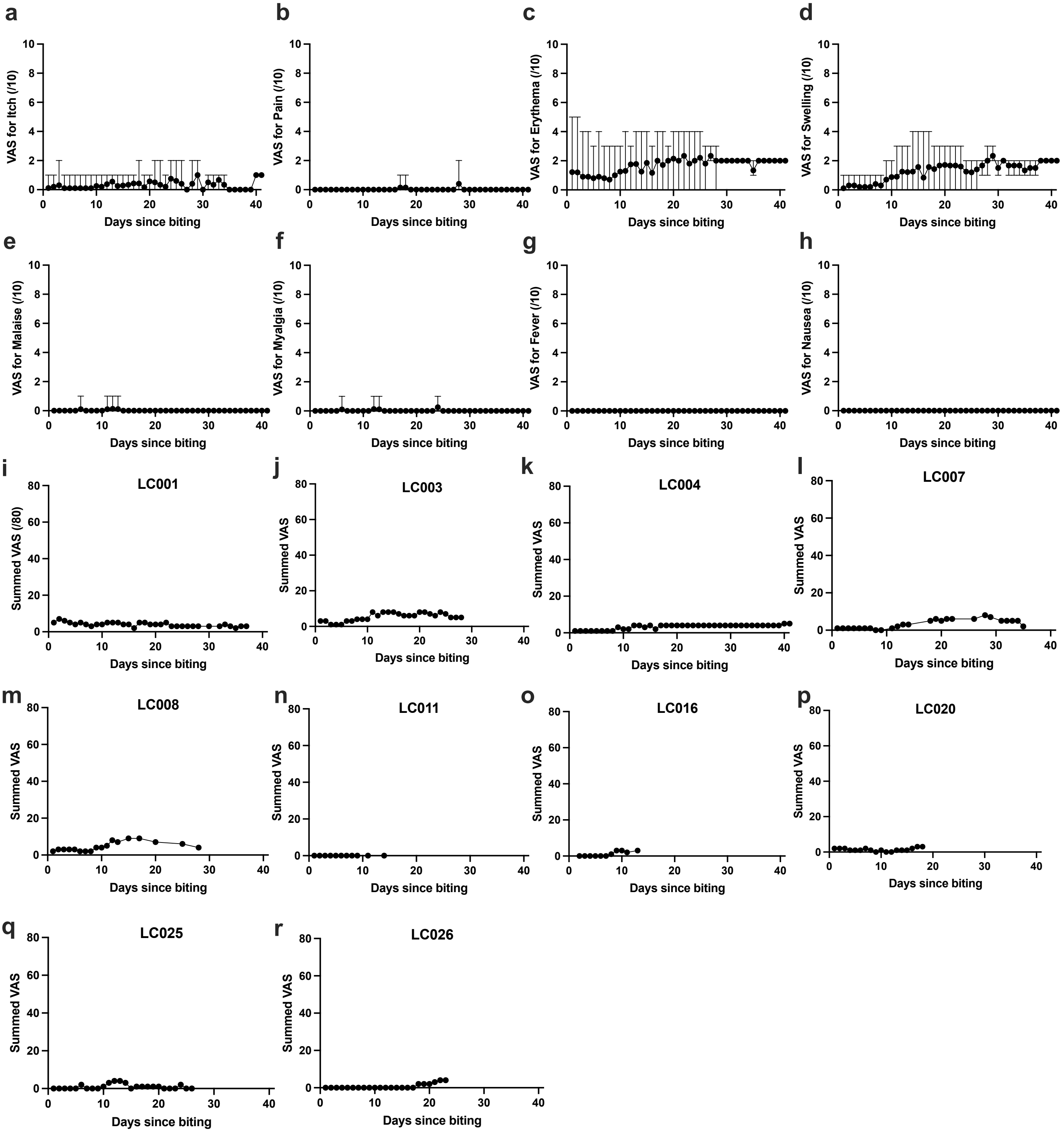


**Extended Data 6 Visual Analogue scales by feature and participant (relates to Figure 3)**

**a-h.** VAS scores out of 10 for itch, pain, erythema, swelling, malaise, myalgia, fever and nausea. Data shown as mean and range for all partcipants with positive bite.
**i-s.** Summed VAS scores for each participant. Data shown as total score out of 80.
