## Extended Data 3 for "Safety, effectiveness, and skin immune response in a controlled human infection model of sand fly transmitted cutaneous leishmaniasis"

**Study Acronym: LEISH_Challenge**

### A clinical study to develop a controlled human infection model using *Leishmania major*-infected sand flies

**Protocol version:** 1.2

**Protocol date:** 26th April 2022

**Sponsor:** University of York

**ClinicalTrials.gov ID** NCT04512742

| Authorised by | Role | Date | Signature |
|---|---|---|---|
| Charles Lacey | Chief Investigator | | |
| Michael Barber | Sponsor Representative | | |

| | |
|---|---|
| Chief Investigator | Professor Charles Lacey<br>Hull York Medical School<br>University of York<br>York, YO10 5DD<br> |
| Principal Investigators | Professor Paul Kaye<br>Hull York Medical School<br>University of York<br>York, YO10 5DD<br><br><br>Professor Petr Volf<br>Charles University<br>Department of Parasitology<br>Prague<br>Vinicna 7, Prague 2, 128 44<br>Czech Republic<br><br><br>Professor Alison Layton<br>Hull York Medical School<br>University of York<br>York, YO10 5DD<br><br><br>Prof Charles L. Jaffe<br>National Centre for Leishmaniasis<br>IMRIC, P.O. Box 12272<br>Hebrew University-Hadassah Medical School<br>Jerusalem 9112102 Israel<br><br><br>Professor Eli Schwartz<br>The Center for Geographic Medicine and Tropical Diseases<br>Department of Medicine<br>The Chaim Sheba Medical Center<br>Tel Hashomer 52621, Israel<br><br><br>Dr Vivak Parkash<br>Hull York Medical School<br>University of York<br>York, YO10 5DD<br>&<br>Department of Infection and Tropical Medicine<br>Sheffield Teaching Hospitals NHS Foundation Trust<br>Sheffield, S10 2RF |

| | |
|---|---|
| |<br><br>Professor Georgina Jones<br>School of Social Sciences<br>Leeds Beckett University<br>Calverly Building<br>Leeds, LS1 9HE<br> |
| Statistician | Prof Victoria Allgar<br>Peninsula Medical School<br>University of Plymouth<br>Plymouth, PL4 8AA<br> |
| Entomologist (York) | Dr Helen Ashwin,<br>Hull York Medical School/Department of Biology<br>University of York<br>York, YO10 5DD<br> |
| Entomologist (York) | Dr Katrien Van Bocxlaer<br>Hull York Medical School/Department of Biology<br>University of York<br>York, YO10 5DD<br> |
| Entomologist (Prague) | Dr Jovana Sadlova<br>Charles University<br>Department of Parasitology<br>Prague<br>Vinicna 7, Prague 2, 128 44<br>Czech Republic<br> |
| Project Management | Department of Biology<br>University of York<br>York, YO10 5DD |
| Laboratories | York Teaching Hospitals NHS Foundation Trust<br>Wigginton Road<br>York, YO31 8HE<br><br>Translational Research Facility,<br>Q Block, HYMS / Department of Biology,<br>University of York<br>York, YO10 5DD |
| Study Site | Translational Research Facility,<br>Q Block, HYMS / Department of Biology<br>University of York<br>York, YO10 5DD |

| | |
|---|---|
| Sponsoring Institution | University of York,<br>Heslington,<br>York, YO10 5DD |
| Sponsor Representative | Dr Michael Barber<br>Research and Enterprise Directorate<br>University of York,<br>Heslington,<br>York, YO10 5DD<br> |
| Funding | UK Medical Research Council,<br>Developmental Pathway Funding Scheme Award,<br>Development of a human challenge model of *Leishmania major* infection as a tool for assessing vaccines against leishmaniasis<br>Ref: MR/R014973/1 |
| Research Nurses and Clinical Trials Assistant (CTA) | Nursing staff and CTA are provided by York Teaching Hospitals NHS Foundation Trust |

Clinical Site and Participating Laboratories

Participant recruitment and study visits will take place at the Translational Research Facility, Department of Biology, University of York.

Haematological, biochemical & immunological screening and safety tests will be conducted at York Teaching Hospital NHS Foundation Trust and Department of Biology, University of York.

Definitions used in this protocol:

Chief Investigator (CI): Takes ultimate responsibility for the design, conduct, analysis and reporting of a clinical study or study.

Principal Investigators (PI): Directly assist the CI in delivery of the study objectives and may take the lead role in the event of temporary unavailability of the CI.

Co-Investigators (Co-I): Assist the CI in the specialist aspects of the study, including ensuring compliance with acceptable practice and applicable policies and regulations.

#### Protocol Signature Page

The signature below confirms agreement by the individual authorised by the Sponsors and persons responsible for signing the clinical study agreement that this study will be conducted in accordance with this protocol and GCP and ICH guidelines. Any amendments to this protocol that have a direct influence on the participants in the study will be approved by the relevant ethics committees before implementation.

I, the Chief Investigator, agree to allow sponsor monitor and auditors, full access to all medical records at the research facility for participants screened or enrolled in the study.

I agree to maintain all study documentation until the Sponsor consents to disposal of files in writing.

I have read and understood the information in the Investigator's Brochure including the potential risks and side effects of the investigational items and will ensure that all colleagues and employees assisting in the conduct of the study are informed about the obligations incurred by their involvement in the study.

| ______________________ | ______________________ | ______________________ |
| --- | --- | --- |
| Chief Investigator name | Chief Investigator signature | Date |

#### Confidentiality Statement

This document contains confidential information that must not be disclosed to anyone other than the Sponsor, the Investigator Team, and members of the Research Ethics Committee. This information cannot be used for any purpose other than the evaluation or conduct of the clinical investigation without the prior written consent of Professor Paul Kaye and Professor Charles Lacey.

#### Contents


| *Section* | *Title* | *Page Number* |
|---|---|---|
| | Abbreviations | 7 |
| 1 | Study Summary | 9 |
| 2 | Background and Rationale | 11 |
| 3 | Objectives | 18 |
| 4 | Study Design | 20 |
| 5 | Recruitment and Withdrawal of Study Participants | 25 |
| 6 | Procedures on Study Participants | 31 |
| 7 | Assessment of Safety | 46 |
| 8 | Management of Data, Samples and Study Procedures | 52 |
| 9 | Adaptive Design and Sample Size Considerations | 56 |
| 10 | Ethics | 57 |
| 11 | Regulatory and Governance Issues | 59 |
| 12 | Indemnity | 59 |
| 13 | Finance | 59 |
| 14 | Publication | 60 |
| 15 | References | 61 |
| Appendix 1 | Grading of clinical and laboratory adverse events | 64 |
| Appendix 2 | Skin excision biopsy SOP | 68 |
| Appendix 3 | Parasitological confirmation SOPs | 70 |
| Appendix 4 | Punch biopsy SOP | 72 |
| Appendix 5 | GAD-7 and DLQI questionnaires | 74 |
| Appendix 6 | Source data definition | 76 |


#### ABBREVIATIONS

| | |
|---|---|
| Ab | Antibody |

| | |
|---|---|
| ADL | Activities of Daily Living |
| AE | Adverse Event |
| ALT | Alanine Aminotransferase |
| AST | Aspartate Aminotransferase |
| BMI | Body Mass Index |
| CHIM | Controlled Human Infection Model |
| CHIMCAD | Controlled Human Infection Model Challenge Agent Dossier |
| CI | Chief Investigator |
| CL | Cutaneous Leishmaniasis |
| CMG | Clinical Management Group |
| CRF | Case Record Form |
| CRP | C-reactive protein |
| CTA | Clinical Trials Assistant |
| DAIDS | National Institutes of Health Division of AIDS |
| DNA | Deoxyribonucleic acid |
| ELISA | Enzyme-linked immunosorbent assay |
| FBC | Full Blood Count |
| GCP | Good Clinical Practice |
| GLP | Good Laboratory Practice |
| GMP | Good Manufacturing Practice |
| GP | General Practitioner |
| HbA1c | Haemoglobin A1c |
| HBsAg | Hepatitis B Surface Antigen |
| HBV | Hepatitis B virus |
| HCG | Human Chorionic Gonadotrophin |
| HCM | Human challenge model |
| HCV | Hepatitis C virus |
| HIV | Human Immunodeficiency Virus |
| HRA | Health Research Authority (UK) |
| HYMS | Hull York Medical School |
| ICH | International Conference on Harmonisation |
| IU | International Units |
| L | Litre |
| LFT | Liver Function Test |
| LLN | Lower Limit of Normal |
| LMICs | Lower- & Middle-income Countries |
| MHRA | Medicines and Healthcare Products Regulatory Agency |
| mg | milligram |
| Ml | Millilitre |
| MRC CTU | Medical Research Council Clinical Trials Unit |
| NHS | National Health Service |
| NIH | National Institute of Health |
| OTC | Over the counter |
| PCCL | Parasitologically confirmed cutaneous leishmaniasis |
| PPI | Patient Public Involvement group |
| PI | Principal Investigator |
| PKDL | Post-Kala Azar dermal leishmaniasis |
| R&D | Research and Development |

| | |
|---|---|
| RDT | Rapid Diagnostic Test |
| RN | Research Nurse |
| REC | Research Ethics Committee |
| SAE | Serious Adverse Event |
| SAP | Statistical Analysis Plan |
| SD | Standard Deviation |
| SMF | Study Master File |
| SMG | Study Management Group |
| SOC | Standard of Care treatment |
| SOP | Standard Operating Procedure |
| SSF | Study Site File |
| SUSAR | Suspected Unexpected Serious Adverse Reaction |
| TOPS | The Over-Volunteering Prevention System |
| TRF | Translational Research Facility |
| TSE | Transmissible spongiform encephalopathies |
| U&E | Urea and Electrolytes |
| UK | United Kingdom |
| ULN | Upper Limit of Normal |
| UoY | University of York |
| US | United States |
| VL | Visceral Leishmaniasis |
| WCC | White cell count |
| WHO | World Health Organisation |
| YTHFT | York Teaching Hospital Foundation Trust |
| µg | microgram |

#### 1.0 STUDY SUMMARY

| | |
|---|---|
| **Title** | A clinical study to develop a controlled human infection model using *Leishmania major*-infected sand flies |
| **Study Centres** | Translational Research Facility<br>Department of Biology<br>University of York<br>York, YO10 5DD, UK |
| **Study Identifier** | LEISH_Challenge |
| **Design** | Controlled human infection study, with an adaptive design |
| **Population** | Healthy subjects 18-50 years old; Male or Female |
| **Sample Size** | Up to 18 with confirmed sand fly bite (adaptive study design) |
| **Follow-up duration** | Up to 12 months |
| **Planned study Period** | October 2021 – October 2023 |
| **Primary Objective** | Development of a controlled human infection model of *Leishmania major* using sand fly transmission which is (a) effective and (b) safe |
| **Outcomes** | Effectiveness – development of parasitologically confirmed cutaneous leishmaniasis (PCCL) lesions<br>Safety – (i) lack of study-associated SAEs or grade 3 AEs, (ii) cutaneous leishmaniasis lesion clearance at 1 year follow up, using excision biopsy and if necessary additional cryotherapy |
| **Secondary Objectives** | 1. Determine rate of PCCL lesion development following infected sand fly bite<br>2. Determine response to *Leishmania major*-infected sand fly bite in terms of immunohistology and immunopathology<br>3. Determine parasite load in PCCL lesions in comparison to number of sand fly bites received and rate of lesion development<br>4. Determine acceptance and psychological impact of *Leishmania major*-infected sand fly challenge |
| **Outcomes** | 1. As determined by clinical examination, then biopsy and parasitological confirmation<br>2. Analysis of immune response and inflammation (for example macrophage and T-cell phenotype) and histology compatible with CL.<br>3. By quantitative PCR analysis of biopsy tissue of lesion site<br>4. Using psychometric questionnaires and focus groups |

| | |
|---|---|
| **Exploratory objectives** | In the case of a ≤66% take rate of *Leishmania major* with *Phlebotomus duboscqi*, to evaluate the take rate using *Phlebotomus papatasi*<br><br>To deep phenotype and compare CL and normal skin biopsies, in those who agree, using digital spatial profiling of host and parasite mRNA and protein expression, and mass spectroscopy imaging.<br><br>To determine human reactogenicity to *Leishmania major*-infected sand fly bite in macroscopic, dermoscopic, immunological and biochemical terms |

#### 2.0 BACKGROUND AND RATIONALE

##### 2.1 Background and unmet clinical need

The leishmaniases are global diseases.[1] They affect ~150 million people in 98 countries worldwide, are recognized by WHO as major neglected diseases of poverty, and disproportionately affect populations in low and middle-income countries (LMICs).[1,2] Visceral leishmaniasis (VL) results in more than 20,000 deaths annually; the cutaneous leishmaniases (CLs) impact quality of life for millions of people.[1] The leishmaniases are spread mainly by a group of insects called phlebotomine sand flies (Diptera: Psychodidae).

Although liposomal Amphotericin B has revolutionised VL treatment in South Asia, it is less effective elsewhere. Treatment for CL has changed little in the last 50 years. Drug resistance, the fragility of health systems and the limited impact of vector control represent significant challenges to controlling leishmaniasis with our current tools. The availability of an effective vaccine would have a major impact on health and economic development in LMICs where leishmaniasis is endemic. CL is also now not infrequently seen in the developed world, both in travellers returning from endemic areas and refugees.[2] However, no vaccine against human leishmaniasis has yet been successfully developed.[3][4] This development process would be immensely strengthened if there was a rapid, standardised method for the early evaluation of the efficacy of candidate vaccines.

Our proposed solution to this roadblock is the development of a Controlled Human Infection Model (CHIM) of sand fly transmitted *Leishmania major* infection.[4] Stage 1 of this project has comprised three components – a patient & public involvement group, the development of a *L. major* parasite bank at GMP, and an uninfected sand fly biting study. A focus group discussion at the end of the uninfected sand fly biting study has assessed acceptability and suggested modifications to this CHIM study. Stage 2 of the project is this study, which is to conduct controlled human infection in healthy participants. This study will establish effectiveness and safety, also provide data on a number of secondary objectives and will inform the subsequent development and use of the CHIM in leishmaniasis vaccine research. Our CHIM will therefore provide a new tool to assess vaccine efficacy which will subsequently allow evidence-based decisions to be made on down selection / progression of candidate vaccines against leishmaniasis.

##### 2.2 Leishmania parasite life cycle

The phlebotomine sand fly is integral to the transmission of *Leishmania.*[5][6] Female sand flies take a ‘blood meal’ from an infected host thereby becoming infected. Ingested amastigotes then transform into procyclic promastigotes which expand in numbers in the sand fly midgut. Accompanied by a series of developmental changes, parasites eventually reside as infectious metacyclic promastigotes in the anterior midgut near the stomodeal valve.[5][7]

The sand fly infects a further host by regurgitating these metacyclic promastigotes from the proboscis at the next biting event / blood meal. Promastigotes are then phagocytosed by macrophages and other phagocytes in the host where they transform into amastigotes (tissue stage). Multiplication of amastigotes then takes place within phagocytes, infecting other cells when the host cells burst.[6]

#### 2.3 Sand fly life cycle

Phlebotomine sand flies (Diptera: Psychodidae: Phlebotominae) are found in temperate climates where they bite hundreds of thousands of people every day. They are central to the life cycle and transmission of the *Leishmania* parasite. There are 4 distinct stages in their life cycle (egg, larvae, pupae, adult). An adult female may lay up to 70 eggs during a single cycle. Eggs are laid approximately 7 days after blood meal and hatch after a further 7 days. Larvae development takes 20-30 days. Adult sand flies emerge approximately 10 days later.(5)(6)

#### 2.4 Developing a *Leishmania* CHIM

Deliberate exposure of humans to infectious pathogens has been studied as a way to better understand disease and infection for several hundred years.(8,9) There has been much ethical debate regarding these studies given the deliberate nature of such infections.(10,11) Some of the very first deliberate human challenge infections involved smallpox, initially variolation and latterly and more famously Edward Jenner's work involving cowpox in the development of a vaccine against smallpox.

A similar approach has been employed for centuries with CL, referred to as leishmanization.(12,13) This method stems from the observation that after spontaneous resolution of Old World cutaneous leishmaniasis, individuals were nearly always protected against further infection. Therefore to protect against CL lesions arising on cosmetically important areas such as the face, individuals (often children) would be inoculated with live virulent parasites on the buttock, which would then be left to self-heal.(12)

In the last 60 years, a more structured, evidenced-based and ethical approach to deliberate infection has been employed. Scientists have sought to investigate diseases where a deliberate infection would be beneficial in either better understanding a disease or to test potential therapies(10), including diseases as diverse as influenza(14), norovirus(15), malaria(9) and dengue(16). This change in emphasis has been recognised through major funding from UK Government (e.g. the UKRI MRC/BBSRC-funded Hic-Vac network; https://www.hic-vac.org), and the Wellcome Trust (https://wellcome.ac.uk/grant-funding/schemes/human-infection-studies-vaccine-development), and by policy documents produced by the Academy of Medical Sciences (https://acmedsci.ac.uk/policy/policy-projects/controlled-human-infection-models).

The design of our study is influenced by data from *Khamesipour et al* (12). They provide clinical data on the rate of development of a CL ulcer after needle inoculation. 82.6% (19/23) of *Leishmania*-naïve volunteers developed an ulcerated lesion, and the earliest time point for ulcer development was 30 days (mean time to onset 66 +/- 23 days). Although we intend to intervene earlier than the ulcer development stage (see 6.3), as we are using sand fly transmission our assumption in terms of protocol design is that the time course of CL lesion evolution in our study may be of a similar order.

More recently, several methods have been used to try and emulate natural infection, including the use of a natural vector to transmit an infection, and with both *Plasmodium falciparum* and *Plasmodium vivax*, mosquito biting has been used to transmit malaria to humans.(9)

There has been significant research on the interplay between sand fly saliva and *Leishmania* parasites that promotes infectivity in pre-clinical models.[5] It has been shown that sand fly saliva is crucial for optimal infection. In addition, vaccines that protect mice against needle challenge may fail to do so after natural challenge[17] and therefore we believe it is important to use the natural sand fly vector to mimic the natural transmission process.

There is no clear indication in the medical literature to determine which of the major sand fly vectors of *Leishmania major* - *Phlebotomus papatasi* or *Phlebotomus duboscqi*, will be most effective at transmitting infection to a human host, this being governed by multiple biological processes. Both species have a similar mode of feeding and can support *L. major* development (see below). In our previous study, FLYBITE (see 2.5 below), we observed no significant difference in biting rates on humans (3.00±1.27 vs 3.3±0.81 flies fed from 5 applied, for *P. duboscqi* vs. *P. papatasi*, respectively; $p=0.56$). Nevertheless, based on pre-clinical data (see 2.8 below), *Phlebotomus duboscqi* was determined to be the lead candidate for use in this CHIM study.

#### 2.5 The FLYBITE study: A clinical study to develop a sand fly biting protocol using pathogen-free blood-fed sand flies

We have recently completed a clinical study to develop a sand fly biting protocol using pathogen-free blood-fed sand flies (the FLYBITE study). Twelve healthy participants were enrolled into the study and all 12 participants experienced at least one successful sand fly bite (Figure 1).

There were no SAEs or grade 3 AEs, and the biting methodology was successfully developed. The study participant feedback was extremely positive (see 2.6).

Figure 1 – CONSORT diagram of the FLYBITE study

| Participant number | Phlebtomine species | Number of sand flies fed |
|---|---|---|
| 1 | *P. duboscqi* | 2 |
| 2 | *P. duboscqi* | 4 |
| 3 | *P. papatasi* | 3 |
| 4 | *P. duboscqi* | 4 |
| 5 | *P. duboscqi* | 3 |
| 6 | *P. papatasi* | 4 |
| 7 | *P. papatasi* | 2 |
| 8 | *P. duboscqi* | 1 |
| 9 | *P. papatasi* | 4 |
| 10 | *P. papatasi* | 4 |
| 11 | *P. duboscqi* | 4 |
| 12 | *P. papatasi* | 3 |

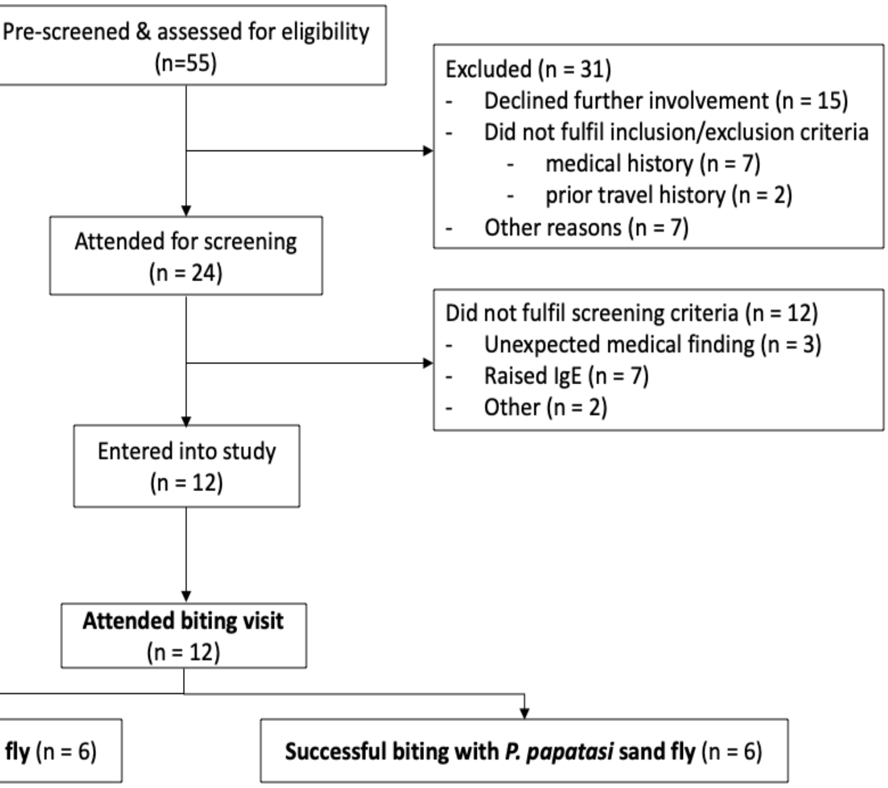


#### 2.6 Focus Groups and Patient and Public involvement

With a focus towards patient-centred healthcare, the discussion on stakeholder involvement in care has also moved to include research and clinical studies. The consensus is thus that high-quality research is dependent on patient and public involvement (PPI).[18] Therefore prior to the FLYBITE study, a public involvement consultation exercise with participants recruited widely was carried out to make clinical investigators aware of the public perception of human challenge models as well as to inform the study design.

A further consultation exercise took place in the form of a focus group with the FLYBITE study participants after completion of the study visits. Two separate focus groups took place shortly after the final FLYBITE study visits, with 5 participants in each lasting 3 hours. Participants were randomly assigned to each session. Georgina Jones (GJ) chaired and facilitated the group. Each session was divided in 2 halves, with the first being conducted by GJ with Liz Greenwood also present, however no clinical staff were present.
During the first session participants discussed experiences of taking part in the FLYBITE study, including recruitment, sand fly biting procedures, follow-up visits, communication and general administrative/logistical issues.
The second session was a discussion of the LEISH_Challenge study, and attitudes to *Leishmania* CHIM, at which all the study investigators were present.

Overall, the volunteers positively described their experiences in the FLYBITE study. In particular, they enjoyed the interaction with the study team, enjoyed the social aspect, had no safety concerns and felt appreciated, valued and well cared for throughout the whole study. The volunteer's experiences of being bitten was also positive. Generally, the individual flies were smaller than they had anticipated, and they also expected a higher number of sand flies to be placed in the biting chamber. Overall, the volunteers felt that taking part in the study had been 'unremarkable', with no surprises as everything had been explained to them beforehand. The physical effects of the bite were also perceived to be very minimal and generally were much less than they had expected prior to the start of the study. At no point did any of the volunteers want to exit the study. However, for some, the time commitment was greater than they had anticipated. Despite this, the volunteers expressed their gratitude to the team for being flexible and accommodating their competing time demands as much as possible during the duration of the study. Also, whilst they acknowledged the time commitment and would be willing to use other virtual follow-up methods e.g. uploading photos, they still felt the face-to-face contact was important and would wish this to continue.

##### 2.7 *Leishmania major* isolate MRC-02 as a challenge agent

An important consideration for selecting an infectious agent for use as a challenge agent is to ensure that its provenance can be fully defined. This safeguards for example, against the presence of other blood borne infections in the parasite donor and in vitro exposure to agents (e.g. bovine serum) able to cause transmissible spongiform encephalopathies (TSEs). As no such lines of *L. major* are available, parasites were freshly sourced from new clinical cases of CL. Parasites were obtained at time of diagnosis by ES under aseptic conditions at the Department of Medicine, The Chaim Sheba Medical Center, Israel. *L. major*_MRC-02 was obtained from a patient presenting with two ulcerated papules (~ 1.5 cm diameter) on their leg and a small nodule on their neck approximately five months after exposure to sand flies in the endemic area of the Negev. The patient elected for no treatment and there was complete healing 4 months later. At 18-month follow up, lesions remained healed with some scarring. The patient was seronegative for HIV, HBV, HCV and HTLV1.

*L. major*_MRC-02 was established at passage 1 at the Hebrew University by CJ, using GMP compatible culture media. Foetal calf serum was obtained from a TSE-free certified source (Thermofisher). Cryopreserved stocks were maintained in liquid nitrogen and samples shipped to York and Prague for further characterisation and to the Contract Development and Manufacturing Organisation (Vibalogics, Cuxhaven, Germany). Details of the characterisation and production of *L. major*_MRC-02 under GMP is described in our Controlled Human Infection Model Challenge Agent Dossier (CHIMCAD).

*L. major*_MRC-02 was genotyped by next generation sequencing under contract at Genome Quebec, using DNA prepared at York. DNA was also obtained from this line after passage in BALB/c. Data analysis was conducted by Dr Greg Matleshewski and Dr Patrick Lypaczewski at the University of McGill, Quebec, Canada. The data (i) confirm its identity as an *L. major* of likely Israeli origin, (ii) identified single nucleotide polymorphisms compared to the reference strain *L. major* Friedlin and (iii) confirmed that MRC-02 was genetically stable after a single passage in mice, based on both sequence and copy number variation (see CHIMCAD for further details).

##### 2.8 The two vectors *P. duboscqi & P. papatasi* and their prioritisation

Colonies of *P. duboscqi* and *P. papatasi* originating in Senegal and Turkey, respectively, were maintained in the insectary of the Department of Parasitology, Charles University in Prague, under standard conditions (26°C on 50 % sucrose, humidity in the insectary 60-70% and 14 h light/10 h dark photoperiod) as described previously (Volf and Volfova 2011). The sand fly colonies were screened by RT-PCR and found to be negative for phleboviruses (inc. Sandfly Fever Sicilian Virus group, Massilia virus and Toscana Virus) and Flaviviruses (targeting a conserved region of the NS5 gene).

*L. major*_MRC-02 log-phase promastigotes were used to experimentally infect *P. duboscqi* and *P. papatasi* using a membrane feeder. At days 3, 6 and 15 post feed, groups of sand flies were dissected to identify developmental stages of *Leishmania* and their position in the sand fly gut (endoperitrophic space, abdominal midgut, thoracic midgut, cardia and stomodeal valve). Parasite loads were estimated in situ (light, <100; moderate 100-1000; heavy >1000) and quantified after isolation fixation and counting. *Leishmania* with flagellar length ≤ 2x body length were scored as procyclic forms and >2x body length as metacyclic forms (see CHIMCAD for further details).

A series of studies were conducted using *L. major*_MRC-02 to compare infection rates, parasite developmental status over time, and onward transmission to BALB/c mice. These data are described fully in the CHIMCAD. Whilst both sand fly species effectively supported growth and development of *L.major* MRC-02 and transmission to mice, *P. duboscqi* showed a trend towards more rapid and consistent parasite development, and less mortality after feeding. As our purpose is to develop a CHIM with a high take rate, and as we will be carrying out curative interventions very early in the disease course, we have chosen to use *P. duboscqi* for the first six eligible and consented volunteers.

##### 2.9 The histological and parasitological diagnosis of CL

This is a CHIM study and the aetiological agent is therefore known. However, as we will be terminating infection at an early stage according to clinical criteria, formal confirmation of the CL diagnosis will be essential to exclude any other conditions mimicking the clinical appearance of CL. Examples of this could be on-going reactions to sand fly bites or bacterial infection at the bite site. Confirmation of the CL diagnosis will be through histological, molecular, and parasitological investigations carried out on the punch biopsy from the clinically suspected CL lesion. We are essentially following WHO guidelines[19], although we will use an improved method of detecting amastigotes in histology sections using RNAScope technology, or using an antibody for oligopeptidase B from *L.major* (OPB).[20] We will use the WHO case definition modified to our diagnostic protocols, in that a patient is considered to be a parasitologically confirmed CL (PCCL) case if at least one of the techniques – histology, PCR, or culture – is positive. Standard procedures will be used for skin biopsy (Appendix 2), histology, quantitative PCR and parasite culture (Appendix 3).

To maximise our understanding of the disease process and inform future vaccine development, biopsies will also be subject to deep phenotyping using a range of molecular immunology approaches, including digital spatial profiling of host and parasite mRNA and protein expression,[20] and mass spectroscopy imaging. Much greater knowledge can be gained and conclusions derived if these assays are performed on lesion tissue and a matched normal skin biopsy from the same subject. We will thus ask the study subjects if they are willing to have a second punch biopsy from normal skin on the contralateral forearm. This will be voluntary and the subjects can decline. Volunteers will be able to have the biopsy of healthy skin performed at a later visit.

##### 2.10 Study protocol for the treatment of the CHIM cutaneous leishmaniasis lesions

In normal clinical practice, CL lesions cause by *L. major* are regarded as benign. If they are small, not disfiguring, the patient is not immunosuppressed, and if the patient prefers the lesions are treated conservatively, i.e. left to heal spontaneously. There are different definitions of ‘small’, e.g. the WHO manual[(19)] defines this as up to 3 lesions, <4cm, and the LeishMan recommendations[(21)] defines this as up to 3 lesions, <3cm. No treatment is quite a common option in clinical practice, and our challenge strain, *L. major*_MRC-02, was derived from a patient that had 2 lesions, both <2cms, who was treated conservatively with healing at 9 months.

There are a number of other ‘relatively non-invasive’ therapies used for CL lesions caused by *L. major*, including cryotherapy, local injection with antimonials, heat treatments, or the use of topical Paromomycin ointment, which are recommended in the WHO and LeishMan guidelines.[(19,21)] A recent meta-analysis confirmed the efficacy of cryotherapy alone as a treatment for all types of CL. [(22)] An RCT of local treatments for CL due to *L. major* showed that cryotherapy alone was 100% effective for lesions ≤ 1 cm. [(23)]

In the setting of our CHIM study, leaving CL lesions to heal spontaneously is not a practical option for a number of reasons: (a) such prolonged healing as above, would make the study very lengthy, place undue commitments on both study participants and staff, increase the risk of study fatigue and participant default from follow up, and thus directly threaten the integrity of the study; (b) lesions could possibly progress, ulcerate, and undergo local spread, necessitating later treatment intervention, which would be more complex to administer, more risky for our study subjects, and with greater risk of scarring.

Therefore, we regard early intervention in the time course of any CL lesion development as an essential part of the methodology of our study. In dermatological practice, excision biopsy of lesions is used widely for both benign and neoplastic lesions. Excision biopsy of small CL lesions is not commonly used in routine tropical medicine practice, although it is mentioned as an option in some treatment reviews.[(24)] However, in the setting of a UK specialist tertiary referral centre, cases of CL lesions where excision has been used to establish the diagnosis are commonly seen, and these cases usually have good treatment outcomes and do not require additional treatment (see personal communication, Dr Steve Walker, Hospital for Tropical Diseases, London, UK).[25] Therefore in the setting of our study we believe excision biopsy of early lesions is the ideal immediate treatment approach, as it will ensure (a) cast iron confirmation that indeed a true CL lesion has developed at the bite site, rather than any other dermatological mimic, and (b) effective removal of all, or almost all, of the parasite load. We will therefore treat the early CL lesions in our study with excision biopsy, and then follow up the subject for 1 year. Full details are provided in Section 6.4.

#### 3.0 OBJECTIVES

##### 3.1 Primary Objective

The primary objective is the development of a controlled human infection model of *Leishmania major* using sand fly transmission which is (a) effective and (b) safe.

For the first six subjects we will expose them to successful biting (defined on p36, and further discussed in 6.9) by *Phlebotomus duboscqi* infected by *Leishmania major* and assess the 'take rate', that is the number of subjects developing parasitologically confirmed cutaneous leishmaniasis (PCCL) lesions. If either 6/6 or 5/6 subjects develop PCCL lesions then a further 6 subjects will be recruited; if only ≤ 4 subjects develop PCCL lesions, then we will follow the adaptive design described in Section 4.2, Figure 2.

**Outcomes**

Effectiveness will be assessed by -

The take rate of parasitologically confirmed cutaneous leishmaniasis lesions in study subjects

Safety will be measured by -

Assessing adverse event data collected through history, clinical examination & blood tests.

The development of any study-associated SAEs or grade 3 AEs at day 4 post-biting will result in a temporary halt and review of the sand fly biting schedule (see 4.2). Therefore, the Clinical Management Group (CMG) will review the safety outcomes 4 days after all biting procedures in real time for each pair of subjects (see 4.2).

Successful treatment of CL lesions in participants, and absence of lesions at 1 year follow up.

##### 3.2 Secondary Objectives

1. Determine rate of CL lesion development following infected sand fly bite
2. Determine response to *Leishmania major*-infected sand fly bite in terms of immunohistology and immunopathology
3. Determine parasite load in CL lesions in comparison to number of sand fly bites received and rate of lesion development
4. Determine acceptance and psychological impact of *Leishmania major*-infected sand fly challenge

**Outcomes**

1. As determined by clinical examination, then biopsy and parasitological confirmation
2. Analysis of immune and inflammatory response (for example macrophage and T cell phenotype) and histology compatible with CL.
3. By PCR analysis of biopsy tissue of lesion site
4. Using psychometric questionnaires and focus groups

**Exploratory objectives**

In the case of a ≤66% take rate of *Leishmania major* with *Phlebotomus duboscqi*, to evaluate the take rate using *Phlebotomus papatasi*.

To deep phenotype and compare CL and normal skin biopsies, in those who agree to donate such, using digital spatial profiling of host and parasite mRNA and protein expression, and mass spectroscopy imaging.

To determine human reactogenicity to *Leishmania major*-infected sand fly bite in macroscopic, dermoscopic, immunological and biochemical terms

#### 4.0 STUDY DESIGN

##### 4.1 Study Centre

The clinical study and clinical research will be carried out at the Translational Research Facility (Q Block), HYMS / Department of Biology, University of York, York, YO10 5DD.

Routine laboratory investigations will be carried out at York Teaching Hospitals NHS Foundation Trust, Wigginton Road, York.

The focus group(s) will take place at the University of York, York, YO10 5DD (or virtually if in-person meetings are not possible).

##### 4.2 Study Adaptive Design

This is a clinical study in up to 18 healthy *Leishmania*-naïve subjects (see inclusion criteria) aged between 18 and 50 years old who develop a confirmed sand fly bite (see 6.9 below). Initially we will study six subjects and expose them to biting by *Phlebotomus duboscqi* infected with *Leishmania major*. We will use an adaptive design, as described in Figure 2 below, that has been pragmatically designed to minimise unnecessary exposure of volunteers to *Leishmania* and maximise the likelihood of developing a reproducible CHIM.

Figure 2. Flow chart of the study adaptive design

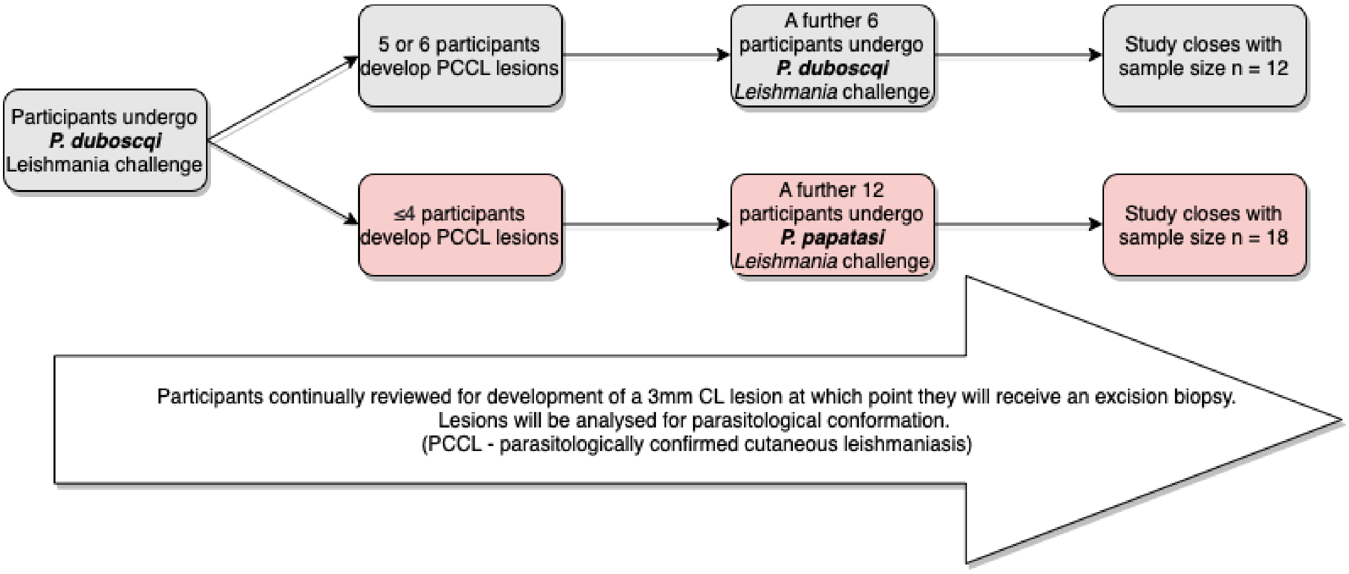


There will be ongoing clinical review of these participants to monitor the development of an early CL lesion. If either 6 or 5 participants have developed PCCL lesions within the 6-month follow up after *Leishmania* challenge, then a further 6 subjects will undergo *Leishmania* challenge by *P. dubosqi*. If only 4 or fewer subjects in the first cohort develop PCCL lesions, then we will switch vector to *P. papatasi* and a further 12 subjects will undergo *Leishmania* challenge.

The sand flies will be fed one blood meal containing parasites prior to the biting procedure. We will attempt to study up to two participants on each biting day, with one participant in the morning, and one in the afternoon.

Screening for the study will take place from up to 90 days to 7 days prior to sand fly exposure. All participants will then undergo the biting procedure with 5 infected sand flies in a single biting chamber, within which access is restricted. The aperture may be up to 6mm in diameter. Follow-up will take place as per Table 1, page 32.

##### 4.3 Sand flies and *Leishmania* parasites

The sand fly colonies are maintained at Charles University, Prague[(6)]. Sand flies will be transported to the Department of Biology, University of York in specialised containment from the Charles University, Prague. The sand fly colonies have been screened by RT-PCR and found to be negative for phleboviruses (inc. Sandfly Fever Sicilian Virus group, Massilia virus and Toscana Virus) and Flaviviruses (targeting a conserved region of the NS5 gene).

Colonies are maintained by Prof Petr Volf in Prague. Sand flies are transported to York by courier inside a sealed unit. Clay is placed in the bottom of the unit to help maintain humidity. The sand flies are shipped at the age of 3 to 5 days of adult development. Upon arrival in York of the sand flies, a designated member of staff will check shipping conditions, organise storage and acknowledge receipt to the supplier, according to our defined SOPs.

After arrival the non-infected sand flies are kept within a secure insectary at the University of York. The room is under negative pressure and is situated within a secure unit. The containment room has an air curtain at the entrance to prevent escape of sand flies and also contains insect electrocutors. Only trained members of staff with health clearance and appropriate experience are allowed access to the sand fly facility.

The sand flies are kept in a dedicated incubator (26ºC, 70% humidity, photoperiod of 12-14 hours light and 10-12 hours dark adding up to a total of 24 hours). They are housed within an insect cage with a feeding membrane (BugDorm, MegaView Science Co., Ltd., Taiwan). Sand flies are maintained on a sugar solution between feeding (soaked cotton wool, 50% sugar solution).

Once the sand flies have arrived at the Department of Biology, University of York, a cotton-soaked sugar solution (50%) is placed within the cage for 24 hours, from which the sand flies can feed. The sugar solution is then removed and the sand flies are starved. Twelve to fifteen days prior to a scheduled biting day, sand flies will be infected with *L. major* using a membrane feeder (Hemotek) containing rabbit blood mixed with $10^6$ / ml promastigotes of *L. major*_MRC-02. Female sand flies prefer to feed in the dark, hence the insect cage is covered with an opaque material during this process. Three to five days before a scheduled biting day, a subset of the engorged sand flies will be killed and dissected to evaluate infection rates by standard methods (see CHIMCAD for further details). If infection rate is below 90% or parasites have failed to develop to infectious forms, the study will be postponed.

On the day of the biting study, the biting chambers will be loaded with infectious sand flies within the secure insectary. A biting chamber (Figure 3) is placed on ice. Sand fly containers are cooled with surrounding ice (as cold temperature reduces the metabolism of the sand flies and immobilizes them) allowing ease of handling. Five female previously engorged female sand flies will be added to the biting chamber using tweezers. In the unlikely event that sand flies are not available on the day of the biting study, the study will be postponed until the sand flies are available.

A record will be kept of each lot of sand flies used during the study. This will include the participants' study number, the description (lot numbers) of sand flies received at study site and date of receipt, as well as a record of when (date of biting challenge) and whom (volunteer number) underwent challenge.

##### 4.4 Study Volunteers and biting procedure

Up to eighteen healthy participants aged between 18-50 years will be recruited. All subjects will be willing and able to adhere to the study conditions, methodology and to give written informed consent. For more details refer to section 5.0.

Participants will undergo the biting procedure with 5 sand flies over the course of 30 minutes. Only one study participant will undergo sand fly biting at any one time (see section 4.6).

**Biting chamber**

Figure 3. Sand fly biting chamber

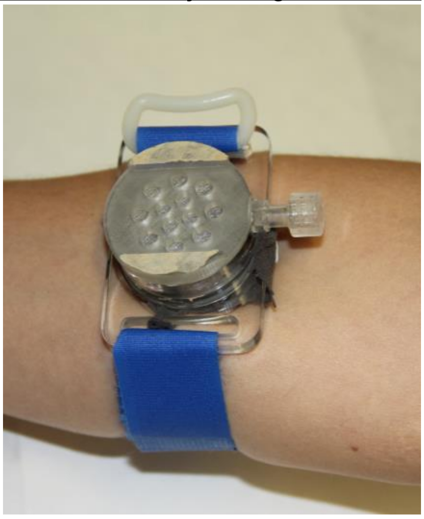

We will use a custom-made sand fly biting chamber with straps, manufactured by Precision Plastics Inc, Maryland, USA. This is a watch-like apparatus with an adjustable Velcro strap. Once the unit is sealed with sand flies inside, they are unable to escape from the sand fly biting chamber (Figure 3). Sand flies are loaded into these biting chambers in the secure insectary.

**Transport of sand flies for human biting studies**

The sand flies will be transported sealed within the biting chamber and a secondary container inside a tertiary container to the Translational Research Facility (Q Block). After the biting study, the biting chamber is transported back to the insectary in the same way for subsequent disposal. The containers are clear plastic and therefore any containment breach will be easily identifiable from the tertiary container. Glass will be avoided to prevent breakages and therefore containment breaches.

**Biting procedure**

On the day of sand fly biting, the biting chambers with the sand flies enclosed within the containers (described above) will be taken to the clinical suite within the Translational Research Facility (Q Block), Department of Biology, where the study visits will take place.

The preferred placement for each biting chamber will be on the volar aspect of the proximal forearm, approximately 2-3 centimetres distal to the antecubital fossa. Discussion within the public involvement consultation exercise as well as the focus groups suggested an alternative option of the upper arm below the deltoid region should also be given. Participants will be asked which arm they prefer (non-dominant limb will be suggested).

The sand fly biting chambers will be secured on the participants arm for 30 minutes after which it will be removed. The participant will be observed both during the 30-minute biting period and for 2 hours after for signs of any localised and/or generalized reaction. A medical team will be in the building at all times. This will consist of at least 1 doctor and 1 nurse. In the unlikely event of a nurse not being available (e.g. due to NHS surge pressures) an additional medically qualified staff member (i.e. doctor) will be acceptable cover.

Appropriate resuscitation equipment (including defibrillator, medical grade oxygen and resuscitation drugs) will be available in the unlikely event it is needed. Clinical and nursing staff will be appropriately trained to use the resuscitation equipment and will have appropriate advanced life support skill qualifications.

Non-clinical personnel will not be allowed within the clinic areas during the participant observation unless express permission is given by study investigators, with the exception of the entomologist. The participants will be asked to self-report any bite sensation experienced whilst wearing the biting chamber. This will be timed and recorded.

Although anaphylaxis is not anticipated based on previous biting studies and a review of the literature, participants will be observed for evidence of anaphylaxis, including but not limited to skin rashes, itching and hives, swelling of the lips, tongue and throat, shortness of breath, or wheezing, fainting, vomiting or diarrhoea and tachycardia (rapid pulse rate). Intramuscular adrenaline (1:1000), and other drugs according to the latest guidelines will be available for administration should anaphylaxis occur, and the participant would then be transferred to the York Teaching Hospital NHS Foundation Trust via ambulance. During the FLYBITE study there were no cases of anaphylaxis or anaphylactoid reaction affecting any of the 12 participants recruited to the biting study.

Localised skin reactions including redness, scaling and swelling may be treated with oral antihistamines (such as cetirizine, chlorphenamine) for pruritus (itch) or any persistent reaction. Any pain or discomfort would be treated with oral paracetamol and/or ibuprofen. The biting site will be covered with a light dressing if necessary. These medications will be available from the Translational Research Facility after being prescribed and dispensed by a medical practitioner on the study team.

##### 4.5 Safety review within protocol

Only 1-2 participants will undergo sand fly biting on any given day, and no participants will undergo sand fly biting simultaneously. Participants will be scheduled to attend for further follow-up post-biting visit (see section 6). The development of any SAE or non-solicited grade 3 or greater AE at the follow-up visits, which are considered possibly, probably or definitely related to the biting procedure (see 7.2) will result in a temporary halt and review of the sand fly biting parameters. Therefore, we will review the safety outcomes 4 days after all biting procedures in real time for all subjects. If an SAE or non-solicited grade 3 AE had been recorded the CI (CL) and PIs (VP, AL & PK) would review both the clinical event, and the biting parameters in terms of the number of sand flies, the species and length of exposure, with regard to progression of the study. If non-solicited Grade 3 / exaggerated reactions had been observed it would be likely that the Investigators (as above) would seek a protocol amendment to decrease the number of sand flies present within the biting chamber, or length of exposure to mitigate any exaggerated response. At the discretion of the clinical investigators/CMG (CL, VP, AL) it would be possible that a sufficient number of grade 2 AEs, would also result in such a halt to the study and consideration of a protocol amendment. As described in the results of the FLYBITE study (section 2.5), 12 participants underwent sand fly biting without development of grade 3 AEs or SUSARs, therefore we do not anticipate any significant reaction in this study. As described earlier in this protocol, we are unable to locate via literature searching any proven cases of anaphylaxis to the sand fly species used in this study.

(For discussion of personal protective equipment and COVID-associated risk mitigation, see section **6.12 Safety of staff and participants during visits)**

#### 4.6 Duration of Study

All participants will undergo a pre-screening assessment. The follow up visits are as described in section 6. After participants have completed their treatment and follow-up, focus groups will be held (see section 6.5 for details).

#### 4.7 Definition of the Start and End of the Study

The start of the study is defined as the date of the screening visit of the first participant. The end of the study is defined as 3 months after the date of the final focus group.

#### 4.8 Study Procedures

All study procedures will be carried out according to study SOPs.

#### 4.9 Non-study treatment

Participants must not have had any immunisation with any vaccine up to 21 days prior to their screening assessment. Participants must also agree to avoid all immunisations in the 21 days, both before and following the *Leishmania* challenge procedure.

All concomitant medication will be recorded in the CRF including any dispensed by a clinical investigator in the management of adverse events.

#### 4.10 Study treatment

Participants will be provided with an oral OTC anti-histamine medication (eg Cetrizine) following the biting study, to be used symptomatically if necessary for any significant localised reaction e.g. itch. Other simple OTC treatments may also be provided (e.g. paracetamol). See section 6.4.2 for further discussion about treatment.

#### 5.0 RECRUITMENT AND WITHDRAWAL OF STUDY PARTICIPANTS

##### 5.1 Number and Source of Participants

We will recruit up to eighteen healthy participants via advertisement both within and outside the university. Publications, and both internal and external websites will be used as well as relevant mailing lists. This will include posters and a university-wide email newsletter.

##### 5.2 Inclusion and Exclusion Criteria

This study will be conducted in participants who meet the following inclusion and exclusion criteria.

###### Inclusion Criteria

The participant must be:

- Healthy adults aged 18 to 50 years on the day of screening
- Willing to give consent for exposure to *Leishmania*-infected sand fly with the intention of causing a cutaneous leishmaniasis lesion
- Willing and able to give written informed consent
- Willing to undergo Hepatitis B, Hepatitis C & HIV testing
- Willing to undergo a pregnancy test during screening and follow-up visits and must not be breastfeeding (pre-menopausal female participants)
- Willing to refrain from blood donation during the study
- Using a reliable and effective form of contraception (pre-menopausal female participants up to 3 months post-biopsy
- Judged, in the opinion of a medically qualified Clinical Investigator, to be able and likely to comply with all study requirements as set out in the protocol
- Without any other significant health problems as determined by medical history, physical examination, results of screening tests and the clinical judgment of a medically qualified Clinical Investigator
- Available for the duration of the study
- Willing to refrain from travel to regions where *Leishmania*-transmitting sand flies are present, from recruitment until an appropriate point (judged by study investigators).
- Willing to consent to a copy of the past medical history to be provided by the participant's GP practice.
- Agree to registration on a national database of study & trial subjects to prevent over-volunteering (TOPS)
- Willing to give consent for study investigators to contact the participants GP in the event of a significant abnormality being observed
- Willing to show identification documents to confirm identity
- Willing to give consent to biopsy(s) of suspected cutaneous leishmaniasis lesions

###### Exclusion Criteria

The participant may not enter the study if any of the following apply:

- Receipt of any vaccine within 21 days of screening
- Administration of immunoglobulins and/or any blood products within the three months preceding the planned study.

- History of significant allergic disease/atopy (e.g. significant/severe eczema, hay fever, asthma) or reactions; or a history of severe or multiple allergies to drugs or pharmaceutical agents, as judged by the clinical investigators
- Any significant chronic skin condition as judged by the clinical investigators
- Any history of confirmed Leishmaniasis infection
- Any history of travel (within the 30 days prior to the *Leishmania*-infected biting visit) to regions where *Leishmania major*-transmitting sand flies are endemic*.
- Any history of more than 30 continuous days stay in regions where *Leishmania major*-transmitting sand flies are endemic within the last 10 years*.
- Any history of severe local or general reaction to insect bites, defined as
  - Local: extensive, indurated redness and swelling involving most of the antero-lateral thigh or the major circumference of the arm, not resolving within 72 hours
  - General: fever ≥ 39.5°C, anaphylaxis, bronchospasm, laryngeal oedema, collapse, convulsions or encephalopathy within 48 hours
- Any history of anaphylaxis
- Females – current pregnancy, less than 12 weeks postpartum, lactating or willingness/intention to become pregnant during the study.
- Any clinically significant abnormal finding on screening biochemistry or haematology blood tests as judged by study investigators
- Total IgE levels > 214 IU/ml
- Any confirmed or suspected immunosuppressive or immunodeficient state, including HIV infection; asplenia; recurrent, severe infections and chronic (more than 14 days) immunosuppressant medication within the past 6 months
- A diagnosis of diabetes type 1 or type 2 or significantly raised HbA1c
- Active Tuberculosis, leprosy, or malnutrition
- Any significant chronic illness requiring hospital specialist input as judged by study investigators
- Any significant psychiatric conditions as judged by general practitioner and/or study clinical team
- Unlikely to comply with the study protocol
- Participating in significant current or recent research (involving an investigational medicinal product or other significant intervention) within the past 3 months (as judged by study investigators)
- Any other significant disease, disorder, finding or medical history, which, in the opinion of a medically qualified Clinical Investigator, may either put the participant at risk because of participation in the study, or may influence the result of the study, or the participant's ability to participate in the study
- Any known risk factors for Creutzfeldt Jakob Disease (CJD) or variant CJD

*This refers to regions where *Leishmania major*-transmitting sand flies are endemic including (but not limited to) the Middle East, Sub-Saharan Africa, and Asia. (see Figure 4, World Health Organisation Map)

##### 5.3 Screening Procedures and Investigations

The following general eligibility criteria will be assessed prior to conducting informed consent during a pre-screen:

- Age

- Availability for the duration of the study (including follow-up)
- General health state (including history of atopy i.e. asthma / hay fever / eczema)
- Allergy status (medications and other allergens)
- Risk of previous *Leishmania* and phlebotomine sand fly exposure as determined by travel history
- Contact details

This pre-screen assessment may take place face-to-face, via phone or via email.
Following the pre-screening assessment, the participant will be given written material and allowed to consider their recruitment into the study. A discussion of the potential risks and obligations will also be highlighted. If they indicate a willingness to participate, they will be assigned a date and time for a formal Screening Visit. It is possible that a screening visit can take place concurrently if the participant has had enough time to read the recruitment material and make an informed decision based on these materials.

The Chief Investigator (or a study physician in accordance with the delegation log) has both an ethical and a legal responsibility to ensure that each participant being considered for inclusion in the study is given a full explanation of the study. If the participant is still willing and interested, they will be asked to sign and date a copy of the Consent Form. A copy will be given to the participant to keep, a copy is to be stored in the participant's case file and the original is to be placed in the Study Site File.

To ensure informed consent, the following information will be provided by a member of the study team at screening
- Pre-HIV test discussion
- The importance of continued follow up in the study to monitor any unforeseen events will be stressed

After informed consent has been obtained, assessments and investigations will be undertaken according to the schedule. These include -
- Medical history,
- General physical examination
- Confirmation of identification and photography
- If female, a pregnancy test
- Blood samples for routine laboratory investigations (haematology and biochemistry) and the blood borne viruses Hepatitis B, Hepatitis C and HIV.
- Blood sample for *Leishmania* antibody test and immune monitoring
- A standard letter will be sent to the GP outlining the study. The GP practice will be asked to provide a summary of the medical history.

A record of the above procedures and any findings will be entered into the Case Report Form (CRF).

A pre-screening questionnaire will be used to identify eligibility at an early stage. If there are any screening failures at this stage, the collected data will be retained unless the participant withdraws consent for this.

Each participant who enters the study by signing a copy of the consent form will be assigned an Identification Number. These numbers will not be reassigned. A screening log of screened participants with their identification number and basic details will be maintained and kept in the study site file to track participants who have been screened for the study.

Photographic records of the sand fly bite site pre- and post-study will also form part of the clinical file.

Following the screening visit a medically qualified Clinical Investigator will review the results of each participant to determine if they are eligible for further involvement in the study. The Chief Investigator (or a medically qualified clinical investigator in accordance with the delegation log) must confirm eligibility for each participant based on the screening assessment. This must be documented in the CRF. If the participant is not enrolled into the biting portion of the study after being deemed ineligible, all screening data and samples collected up to that point will remain available for analysis as part of the study unless the participant withdraws consent for this. If there is deemed to be any information relevant for the participant's GP to be made aware at this stage, consent will be taken to write to the GP. Consent for GP communication will be taken at the initial screening visit.

Participants may be screened up to 90 days before the biting day visit. If this 90 day period has elapsed, participants may still be included in the study if there have been no significant changes to medical history or circumstances, and a fresh set of baseline blood tests has been obtained. If there has been a significant change in circumstances, then participants can take part in the study only after a full re-screen has taken place. The original study ID number will be unaltered in these circumstances.

##### 5.4 Eligibility and Screening failures

A medically qualified Clinical Investigator must confirm eligibility for each participant based on the screening procedures, including findings from clinical histories, examinations, laboratory results. This must be documented in the CRF. If the participant is deemed to be eligible, further participation can then be taken.

If for any reason the participant is considered a screen failure the participant will be informed by a Clinical Investigator, notified of all of their results and the reason for the screen failure.

Eligibility will be confirmed by 2 members of the clinical team prior to enrolment of participants into the challenge phase of the study.

##### 5.5 Re-Screen

If the investigator believes that one or more of the laboratory screening test results are an anomaly and temporary (as described in MHRA Good Clinical Practice guidelines; 1st Edition 2012, Section 11.4.3 - page 377), then a repeat of the relevant tests may be carried out. The reason for carrying out a repeat testing must be clearly documented in the CRF and subjects may only undergo repeat testing to confirm or dispute anomalies. If, following repeat testing, any laboratory parameter falls outside of the inclusion range for the study then the subject will be excluded from the study and no further re-screening should take place. If an anomaly has been reported as a result of lab error, then this will not count as a repeat test.

#### 5.6 Pregnancy

If a participant is found to have a positive pregnancy test at the screening visit, they will be considered a screen failure. The participant will be given the result by a Clinical Investigator, and a report sent to their GP. They will still be able to take part if they expressly wish for this at a later date, but only if they attend for repeat screening (e.g. false positive pregnancy test or after termination of pregnancy).

Pregnancy testing will be performed by use of serum β-human chorionic gonadotropin (β-HCG) result at the screening visit. Urinary pregnancy testing (urinary β-human chorionic gonadotropin) will only be used if a suspicion of pregnancy arises during the study. . The following serum β-HCG values will prompt further investigation:

- Females pre-menopause: ≥ 1 IU/L (may suggest pregnancy)
- Females post-menopause: ≥ 7 IU/L (may suggest other pathology that will need follow-up)

These are the correct values at the time of writing but may be altered if the reference ranges are revised by the testing laboratory (York Teaching Hospitals NHS Foundation Trust).

If a female participant becomes pregnant following infected sand fly bite, she will be followed up as other participants. The pregnancy will be reported to the Sponsor in accordance with the SOP for Research Related Adverse Event Reporting Procedure, and a report sent to their GP. (see also Section 7.6)

#### 5.7 Procedure for follow-up of adverse events and pregnancy

The Clinical Investigators will make every effort to monitor all adverse events, regardless of severity, until resolution or stabilisation, in order to report this on the CRF during the study.

After the database is locked and the study is closed, any additional information about adverse events or pregnancy that comes to the attention of a Clinical Investigator will be reported by email to the Chief Investigator and the Sponsor.

#### 5.8 Baseline and continuous psychological assessment

Psychological well-being will be evaluated at baseline and at regular intervals during selected visits (see schedule). This will be undertaken to determine acceptability of the study and its procedures to the participant, and whether participation is associated with any undue psychological outcomes. Two instruments will be used, the GAD-7 psychometric questionnaire / quality of life measure[26] and the DLQI (Dermatology Life Quality Instrument).[27] These questionnaires may be filled remotely and returned electronically at the discretion of the study investigators.

The GAD-7 is a brief 7 item generalised anxiety measure with normative data available. It has good thresholds and cut-off values and will be able to capture the key generalised anxiety components that we identified as possible issues in the FLYBITE study. It is also not overly burdensome. The DLQI is a validated generic dermatology QOL questionnaire and gives more capacity to collect other patient reported psychosocial dermatological symptoms.

##### 5.9 Regions of endemicity of *Leishmania major*-transmitting sand flies

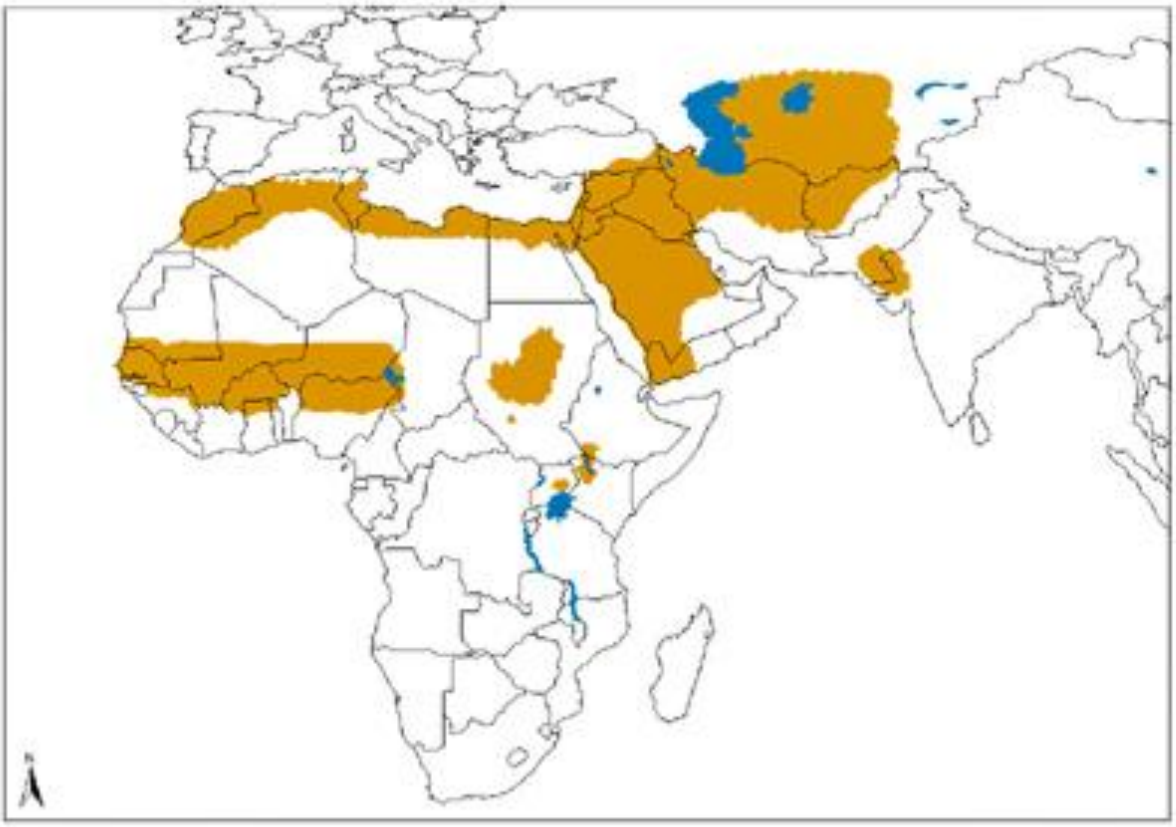

Figure 4.Geographical distribution of Old World cutaneous leishmaniasis due to *L. major* (https://www.who.int/leishmaniasis/leishmaniasis_maps/en/index1.html)

Regions where *Leishmania major*-transmitting sand flies are endemic including (but not limited to) the Middle East, parts of North and Sub-Saharan Africa, and Asia. The affected countries are listed below, although not all regions of a given country may be affected by *Leishmania major* (including, but not limited to):

Morocco, Algeria, Tunisia, Libya, Egypt, Mauritania, Senegal, The Gambia, Guinea-Bissau, Guinea, Mali, Burkina Faso, Ghana, Togo, Benin, Nigeria, Niger, Chad, Cameroon, Sudan, South Sudan, Ethiopia, Kenya, Uganda, Yemen, Saudi Arabia, Jordan, Israel, Lebanon, Iraq, Syria, Turkey, Iran, Pakistan, India, Afghanistan, Tajikistan, Kyrgyzstan, Uzbekistan, Kazakhstan, Turkmenistan, Gaza, West Bank

#### 6.0 PROCEDURES ON STUDY PARTICIPANTS

##### 6.1 Study procedures

Procedures will be performed on the visit time points indicated in the schedule of procedures. Additional procedures or laboratory tests may be performed, at the discretion of a Clinical Investigator if clinically required.

**Observations** which will be documented include blood pressure, pulse rate, temperature, oxygen saturations and respiratory rate. Height and weight will also be recorded.

**Blood tests** will be drawn for the following laboratory tests at visits as per the schedules:

- Haematology: Haemoglobin, White blood cells, Neutrophils, Lymphocytes, Platelets
- Biochemistry: Renal function, liver function, β-Human Chorionic Gonadotropin, C – reactive protein, HbA1C
- Blood Borne Virus Screen: Hepatitis B surface antigen, HIV antibodies, Hepatitis C antibodies
- Immunology: Total IgE,CD4 & CD8 subsets; immune monitoring
- Cellular responses: peripheral blood mononuclear cells isolation
- β-human chorionic gonadotrophic: in women at screening
- *Leishmania* & sand fly testing: Serology and other confirmatory tests to determine presence of *Leishmania* and sand fly-related factors

**Urine** will only be collected for dipstick testing for diagnosis and management of intercurrent medical presentations during follow up, e,g. systemic illness, UTI, suspicion of pregnancy, etc:

**General examination** which will be performed will include cardiovascular, respiratory, neurological, abdominal, inspection of proposed bite site and palpation of axillary and cervical lymph nodes. A general examination of the skin to exclude relevant inflammatory dermatoses will be undertaken.

Haematology, biochemistry, blood-borne virus testing and IgE will be performed at the York Teaching Hospitals NHS Trust. Blood taken for these tests will be transferred usually within 24 hours to the hospital laboratory. In exceptional circumstances, samples may be temporarily stored at the University of York prior to transfer to the York Teaching Hospitals NHS Trust for testing. All other tests will be performed within the Department of Biology and stored according to standard local protocols.

**Analysis of biopsy samples:** including immunohistology, immunopathology, parasite culture and PCR. Analysis of the immune/inflammatory response will also be carried out. Skin samples of suspected CL lesions will be compared to optional healthy skin samples.

##### 6.2 Schedule of Visits

The schedules of visits post-challenge and pre-biopsy are presented below in Table 1, Parts 1 & 2. Participants will be monitored and followed up in the Translational Research Facility (TRF).

**Table 1, Part 1: Schedule of Visits, Challenge Phase, out to 28 days post-challenge follow up**

| | **Pre-screening**<br>Days -90 to -14 | **Visit 1**<br>Screening<br>Days -90 to -7 | **Visit 2**<br>Sand fly biting<br>Day 0 | **Visit 3**<br>FU<br>Day 4 | **Visit 4**<br>FU<br>Day 14 | **Visit 5**<br>FU<br>Day 28 |
|---|---|---|---|---|---|---|
| Window | N/A | N/A | N/A | ± 2 day | ± 4 days | ± 5 days |
| Information & discussion with the participant | X | X | | | | |
| Consent | | X | X | | | |
| History, Examination, Image documentation | | X | X | X | X | X |
| Potential clinical diagnosis of CL, and excision biopsy | | | | | | X |
| FBC, U+E, LFT, CRP, | | X | | | | X |
| HbA1c | | X | | | | |
| Total IgE, CD4 / CD8 | | X | | | | |
| Blood borne virus screen (HIV, Hepatitis B/C) | | X | | | | |
| *Leishmania* serology | | X | | | | X |
| Peripheral blood mononuclear cells & immune monitoring | | X | | | X | X |
| Serum β-HCG pregnancy test (females) | | X | | | | |
| Quality of Life questionnaires | | X | | | X | X |
| Blood volume | | 60ml | 0ml | 0ml | 30ml | 50ml |

**Table 1, Part 2: Schedule of Visits, Challenge Phase, from day 42 to day 154 post-challenge follow up, pre-biopsy**

| | **Visit 6*** Day 42 | **Visit 7** Day 56 | **Visit 8*** Day 70 | **Visit 9** Day 98 | **Visit 10*** Day 126 | **Visit 11** Day 154 |
|---|---|---|---|---|---|---|
| Window | +/- 7 days | +/-7 days | ± 14 days | ± 14 days | ± 14 days | ± 14 days |
| History, Examination, Image documentation | X | X | X | X | X | X |
| Potential clinical diagnosis of CL & excision biopsy | X | X | X | X | X | X |
| FBC, U+E, LFT, CRP | | X | | X | | X |
| *Leishmania* serology | | X | | X | | X |
| Peripheral blood mononuclear cells & immune monitoring | | X | | X | | X |
| Quality of Life questionnaires | | X | | X | | X |
| Blood volume | 0ml | 50ml | 0ml | 50ml | 0ml | 50ml |

*Scheduled as remote visits by video calling

Any diagnosis of an intercurrent illness during the study, which is not related to the study or its procedures, will either be referred to the subject's GP, or if necessary, the Emergency Department at York Hospital.

**Visit 1: Pre-screening, screening and Enrolment**

All potential participants will have a pre-screening assessment conducted either face-to-face, via email or via telephone to determine eligibility and availability. Their eligibility will be assessed by one of the investigators, and if they fulfil basic criteria for progression to screening, the study will be discussed with them, and information leaflets provided.

All potential participants will then have a screening assessment, which will take place between 90 & 7 days prior to the sand fly biting visit. If they are suitable and agree to participate in the study then informed consent will be undertaken. At the screening visit the subjects will undergo a full history, examination and blood testing as detailed in the case record form (CRF). Following this visit a provisional date and time for the sand fly biting visit will be agreed. Once the results of all the screening tests and the past medical history (provided by the participants GP practice) have been received the CI will confirm eligibility in the CRF and the date and time of the biting visit confirmed with the participant. A neutral scent-free body wash will be provided for use on the biting day.

During the screening visit the participants will also be asked to provide their National Insurance or passport number so that this can be entered on to a national database which helps prevent participants from over-volunteering for clinical studies and trials (www.tops.org.uk).

A photograph will be taken for identification purposes and which will be placed in the participants file only. Participants will be asked to provide a form of identification prior to entry into the study.

See also **5.3 (Screening Procedures and Investigations)**

**Visit 2: Sand fly biting visit**

A further brief assessment of eligibility will be conducted at the beginning of this visit, prior to the sand fly biting which will include a further consent process to check understanding of the study and risks.

The importance of continuing to attend follow up visits to monitor any unforeseen events will again be stressed to the participant

The participants will be asked to avoid wearing deodorants, aftershaves and perfumes on the day. Participants will be asked to wash with a neutral scent-free wash on the day, prior to attending for sand fly biting visit. (This will be provided at the screening visit).

The placement for each biting chamber (containing the sand flies) will be on the volar aspect of the forearm, approximately 2-3 centimetres distal to the antecubital fossa. This site has been chosen for both cosmetic reasons and reduced likelihood of scarring and will therefore be recommended to participants. Either arm can be used, but we will suggest the use of the non-dominant arm. During the FLYBITE study, all participants agreed to the volar aspect of the forearm as an agreed area for sand fly biting, hence it is anticipated that this site will be acceptable to all the participants.

The selected site will be examined to ensure suitable skin quality and photographed prior to placing the biting chambers. Photography will be repeated over the course of the study. Video recording will be employed to record sand fly feeding behaviour but limited to the biting chamber, and not so that the participant could be identified. The area around the biting chamber will be marked with a suitable marker pen. This will ensure that visual and dermatoscopic inspection can occur accurately at the bite site. Participants will be asked to mark this area with a pen that will be provided, if they are happy to do so in order that the biting region can be readily identified at follow-up and in pictures. Data relating to sand fly behaviour will be recorded by study investigators. Participants' sand fly biting experiences will also be recorded. This can include a brief description that may be recorded in the CRF including any features such as pain and itch. A visual analogue score may be used to record these features (see also Diary Card section below). It is feasible that participants may give a video account of their experiences, although this will be optional and the participant will have the choice to for this to remain anonymous (i.e. any recognisable features will be hidden).

All participants will be issued with a study participant identity card and encouraged to contact the research team if there are any problems.

Participants will be required to stay within the TRF for 2 hours after the end of sand fly biting procedure. During this time, observations (including temperature, heart rate, blood pressure, respiratory rate) will be performed at 30 minutes intervals. Further observations will be taken if there are any significant issues either reported by the participants or observed by the clinical team. During this period further information such as number of visible bites and dermoscopy will also be recorded. A further clinical review will take place prior to the participant leaving the unit. An OTC antihistamine will be provided with instruction for use if necessary, in the event of a significant localised reaction at the bite site e.g. itch.

Up to two participants will be studied each day of biting, with one in the morning, and one in the afternoon. The biting days will be dependent on availability of sand flies and other logistical features. The biting days will take place on weekdays only.

A successful sand fly bite is defined as the presence of any of (a), (b), or (c):

a) Suspected sand fly biting activity noted by clinical investigators (including video and photography during biting)
b) Bite compatible lesions by dermoscopy or photography immediately after biting
c) Presence of dilated abdomen in any of the sand flies in the biting chamber following withdrawal of biting chamber from participant.

If none of the above are present, a clinical judgement will be made by study investigators as to presence of successful bite

See also section 6.9 Suspected sand fly biting failure.

**Diary Cards**

Participants will be given a diary to record both local (cutaneous) and systemic symptoms from the biting visit until the final follow-up visit using a Visual Analogue Score (VAS). This will not preclude participants reporting any adverse reactions as detailed in section 7.0. The diary card will be collected at each visit and a new blank one provided. Participants will be also given the option to return this electronically by either taking a picture of a physical diary card and emailing it, or filling in a secure electronic version. There will be space to include extra relevant information including new medications and use of any OTC medications (including those prescribed by study clinicians). If an electronic version is chosen, the participant will receive an alert card as below.

The diary card will also contain information about the study for the attention of health care professionals, in case urgent medical care is needed. It will also contain urgent/emergency contact numbers for both the study clinicians and for the NHS.

The diary card or alert card will also have a ruler printed on it, which can be used to record the size of the lesion when photography is taken by the participant.

Diary card data in of its own may not constitute an adverse event, but will aid investigators to determine presence of AEs which will be recorded in the CRF. Presence of any significant or non-solicited grade 3 or higher AEs or any SAEs recorded on the diary card, as judged by investigators, will be recorded on an AE form and reported to the Sponsor.

**Follow-up Visits**

Further assessments will take place at 4, 14, 28, 42, 56, 70, 98, 126, and 154 days after the biting visit. Visits 3, 4, 5, 7, 9, 11 (numbering as per schedule) will take place in person in the TRF, whereas Visits 6, 8, 10, are scheduled to be 'remote visits' conducted by Video Calling. These can be changed to in person visits if there are any clinical problems. The windows of compliance with the protocol for these visits are indicated in the schedule, Table 1, parts 1 & 2.

Participants will be assessed for local and systemic adverse events using a focused history, physical examination, dermoscopy and photography. Blood will also be taken for exploratory immunology analysis as detailed in Table 1. The presence or absence of lesion will be recorded, and the size of a lesion will be measured. It is anticipated that an early lesion may be observed clinically but may not be measurable accurately in terms of size.

Participants will be asked to take digital pictures of the lesion on a twice weekly basis (or greater if necessary and agreed to by the participant), although this will be optional. There will be an SOP provided to give information about how this should occur. A ruler will be provided on the diary card to facilitate picture taking. Participants will be asked if they are happy to take this with their own mobile phone camera (depending on the quality of baseline imaging as determined by study investigators). Alternatively, a digital camera may be provided to facilitate this. Participants will be encouraged to email these pictures/transfer electronically after they have been taken, or else provide them at the next study visit. Further imaging will be taken by study investigators at study visits.

Participant photography will allow study investigators to monitor the development and/or progression of any lesion. It will aid in the decision by the study investigators to attend for a further study visit if needed. Given that the development of a CL lesion may take some time, this process will negate the need for frequent face-to-face follow-ups whilst also capturing useful data that may not be gleaned without daily follow-up.

Follow-up visits will focus on the reactogenicity experienced by participants and the potential development of a CL lesion. If a lesion develops which is ≥ 3mm in diameter and has clinical characteristics of a CL lesion the subject will exit ‘passive follow up’ and enter the biopsy & treatment phase of the study – see the section ‘Potential development and clinical diagnosis of cutaneous leishmaniasis lesions’ below & Table 2.

**Visit 3**
This follow-up visit will be at day 4 ± 2 days either side of the target date determined by the date of biting visit. The participants will be assessed for local and systemic adverse events using a focused history, physical examination and image documentation.

**Visit 4**
This follow-up visit will be at day 14 ± 4 days either side of the target date determined by the date of biting visit. The participants will be assessed for local and systemic adverse events using a focused history, physical examination and image documentation.

**Visit 5**
This follow-up visit will take place at 28 ± 5 days. The participants will be assessed for local and systemic adverse events using a focused history, physical examination and image documentation. Blood will also be taken for analyses as detailed in Table 1.

**Visit 6**
This follow-up visit will be at day 42 ± 7 days. The participants will be assessed for local and systemic adverse events using a focused history and image documentation.

**Visit 7**
This follow-up visit will take place at 56 ± 7 days. The participants will be assessed for local and systemic adverse events using a focused history, physical examination and image documentation. Blood will also be taken for analyses as detailed in Table 1.

**Visit 8**
This follow-up visit will be at day 70 ± 14 days. The participants will be assessed for local and systemic adverse events using a focused history and image documentation.

**Visit 9**
This follow-up visit will take place at 98 ± 14 days. The participants will be assessed for local and systemic adverse events using a focused history, physical examination and image documentation. Blood will also be taken for analyses as detailed in Table 1.

**Visit 10**
This follow-up visit will be at day 126 ± 4 days. The participants will be assessed for local and systemic adverse events using a focused history and image documentation.

**Visit 11**
This follow-up visit will take place at 154 ± 5 days. The participants will be assessed for local and systemic adverse events using a focused history, physical examination and image documentation. Blood will also be taken for analyses as detailed in Table 1.

##### 6.3 Potential development and clinical diagnosis of a cutaneous leishmaniasis lesion

We anticipate that from Visit 4 onwards a subject may develop an early CL lesion at the bite site. Close liaison with our Dermatology expert (AL) will be utilised at this stage of the study. The following criteria will be used to determine the presence of a clinically compatible cutaneous leishmaniasis lesion, following successful sand fly biting:

*If a lesion is present at 14 days (visit 4) or more post-sand fly biting, which is papular, raised, erythematous and ≥3mm in diameter, and clinically compatible with a CL lesion as judged by study investigators, then the participant will be deemed to have a clinically compatible cutaneous leishmaniasis lesion.*

##### 6.4 Procedures and follow up subsequent to clinical diagnosis of a cutaneous leishmaniasis lesion

These are summarised in Table 2 below.

###### 6.4.1 Biopsy visit

If a lesion fulfils these characteristics it will be excised using a formal excision biopsy, according to our excision biopsy SOP (Appendix 2) and submitted to the PK laboratory for parasitological confirmation, again according to our defined SOP (Appendix 3). The biopsy can take place from visit 4 onwards.

The biopsy procedure will be carried out in the TRF. It can either be conducted at the same time as the routine follow up visit when the diagnosis is made, or at newly arranged visit.
The participant will be asked if they are willing to donate a second biopsy (up to 4 mm punch biopsy) from normal skin at the contralateral site on the other forearm. Donation of this second normal biopsy will be completely voluntary and can take place either at the time of biopsy of the suspected CL lesion, or at a future date. The participant will be asked to sign a further consent form for receiving either one or two biopsies, as well as adhering to the subsequent follow up.
Risks as per all biopsies will include a small risk of infection, bleeding and pain as well as scarring.

###### 6.4.2 Follow up post-biopsy / biopsies

Study subjects will be followed up 10 days after biopsy / biopsies for review and suture removal. There will be further follow ups at days 30, 60, 90, 180, and 360 after biopsy. If any lesions develop during follow up at, or near the original CL biopsy site, that are clinically compatible with recurrent cutaneous leishmaniasis, they will be treated according to relevant guidance and specialist input. The follow ups at days 90, 180, and 360 are to confirm on-going cure, and are of similar duration to the recommended follow ups at 3 months and 1 year post-treatment of clinical CL. [21] Should any unexpected issues in clinical management arise, we will liaise with, and refer the case to the Department of Infectious Diseases, Royal Hallamshire Hospital, Sheffield or (other regional specialist as appropriate). It is feasible that some of these visits can take place virtually, at the discretion of the CI and if there are no significant concerns expressed by the participant.

**Table 2: Schedule of Visits, Treatment Phase, from day 0, the day of biopsy**

| | **Visit 1** Day 0 | **Visit 2** Day 10 | **Visit 3** Day 30 | **Visit 4** Day 60 | **Visit 5** Day 90 | **Visit 6** Day 180 | **Visit 7** Day 360 |
|---|---|---|---|---|---|---|---|
| Window | - | +/-3 days | ± 3 days | ± 7 days | ± 14 days | ± 21 days | ± 21 days |
| History, Examination, Image documentation | X | X | X | X | X | X | X |
| Informed consent to biopsy / biopsies, treatment and follow up | X | | | | | | |
| Excision biopsy / biopsies | X | | | | | | |
| FBC, U+E, LFT, CRP | X* | | X | | X | | |
| *Leishmania* serology | X* | | | | X | | |
| Peripheral blood mononuclear cells & immune monitoring | X* | | X | | X | | |
| Quality of Life questionnaires | X | X | X | X | X | X | X |
| Blood volume | 50ml | 0ml | 45ml | 0ml | 50ml | 0ml | 0ml |

* - Depending on the transition from Table 1, Challenge Phase, these investigations will not be repeated if they have been previously performed in the last 7 days

##### 6.5 Focus Group(s)

We intend to hold a focus group or groups with all the participants from the challenge study after the last subject final follow-up visit. This will be done using our established protocol and methodology which was utilised successfully in the non-infected sand fly bite study. Because of the design of the study with (a) variable numbers and (b) the variable lengths of time taken to complete the study, as well as the length of the study, it is not possible to be precise as to the numbers of subjects that may be available for participation in the focus group. However, if 4 or more participants are available to take part in a focus group then this will take place. It may also be more pragmatic to hold two, or even potentially three focus groups, the initial focus group taking place before all participants have completed the study. Such focus groups will be at times that are mutually agreed with study participants and clinical staff. The decisions on the timing and numbers of subjects in the focus groups will be made by the study team in conjunction with the health psychologist (Prof Jones) and whether 1, 2 or 3 focus groups are to take place. The ideal number of participants in each focus group is 4 – 8 participants. The focus group(s) will take place at the University of York, York, YO10 5DD. It is feasible that the focus group(s) could take place virtually if an in-person meeting is not possible.

The aim of the focus group will be to elicit the participant's experiences of participation in the CHIM in greater depth and gather information to help inform the design any subsequent leishmania CHIM studies. The focus group will be digitally recorded (with consent) and it is anticipated that the focus group will last between 2-3 hours based upon the initial public involvement activity and the FLYBITE focus group. It will be undertaken by GJ who has experience in conducting focus groups and using this type of methodology. The focus group will be fully transcribed verbatim and analysed using NVivo software (QSR International Pty Ltd). An inductive content analysis approach will be employed.[(28)] To establish the trustworthiness of the analysis, one member of the team will independently read the transcripts line by line and identify emergent themes.[(29)] A second member of the team will independently check a proportion of these (50%) to verify the coding. Discussion of, and agreement upon, common patterns and broader themes from the participant's experiences will be reached between these colleagues. Any dissident views and areas of diversity will be considered and discussed with the wider study team.

Participants will be asked about their experiences of taking part in this study, centred on the following points:

(a) The recruitment process (to include screening and pre-screening)
(b) The procedures involved in the *Leishmania* challenge visit itself
(c) The experience of being bitten, and the subsequent evolution of the lesion
(d) The follow-up visits (to include testing procedures/photography)
(e) The biopsy procedures
(f) The topical treatment used
(g) Any safety concerns and if so, how the participants felt about the processes in place to mitigate these
(h) If taking part in this study is what the participants expected and if not, what was different or unanticipated
(i) Potential improvements for subsequent studies
(j) Any other discussion points raised by participants

It is feasible the focus group(s) may take place virtually using video conferencing technology. This will only occur after discussion with the SSG.

#### 6.6 Additional Visits

Additional visits and assessments may be required to evaluate an adverse event and/or to identify a diagnosis. As per normal clinical standards of care, a further biopsy might be necessary to provide a differential diagnosis, for example later during the treatment phase of the study. Such visits and assessments are deemed compatible with the protocol.

#### 6.7 Virtual visits

Participants will be able to conduct select visits via phone or email at the discretion of the CI. This will only be possible if there are no complications noted, and the participant has been compliant with the study schedule up until that point. Evidence of compliance will include attending for blood tests and sharing diary card and participant-recorded photography in a timely manner. This virtual visit will be possible so long as the interval between face-to-face visits does not exceed 3 months (except the final visit which may take place virtually at the discretion of study investigators). It is also feasible that video calling be used to conduct some virtual visits. An NHS accredited app such as Nye (https://www.meet.nye.health) can be used to conduct such a visit or other widely available technology such as Zoom.

#### 6.8 Failure to participate in a visit

If a participant fails to take part in a study visit, in the first instance a repeat visit will be offered within the visit window. If the participant is unable to attend this visit, they will not be excluded, but a further visit will be offered as close as possible to the visit window. If this occurs on more than one occasion for any given participant, it will then be deemed a protocol deviation, and a file note prepared and submitted to the Sponsor Representative. All visits outside of the windows in the schedule will be discussed at the SMG.

#### 6.9 Suspected sand fly biting failure and participant replacement

The evidence for sand fly biting includes -

(a) Participant-reported biting sensation during, and immediately after biting
(b) Suspected sand fly biting activity noted by clinical investigators (including video and photography during biting)
(c) Presence of bite compatible lesions by dermoscopy or photography immediately after biting
(d) Presence of a dilated abdomen in any of the sand flies in the biting chamber following withdrawal of biting chamber from participant.

Our experience with the FLYBITE study leads us to believe that the possibility of sand fly biting failure is low. Nevertheless, it seems wise to consider this possibility and incorporate into the protocol. If none of b, c, or d were observed this would strongly suggest absence of a successful bite, and a negligible likelihood of *Leishmania* transmission to the participant. Visits 3, 4, & 5 out to 28 days would still be completed, to check whether any later changes at the site indicated that successful biting had occurred. If no changes were observed the participant would be deemed to have failed biting, and further clinic visits would cease. We would organise a later Video Calling follow up at 6 months to check on the participant’s welfare.

If sand fly biting failure was determined in any participant at the 28 day follow up (visit 4), an additional volunteer may be recruited, so that the size of the cohort in follow-up need not be limited by biting failure. Such an additional volunteer would be assigned a unique identification number.

##### 6.10 Lack of development of a suspected cutaneous leishmaniasis lesion

If a subject had experienced successful sand fly biting (as defined above) but failed to develop a lesion at the bite site during follow up out to visit 11 (day 154), then that would constitute the last formal study visit. The subject would be provided with a specific information leaflet (Appendix 5) and instructed to contact the study team should any lesion(s) subsequently appear.

##### 6.11 Failure of parasitological confirmation of a suspected cutaneous leishmaniasis lesion

If a subject developed a suspected CL lesion that was biopsied, but then the parasitological investigations were negative, then the investigators CL, AL, PK, and VP would review the clinical and histological data, to decide on a presumptive diagnosis and any necessary management. Formal follow up in terms of the study would cease at that point, but additional visits in terms of alternative diagnoses would be carried out as indicated.

##### 6.12 Safety of staff and participants during visits

We have incorporated several safety features into the conduct of this study in relation to the recent SARS-CoV-2 pandemic. Safety of both participants and study investigators and staff is of the utmost importance whilst conducting this study.

- All participants will be given allocated time slots to attend and overlaps between participants will be avoided.
- Participants will attend via a dedicated entrance to the building which leads directly into the clinical area and is not used by non-study staff during study visits.
- The clinical area will not be accessible by non-study staff during the study visits.
- All surfaces and equipment will be cleaned using an appropriate disinfectant between participant visits, including the sand fly biting chamber (e.g. Tristel Fuse or similar product).
- When examining and during contact with participants, disposable gloves and an apron will be used by study investigators.
- Surgical masks and/or visors will be used in accordance with NHS and research practice depending on the latest government advice at the time of the study.
- Participants will be encouraged to wear face coverings in accordance with any national guidance.
- The minimum number of study investigators will be present at any one time with the participant to ensure adequate distancing (this will be typically be 2 investigators).
- Advice for the conduct of this study has been sought from specialists from The Department of Infection and Tropical Medicine, Sheffield Teaching Hospitals NHS Foundation Trust, Sheffield.

If a participant develops a fever during the conduct of this study, they may have to undergo further testing as dictated by government advice at the time of the study. This may also require a period of isolation for the participant and their household. It was noted in the FLYBITE study that some study participants developed a subjective fever between visits, hence it is feasible in this study that this could develop.

In the unlikely event a fever develops during the first 48 hours after sand fly biting, in accordance with any government advice, self-isolation may necessary. However, if the fever settles within 48 hours, it can be concluded that the fever is most likely due to sand fly biting and the participant can terminate any period of self-isolation. A significant fever in this situation is noted as >37.7C.

Participants will be advised that if they develop a fever before attending for a study visit, they must not attend, including if the fever and any subsequent self-isolation period overlaps with a visit date. Participants will be provided with a thermometer for use at home to substantiate any subjective fever they may develop and at the request of study investigators. In the event that a participant is unable to attend a visit due to any isolation period being necessary, then the visit may be deferred until such a time when it may be feasible. At the discretion of the study investigators, it is possible that these visits may also be ‘virtual visits’, in addition to those already described in this protocol.

Participants may also be required to self-isolate for other reasons or symptoms as per any national guidance and will be supported in doing so. This will include deferring visits and/or virtual visits as described above. A missed visit in the context of symptoms requiring self-isolation as per national guidance will therefore not constitute a protocol deviation.

Study participants will be encouraged to undertake regular asymptomatic testing for SARS-CoV-2, if such facilities exist and are easily accessible (e.g. lateral flow testing), although this will not be mandatory.

#### 7.0 ASSESSMENT OF SAFETY

The Clinical Investigators are responsible for the accurate recording of all AEs & SAEs. The Clinical Investigators are also responsible for reporting of all SAEs to the Sponsor, the University of York within 24 hours of being aware as detailed in 7.4. Additionally, there will be reporting of some adverse events to the Research Ethics Committee (REC) as detailed in 7.4.

For all adverse events, the Clinical Investigators will take appropriate action to ensure the safety of all participants and staff in the study. All adverse event reporting will be carried out in accordance with the Study-Specific Adverse Event Reporting SOP.

##### 7.1 Definitions

Adverse Event (AE)

An AE is any untoward medical occurrence in a participant who has consented and is participating in a clinical study, including occurrences which are not necessarily caused by or related to that investigational item (in this case Sand Flies). An AE can therefore be any unfavourable and unintended sign (including an abnormal laboratory finding), symptom or disease occurring during such a clinical study. Any adverse event noted prior to Day 0 (leishmania challenge) does not need to be recorded.

Information on adverse events will be collected on the day of challenge and at follow up visits as defined in the schedule. AEs will be recorded by study staff through direct questioning and examination. Events will be graded according to the table in Appendix 1. In addition, systemic laboratory adverse events will be collected through routine laboratory testing according to the schedule. These will be recorded on the standard laboratory report. The information obtained will be recorded in the participants file.

Any results that are outside of the normal range for the laboratory at the local hospital but in the opinion of the CI are not clinically significant and do not meet the criteria for being classed as an AE will not be classed as such. For events or results that are not specified in the tables, a decision will be made by the CI or a study physician, as delegated by the CI, as to whether the result is clinically significant. Only those events or results that are deemed clinically significant will be classed as AE's.

Serious Adverse Event (SAE)

An SAE is an AE that results in any of the following outcomes, whether or not considered related to the sand fly bite.

- Death (i.e. results in death from any cause at any time)
- Life-threatening event (i.e. the participant was, in the view of a clinical investigator, at immediate risk of death from the event that occurred). This does not include an AE that, if it occurred in a more serious form, might have caused death.
- Persistent or significant disability or incapacity (i.e. substantial disruption of one's ability to carry out normal life functions).
- Hospitalisation, regardless of length of stay, even if it is a precautionary measure for continued observation. Hospitalisation (including inpatient or outpatient hospitalization for an elective procedure) for a pre-existing condition that has not worsened unexpectedly does not constitute a serious AE.
- An important medical event (that may not cause death, be life threatening, or require hospitalisation) that may, based upon appropriate medical judgment, jeopardise the participant and/or require medical or surgical intervention to prevent one of the outcomes

listed above. Examples of such medical events include allergic reaction requiring intensive treatment in an emergency room or clinic, blood dyscrasias, or convulsions that do not result in inpatient hospitalization.

- Congenital anomaly or birth defect.

**Table 3.0: Solicited Adverse Events**

| Site | Adverse Event |
|---|---|
| Local (at the site of bite) | **Symptoms**<br>Itch<br>Pain / Discomfort<br><br>**Signs**<br>Erythema – grades 1,2,3,4<br>Swelling - grades 1,2,3,4<br>Blister - Vesicle < 5mm, or Bulla ≥5mm |

See Appendix 1 (Solicited study adverse reactions) for further explanations of solicited adverse events.

#### 7.2 Causality Assessment

For every AE, an assessment of the relationship of the event to the sand fly *Leishmania* challenge will be undertaken by a Clinical Investigator.  An interpretation of the causal relationship of the intervention to the AE in question will be made using clinical judgment, based on the type of event; the relationship of the event to the time of challenge and alternative causes such as intercurrent or underlying illness and concomitant therapies.

**Table 4.0: Guidelines for assessing the relationship of an AE to sand fly *Leishmania* challenge**

| | | |
|---|---|---|
| 0 | No Relationship | Adverse events that can be clearly explained by extraneous causes and for which there is no plausible association with study product. Or adverse events for which there is no temporal relationship |
| 1 | Unlikely | Adverse events that may be temporally linked but which are more likely to be due to other causes than this study |
| 2 | Possible | Adverse event that could equally well be explained by the study or other causes, which are usually temporarily linked.<br>Or of a similar pattern of response to that seen with other vector biting studies. |
| 3 | Probable | Adverse events that are temporarily linked and for which the study product is the more likely explanation than other causes.<br>Or known pattern of response seen with other vector biting studies. |
| 4 | Definite | Adverse events that are temporarily linked and for which the study product is the most likely explanation.<br>Or known pattern of response seen with other vector biting studies. |

#### 7.3 Reporting Procedures for All Adverse Events

All adverse events occurring during the study observed by a Clinical Investigator or reported by the participant whether or not attributed to study procedure will be reported in the CRF. The severity of clinical and laboratory adverse events will be assessed according to the AE grading located in the appendix section of the protocol.

We know from the results of the FLYBITE study that successful sand fly biting was universally associated (all cases) with grade 1 & 2 'adverse events' as detailed above (Table 3). As these are in reality 'intended AEs', solicited study events will be recorded in the CRF but not reported to the sponsor as an AE (or recorded on an AE form) unless Grade 3 or higher (see Appendix 1).

##### 7.3.1 Follow up

All adverse events that result in a participant's withdrawal from the study or that are present at the end of the study, will be followed up until a satisfactory resolution occurs, or until a non-study related causality is assigned, or if deemed clinically stable by study clinical investigators.

##### 7.3.2 Ongoing and end of study reporting

At the conclusion of the study all AEs / SAEs will be tabulated. Analysis and subsequent conclusions will be included in the final study report.
The CI will send the final study report to the sponsor and REC within 12 months of the End of Study Declaration.

##### 7.3.3 Emergency Contact process

All study participants will be given emergency contact numbers to report and discuss any issues including AE's. There will be an out-of-hours rota for clinical staff, with a single phone number for study participants to call. In addition to the emergency number, a phone number for the study office (during working hours) and study investigator email address will be provided.

##### 7.3.4 Unexpected screening findings

Participants will also undergo screening including a history, physical examination and blood tests (as detailed above). If there are any abnormal or unexpected findings from these screenings, this will be discussed with the participant by the clinical team. The participant will be referred to an appropriate medical speciality, or their GP. This may not necessarily exclude the participant from the study if this does not otherwise impact on the inclusion and exclusion criteria. The decision for ongoing participation will be made in conjunction with the SMG.

#### 7.4 Reporting Procedures for Serious Adverse Events

The Investigator will complete an SAE report form and notify the Sponsor within 24 hours of becoming aware of the SAE. All SAEs that are causally linked to the administration of the protocol and unexpected will be reported to the REC within 15 days of being made aware of the event. In addition, all non-solicited grade 3 & 4 AEs regardless of relationship will be referred to the REC for consideration of whether that they constitute the need for a halt to the study and/or protocol amendment.

#### 7.5 Withdrawal of Participants

A participant has the right to withdraw from the study at any time and for any reason and is not obliged to give his or her reasons for doing so (including during the sand fly biting itself). A Clinical Investigator may withdraw the participant at any time in the interests of the participant's health and well-being. If withdrawal is due to an adverse event, appropriate follow-up visits or medical care will be arranged until the adverse event has resolved or stabilised. If a participant is considered to have failed the screening assessment or withdraws from the study at any time, either by choice or on the recommendation of clinical personnel, data and samples collected up to that point will remain available for analysis as part of the study, unless a participant withdraws consent for this.

Given the nature of the CHIM, the study intervention cannot be withdrawn after exposure to an infected sand fly bite. As such study participants will be strongly encouraged to continue to attend follow-up visits. If a study participant decides to withdraw from any further follow-up, the sponsor will be informed, and the participant will be given contact details of the study investigators and where to seek medical attention if necessary. The participant's GP will also be contacted to inform them of events and the referral pathway if the participant later attends their GP with a suspected CL lesion. If a participant withdraws from the study, then later decides to return they will be allowed to do so but only after agreement from the SMG. The decision may be made by SMG to excise an early CL lesion in such participants to avoid an untreated CL lesion with loss to follow-up. It is noted that *Leishmania major* is not associated with dissemination and in most healthy immunocompetent subjects an untreated lesion will self-heal.

If the participant is not enrolled into the biting portion of the study after being deemed ineligible, all screening data and samples collected up to that point will remain available for analysis as part of the study unless the participant withdraws consent for this. If there is deemed to be any information relevant for the participant's GP to be made aware at this stage, consent will be taken to write to the GP. Consent for GP communication will be taken at the initial screening visit.

If a participant wishes to withdraw from the study, and then requests for their existing, un-analysed samples to be destroyed, the CI will initially discuss with the Sponsor what appropriate action should be taken. The final decision will then be made in conjunction with the participant as to which samples may be used in analyses if at all.

If they withdraw, then participants may be replaced in the study. A standby list of potential further participants will be maintained in accordance with the study documentation.

##### 7.6 Pregnancy Reporting

If a female participant becomes pregnant during the 90 days following infected sand fly biting, she will be followed up as other participants. The pregnancy will be reported to the Sponsor in accordance with SOP for Research Related Adverse Event Reporting Procedure. The treatment decision in this case will be based on a discussion with the study clinical team and the participant.

Study investigators may offer additional pregnancy testing during the study if deemed necessary.

##### 7.7 Temporarily Interruption or Discontinuation of the Study

**Definitions:**

"Discontinuation" is the permanent withholding of further sand fly exposure visits from all or some study groups in the study.

"Interruption of sand fly biting" is the temporary withholding of sand fly exposure if a serious adverse reaction is experienced during the study.

**Clinical Criteria for Interruption or Discontinuation of *Leishmania* challenge**

If any of the following occur, interruption or discontinuation of all further *Leishmania* challenge will take place and the Sponsor will be notified within 24 hours.

(1) Death in any subject in which the cause of death is judged to be possibly, probably or definitely related to sand fly bite exposure
(2) The occurrence in any subject of an anaphylactic reaction to sand fly bite
(3) If two or more participants experience an unexplained, unexpected grade 3 or 4 clinical or laboratory event (confirmed on attendance or repeat testing) that has not resolved within 72 hours and considered possibly, probably or definitely related to *Leishmania* challenge.

The Sponsor will notify the Study Steering Group (SSG) and the SSG and / or the Sponsor will determine whether or not to call an unscheduled meeting to review the safety data, and whether or not to hold further sand fly biting visits until this has taken place.

If the study is halted or stopped for a reason involving risk to a participant's health or safety then an Urgent Safety Measure will be implemented in accordance with the Safety Reporting SOP. If the study is halted or stopped for any other reason the CI or Sponsor representative will notify the HRA not later than 15 days from the date of the halting of the study in accordance with Safety Reporting SOP.

Restarting the study will be a substantial amendment.

As *Leishmania* challenge is a new clinical study area we have also included on-going real time safety reviews within the protocol (see 4.5). No more than 2 participants will undergo sand fly biting on any given day, and no participants will undergo sand fly biting simultaneously. Participants will be scheduled to attend shortly after the biting visit (see schedule). The development of any SAE or non-solicited grade 3 AE at the first post-biting review considered possibly, probably or definitely related to the biting procedure will result in a temporary halt and review of the sand fly biting parameters. Therefore, we will review the safety outcomes 4 days after all biting procedures in real time for all subjects.

If an SAE or non-solicited grade 3 AE had been recorded the CI (CL) and PIs (VP, AL & PK) would review both the clinical event, and the biting parameters in terms of the number of sand flies, the species and length of exposure, with regard to progression of the study. If non-solicited Grade 3 / exaggerated reactions had been observed it would be likely that the Investigators (as above) would seek a protocol amendment to decrease the number of sand flies present within the biting chamber, or length of exposure to mitigate any exaggerated response. At the discretion of the clinical investigators (CL, VP, AL) it would be possible that a sufficient number of grade 2 AEs, would also result in such a halt to the study and consideration of a protocol amendment.

##### 7.8 Sponsor

The Sponsor(s) reserves the right to terminate the study at any time.

##### 7.9 Blood Tests

If any blood tests are out of the laboratory range, they will be recorded in the CRF and reviewed by a clinician. For significantly out-of-range blood tests, an out-of-hours emergency clinician contact number has been provided to the testing laboratory (a separate SOP is available which details the criteria to inform a clinician urgently).

A decision will be made by clinical investigators to record blood tests which are out of the laboratory range as clinically significant on the CRF, if applicable. Only clinically significant abnormal blood tests will be reported as AEs, as judged by study investigators. Reference ranges are provided in Appendix 1 to serve as a guide, although the decision regarding clinical significance will be made by clinical study investigators.

##### 7.10 Close of Study

The close of study is defined as 3 months after the last focus group.

#### 8.0 MANAGEMENT OF DATA, SAMPLES AND STUDY PROCEDURES

##### 8.1 Source Data and Case Report Forms (CRFs)

Consenting participants will be allocated a CRF which will act as the source data. The CRF will hold personal identifiable information on the participant, including name, address, and date of birth and participant study number. The participant's file will be held at the TRF in a secure location. Permission will be obtained as part of the informed consent process to allow the research team and other responsible individuals access to the participants' study records.

The University will put in place appropriate technical and organisational measures to protect personal data and/or special category data. For the purposes of this project we will store data on dedicated servers and / or on the University of York central data store, which provides secure long term storage for data, including daily backups, according to the University of York Research Data Management Policy.

A study master file (SMF) will be created to include the study protocol, original signed consent forms and ethics/governance documentation. It will also include a list of study group participants, their email address and their telephone number. The study master file will be stored at Department of Biology in a secure, fire and rodent-proof cabinet that is only accessible to authorised members of staff.

Information will be treated confidentiality and shared on a need-to-know basis only. Only the minimum amount of data necessary for the project will be collected. In addition, data will be anonymised or pseudonymised wherever possible. As there is only one study site, the terms 'study master file' and 'study site file' (SSF) are used interchangeably within this protocol.

Data collected directly from the participant or from medical examinations will be entered directly into the CRF. All laboratory reports will be filed in the CRF after review and signed off by a medically qualified Clinical Investigator. Data collected at the clinical site will be transcribed directly onto case report forms. The type of data to be recorded in the CRF will be in line with the details provided in the study schedule section. Appendix 2 provides details of what constitutes source data in this study. CRF's will be identified with participant study number and initials. No personally identifiable information will be sent outside of the University except for communication with the GP.

The statistical investigators will undertake data management responsibilities, which include the provision of study database, data entry and validation procedures. The statistician will work with the study team to draft the CRFs. A separate statistical analysis plan document will be available in conjunction with the study statistician.

##### 8.2 Screening and Study Entry Logs

Screening and study logs containing study numbers, name, date of birth will be kept in the study site file, which will be kept in a secure location at the study site, with access restricted to study staff only.

##### 8.3 Access to Data

The investigators will maintain appropriate medical and research records for this study. The Chief Investigator, co-investigators, clinical research nurses and clinical trials assistant will have access to records. The investigators will permit authorised representatives of the sponsor(s), and regulatory agencies to examine (and when required by applicable law, to copy) clinical records for the purposes of quality assurance reviews, audits and evaluation of the study safety and progress.

##### 8.4 Data Protection

The study protocol, documentation, data and all other information generated will be held in strict confidence. No information concerning the study or the data will be released to any unauthorised third party, without prior written approval of the sponsor.

Data will be accessible to the study team at the University of York only. Anonymised data may be reused by the research team or other third parties for secondary research purposes following express approval of the sponsor.

##### 8.5 Archiving of Data

All relevant study documents and data will be securely stored for a minimum of 15 years after the close of the study in accordance with SOP on Archiving of Research Study Documents. All data recorded at the York Teaching Hospitals NHS Foundation Trust (including results from blood tests) will be kept according to local trust protocols but in an anonymised fashion and participants will be unable to withdraw consent for this blood test data to be destroyed. Participant data from all individuals entered in to the sand fly biting portion of the study will be entered into an electronic database. There is no requirement for failed screening participant data to be entered in to the database, however data may be entered at the discretion of the CI. If a participant withdraws consent for data collection and is not entered into the biting portion of the study, their data will not be entered in to the electronic database at the discretion of the CI.

##### 8.6 Confidentiality

Participants will be identified only by their study participant number, initials and date of birth on any documentation or samples that leave the study site. No personally identifiable data will be stored with external organisations. Participants will not be identifiable in any study report or publication and any data (including pictures) will be pseudonymised, unless they give consent for photography or videos to be shared.

##### 8.7 Management of Biological Samples

Blood samples taken at the screening visit for routine laboratory parameters and blood borne viruses during the study will be tested at the York Teaching Hospital Foundation Trust Pathology laboratories. These samples will be identified by the participant's study number, date of birth and initials as required by the laboratory and will only be used for immediate testing and short-term storage as per trust protocol.

All other samples will be tested at the Translational Research Facility, HYMS / Department of Biology, University of York and York Hospitals NHS Foundation Trust. This is where they will be studied by the research team. Samples will be tested in secure laboratory facilities accessible only by authorised research staff. Samples will be labelled with a unique ID number and the date but no personal details, so they cannot be identifiable. The samples will only be used in research projects that have been independently reviewed and approved by the Department of Biology

Ethics Committee. The samples will be stored for 5 years and may be used for additional future research with permission of the study participants. After this period, the samples will be transferred to the research tissue bank. The University of York holds an HTA licence (No. 12604) and samples will be stored according to local HTA standard operating procedures.

Samples tested at the York Hospital will be subject to NHS guidelines on storage, handling and disposal. These samples will not be stored for further research purposes, although testing results will be stored anonymously according to NHS local trust protocols and participants will be unable to withdraw consent for this blood test data in order for them to be destroyed. These results will not be linked to participant's existing NHS records. In the event of abnormal results, these will be shared with the participant's GP with their consent.

Samples tested at the research laboratories in the Translational Research Facility, Department of Biology, University of York, will be stored in locked, alarmed -20ºC and -80ºC freezers in the same facility. Access to the samples will be restricted to members of the research team. Samples will be assigned a participant number, date and other study identifiers; however no other patient identifiable data (including name and date of birth) will be recorded.

PBMC (peripheral blood mononuclear cell) samples will be stored in a limited access, alarmed and monitored liquid nitrogen dewar at the Translational Research Facility, University of York. Only members of the research team will have access to the samples.

Sand flies collected following biting will be culled and then stored in locked, alarmed -20ºC and -80ºC freezers, as above. They will be subject to *Leishmania* and other testing compatible with the tests described in study procedures (section 6.0).

Biopsy tissue will be stored in the TRF.

**8.8 Risk Assessment**

A full study risk assessment will be carried out in accordance with the Sponsor's SOP.

**8.9 Study Management**

The Sponsor delegates certain roles and responsibilities to the CI, as detailed in the SOP entitled "Delegation of Tasks for LEISH_Challenge".

LEISH_Challenge will be managed by a Study Steering Group (SSG), comprising the project scientific lead (Kaye), the Chief Investigator (Lacey), the study statistician (Allgar), Principal Investigators (Volf, Layton, Schwartz, Parkash, Jones), entomologist (Sadlova),) and the Project Manager (Greensted). The SSG will report to the Sponsor when necessary. The SSG will also review and comment on development of SOPs for experimental and clinical work and the development of Case Record Files (CRFs) and Study Master File (SMF).

A Study Management Group (SMG) comprising study clinicians (Lacey, Layton, Parkash), entomologist (Ashwin), study nurses and CTA, and the Project Manager (Greensted) will review ongoing clinical and safety data and decide on continued recruitment. The decisions of the SMG will be communicated to the SSG. AEs will be reported to the Sponsor according to the Protocol.

The Clinical Management Group (CMG), comprising study clinicians (Lacey, Layton, Parkash), will review any adverse event data in real time in order to make rapid decisions about safety. This is with particular relevance to safety data at the day 4 post-biting visit.

##### 8.10 Study Monitoring

The study will be monitored and audited in line with GCP requirements and all adverse events will be recorded and reported. A monitoring plan will be established by the Sponsor prior to the start of the study.

#### 9.0 ADAPTIVE DESIGN AND SAMPLE SIZE CONSIDERATIONS

Adaptive clinical trial designs have become more widely discussed in the last decade or so,[30] and are now increasingly being regarded as useful and beneficial in specific circumstances within clinical research.[31] Adaptive clinical trials are an innovative trial design aimed at reducing resources, decreasing time to completion and number of patients exposed to inferior or harmful interventions, and improving the likelihood of detecting treatment effects or demonstrating new paradigms. The last decade has seen an increasing use of adaptive designs, particularly in drug development. They frequently differ importantly from conventional clinical trials as they allow modifications to key trial design components during the trial, as data is being collected, using pre-planned decision rules.

The development of controlled human infection models has parallels with various historical examples of medical experimentation as referred to in 2.4. There are also similarities in terms of scale in that the initial description of various CHIMs has been in small numbers of subjects. For example, the first description of a malaria CHIM in the modern era using mosquito transmission was in 4 subjects,[32] and the first description, again in the modern era of infusion of blood stage *Plasmodium falciparum* inocula as a CHIM was in five subjects.[33] The malaria blood stage CHIM is an efficient model for malaria vaccine development for two principal reasons (i) the efficiency of transmission in the model is 100%, and (ii) the readout for vaccine efficacy is measured by qPCR through the time to detectability, and then the parasite multiplication rate, and this is of course a continuous variable. This gives ready statistical power to the evaluation of candidate vaccines in the CHIM.[34]

We therefore face two inherent issues in the development of our *Leishmania* CHIM and its subsequent clinical applications in that (i) we need a model that is highly efficient, i.e. the take rate should be 100% if possible, because (ii) the outcome of the challenge is a Yes/No readout, i.e. a categorical variable, and this is a less powerful readout, statistically, than a continuous variable.

We have therefore performed a series of sample size calculations to explore how our proposed *Leishmania* CHIM might operate in practice in the evaluation of candidate *Leishmania* vaccines. Assuming we wanted to do a simple two arm trial of a candidate *Leishmania* vaccine vs placebo using the CHIM that we develop, and we wish to have 90% power to detect a reduction in the number of participants with lesions (Vaccine Efficacy) of 60% with $p<0.05$, then we would need a total sample size of:

- 22, if the take rate in the placebo arm is 100% (6/6)
- 28, if the take rate in the placebo arm is 92% (11/12)
- 36, if the take rate in the placebo arm is 83% (5/6)
- 62, if the take rate in the placebo arm is 67% (4/6)

For vaccine efficacy of 50%, the numbers would be 28, 38, 54 and 90.

We performed further statistical calculations to determine the confidence intervals associated with the higher take rates referred to above. These showed –

| Lesion development / Sample size | Take rate with CI |
|---|---|
| 6/6 | 100% (35 – 100%) |
| 12/12 | 100% (74 – 100%) |
| 11/12 | 92% (62-100%) |
| 10/12 | 83% (52-98%) |

After discussions with statistical experts we felt that take rates greater than 90% that had CIs with a lower bound greater than 60% would provide significant assurance in terms of subsequent development of the model.

We therefore conclude, with regard to all of the above data that -

A. A sample size of 12 is greater than, but similar to other studies developing new CHIM methodologies
B. Only with take rates of >90% do we create a *Leishmania* CHIM that leads to practically achievable numbers in subsequent vaccine studies using the CHIM
C. This justifies our adaptive design where we are trying to achieve take rates of 12/12, or 11/12

#### 10.0 ETHICS

##### 10.1 Declaration of Helsinki

The Investigators will ensure that this study is conducted according to the principles of the current revision of the Declaration of Helsinki, latest amendment 2013.

##### 10.2 ICH Guidelines for Good Clinical Practice

The Investigators will ensure that this study is conducted in full conformity with relevant regulations and with the ICH guidelines for GCP (CPMP/ICH/135/95) December 2016.

##### 10.3 Research Ethics Committee

A copy of the protocol, informed consent forms, any other written participant information and the main advertising material will be submitted to the University of York Ethics Committees and to the Health Research Authority, Research Ethics Committee for approval.

##### 10.4 Participant Confidentiality

No material will be kept on file that refers to the study participant by their full name other than in source documentation kept at the study site. The confidentiality of participants will be respected and maintained at all times.

The participant’s study number only will identify study reports. CRFs, associated database records and blood samples will only be identified by the participant’s study number, initials and date of birth.

##### 10.5 Risks

The Investigators will ensure that the dignity, rights, safety and well-being of participants are given priority at all times.

A risk assessment will be performed to assess the risks and benefits of study participation to the individual participant safety, as well as the risks that underlie the validity of the study results with respect to safety and immunogenicity outcome measurements. The outcome of this assessment will be used to guide the development of procedures with respect to informed consent, confidentiality, study monitoring and audit.

Given the nature of the study, there are some anticipated events. See Table 3 and Appendix 1 for solicited study events.
To date, there have been no reported cases of anaphylactic (life-threatening) reactions to sand fly bites in the medical literature. However, there might still be a small theoretical risk of serious reaction to sand fly bite, just as with any insect bite. Within the very small number of people who develop a serious reaction, a small number of these may be at risk of death. We have appropriate treatment and a medical team on-site at all times in the very unlikely event that a serious reaction occurs.

It is possible that after the biopsy there may still be some parasites left in the body near the site of infection. Normally these help to keep the immune system stimulated and provide protection against reinfection. This might be beneficial if the participant travels to a leishmaniasis endemic region and is exposed naturally. However, the presence of this small number of parasites makes it possible that immunosuppression (e.g. through an organ transplant, HIV or drugs that affect the immune system) could theoretically cause reactivation of the lesion. Although this can happen in other forms of leishmaniasis, we are unable to find any descriptions of this occurring with *Leishmania major* after successful treatment.

Just like with any medical procedure there are some risks involved with the excision biopsy. These are rare and include:
Pain
Infection
Bleeding
Incomplete excision of the lesion. This might mean a repeat biopsy is needed.
Bruising
Numbness at the scar site
Small scar (similar to a chicken pox scar)
Very rarely:-
A small bump might form at the scar site but if this occurs, it is likely to settle over time.
Wound breakdown and ulceration

##### 10.6 Benefits

The results of the study will mainly provide safety information which will help in the development of an effective human challenge model of cutaneous leishmaniasis.

Participants will receive a thorough medical examination. In the event of any abnormal findings, participants will be advised of the best course of action and referred on to the appropriate clinician where appropriate.

All participants will have an assessment of baseline psychological wellbeing. Any significant findings that are highlighted by this assessment will be disclosed to the participants GP if consent for this has been given.

Exposure to sand fly resulting in a cutaneous leishmaniasis lesion, may for some individuals protect against future leishmaniasis infection after further exposure e.g. if participant travels to an endemic setting. This is a potential benefit but study investigators will be unable to quantify this.

##### 10.7 HIV, Hepatitis B and Hepatitis C Testing

Participants will be required to undergo HIV and hepatitis B & C testing as part of the eligibility screening assessment. Participants will receive pre-test counselling in line with current NHS guidance. Any positive tests may be repeated if necessary as per blood testing SOP to determine accuracy of results. If a participant is shown to have confirmed blood-borne virus infection then they will be withdrawn from the study prior to sand fly biting.

##### 10.8 Reimbursement

Participants will be compensated for their time. Details of compensation are provided in section 13.2.

#### 11.0 REGULATORY AND GOVERNANCE ISSUES

##### 11.1 Required approvals

Current regulatory frameworks for this clinical study require that the following approvals and registration must be obtained before commencement of the study:

Clinicaltrials.gov Registration
Ethical approval from HRA, NHS
Ethical Approval from University of York, UK

##### 11.2 Amendments

Both substantial and non-substantial amendments will be submitted according to the Sponsor's SOP: Review and Approval of Amendments.

##### 11.3 GCP, and GLP Compliance

All clinical staff will provide evidence of GCP training as a requirement of their delegation. The study will be monitored and audited in line with GCP requirements and all adverse events will be recorded and reported.

All laboratory procedures will be undertaken in line with standards ordinarily applied at the laboratory.

#### 12.0 INDEMNITY

University of York will act as Sponsor for the project.

##### 12.1 Negligent Harm

Legal liability insurance will be provided for study participants by the sponsor.

##### 12.2 Non-Negligent Harm

**Cover for non-negligent harm (no fault compensation) will be provided for study participants by the sponsor.**

#### 13.0 FINANCE

##### 13.1 Financing

The study will be funded by the Medical Research Council and the Department for International Development. (reference: MR/R014973/1)

##### 13.2 Reimbursement for Participants

Participants will be compensated for their time and for the inconvenience at approximately these rates in relation to the following visits:

Screening visit – £20

*Leishmania* challenge visit £500

Follow-up visits, £40 per visit

Per completed diary card event - £5 (up to once weekly)

Per completed and participant-submitted photograph event - £5 (up to twice weekly)

Biopsy visit £500

Final visit £100

Focus Group visit £100

Total up to approximately £2500 depending on involvement.

Travel costs will be reimbursed in addition in relation to expenses submitted.

Re-imbursement will take place in instalments (but it is feasible due to administrative reasons that the total payment will be given at the end of the study). In certain circumstances, participants may be reimbursed prior to the agreed time points depending on administrative factors and at the discretion of the CI, but only in relation to involvement up until that point. Participants will be reimbursed by an agreed method. If a participant withdraws or is excluded, they will be reimbursed for involvement and visits attended up until the point of withdrawal or exclusion. Participants may decline reimbursement if they wish.

Additional treatments and investigations will be provided free of charge for participants who require it.

#### 14.0 PUBLICATION

The results of the study will be analysed and prepared in a study report for publication in a peer-reviewed professional journal. The Chief Investigator; Professor Charles Lacey, the co-Investigator and Scientific lead; Professor Paul Kaye, Principal Investigators; Professor Petr Volf, Dr Vivak Parkash, Professor Alison Layton, Professor Charle Jaffe, Professor Eli Schwartz and Professor Georgina Jones and study statistician will form the basis of the writing group. Authorship will reflect the work carried out by the Investigators.

**Appendix 1**: **Grading of Clinical and Laboratory Adverse Events**

Based on systems in use at the MRC CTU, DAIDS, and YTHFT.

The tables below define abnormal pathology results. Any results that are outside of the normal range for the laboratory at the York Teaching Hospital NHS Foundation Trust but in the opinion of the CI are not clinically significant and do not meet the criteria for being classed as an AE will not be classed as such. For events or results that are not specified in the tables, a decision will be made by the CI or a study physician, as delegated by the CI, as to whether the result is clinically significant. Only those events or results that are deemed clinically significant will be classed as AE's.

For clinical events in general the following grading will be applied:

- **Grade 1** (mild): Awareness of sign or symptom which may be tolerated with minimal or no interference with activities of daily living. Transient (< 48 hours). May be relieved by over-the-counter medication and no or minimal medical intervention required.

- **Grade 2** (moderate): Notable symptoms and/or greater than minimal interference with ADL's. No or minimal medical intervention required.

- **Grade 3** (severe): Symptoms causing significant incapacity resulting in bed rest or activity reduced by >50% of usual level and/or unable to work. Medical intervention may be required.

- **Grade 4** (extreme / life-threatening): Significant medical intervention / therapy required. Hospitalisation required to prevent permanent impairment or death.

#### Laboratory reference ranges

Any values outside these ranges will not be deemed an AE unless clinically significant (as judged by study investigators).

| Parameter | Male | Female |
|---|---|---|
| **Haematology:** | | |
| Haemoglobin (Hb) | 130 - 180 g/L | 115 - 165 g/L |
| White Blood Cell Count (WCC) | 4 - 11 x$10^9$/L | |
| Platelets (Plts) | 150 - 450 x$10^9$/L | |
| Red Blood Cell (RBC) | 4.3 - 5.8 x$10^{12}$/L | 3.9 - 5.4 x$10^{12}$/L |
| Mean Cell Volume (MCV) | 79 - 101 fL | |
| Haematocrit (Hct or PCV) | 0.39 - 0.5 | 0.36 - 0.47 L/L |
| Mean Cell Haemoglobin (MCH) | 27 - 32 pg | |
| Mean Cell Haemoglobin Concentration (MCHC) | 300 - 370 g/L | |
| Neutrophils | 2 - 8 x$10^9$/L | |
| Lymphocytes | 0.5 - 4.5 x$10^9$ /L | |
| Monocytes | 0.2 - 1.2 x$10^9$ /L | |
| Eosinophils | 0.1 - 0.7 x$10^9$ /L | |
| Basophils | 0.0 - 0.2 x$10^9$ /L | |
| **Biochemistry:** | | |
| Sodium | 133-146 mmol/L | |
| Potassium | 3.5-5.3 mmol/L | |
| Creatinine | 59 -104 µmol/L | 45 - 84 µmol/L |
| Urea | 2.5-7.8 mmol/L | |
| Bilirubin | <21 umol/L | |
| Alanine transaminase/ aminotransferase: | 0 - 45 IU/L | 0 - 34 IU/L |
| Alkaline Phosphatase | 30 - 130 IU/L | |
| Albumin | 35 - 50 g/L | |
| Total Protein | 60 - 80 g/L | |
| Aspartate transaminase/ aminotransferase | 0 – 35 U/L | 0 - 31 IU/L |
| C-reactive protein (CRP) | <5 mg/L | |

#### SOLICITED STUDY ADVERSE REACTIONS

| | Grade 1 (Mild) | Grade 2 (Moderate) | Grade 3 (Severe) | Grade 4 (Extreme) |
|---|---|---|---|---|
| **GENERAL** | | | | |
| Fever<br>l>12 hours | 37.7 - 38.9°C<br>(100.0 – 101.5°F) | 39.0 – 39.7°C<br>(101.6 – 102.9°F) | 39.8 – 40.5°C<br>(103 - 105°F) | >40.5°C (105°F)<br>OR max temp of >105°F |
| Chills/rigors | Mild hot/cold flush requires blanket or occasional aspirin/paracetamol | Limiting daily activity >6 hours, or need regular aspirin/paracetamol | Uncontrollable shaking, treatment from doctor needed | Significant medical intervention (up to and including Hospitalisation) |
| Malaise/abnormal tiredness | Normal activity reduced – not bad enough to go to bed | Fatigue such that ½ day in bed for 1 or 2 days | Fatigue such that in bed all day or ½ day for more than 2 days | Significant medical intervention (up to and including Hospitalisation) |
| General (all over) muscle aches and pains (myalgia) | No limitation of activity | Muscle tenderness, aches/pains limiting activity e.g. difficulty climbing stairs | Severe limitation e.g. can't climb stairs | Significant medical intervention (up to and including Hospitalisation) |
| Headache | No treatment or responds to paracetamol like treatment | Regular paracetamol like treatment needed | Regular strong painkillers needed or other medical intervention required | Significant medical intervention (up to and including Hospitalisation) |
| Nausea | Intake maintained | Intake reduced less than 3 days | Minimal intake 3 days or more | Significant medical intervention (up to and including Hospitalisation) |
| Immediate general reactions (within 6 hours of sand fly bite) | | | Laryngeal oedema insufficient to require intubation; diarrhoea insufficient to require IV fluids, or asthma insufficient to require hospitalisation<br>OR<br>Urticaria, angiooedema | Anaphylactic shock |

| CUTANEOUS | | | | |
|---|---|---|---|---|
| Itch / pruritus | Mild itching, not requiring specific therapy | Moderate itching, requiring intermittent antihistamines | Severe itching, requiring regular antihistamines | Generalised itching, requiring on going medical management |
| Discomfort/pain | Mild pain that responds to paracetamol-like treatment, if needed | Pain requiring regular paracetamol-like treatment | Pain requiring regular strong painkillers | Hospitalisation |
| Erythema<br>(Grades 2,3,4 can be sub-classified as macular or diffuse) | Mild erythema localised to the bite site | Moderate erythema within the aperture of the biting chamber | Diffuse erythema outside the aperture of the biting chamber but < 30% of the arm circumference | Diffuse erythema outside the aperture of the biting chamber and > 30% of the arm circumference |
| Blistering | Fluid filled vesicle(s) at the bite site | Fluid filled vesicle(s) within the aperture of the biting chamber | Bulla / bullae outside the aperture of the biting chamber, with no ulceration | Bulla / bullae outside the aperture of the biting chamber, with ulceration |
| Swelling | Palpable swelling at the bite site | Palpable swelling, within the aperture of the biting chamber | Palpable swelling outside the aperture of the biting chamber | Palpable swelling outside the aperture of the biting chamber, with subcutaneous oedema |

#### Appendix 2. Skin excision biopsy SOP

An excision biopsy of the skin is a very simple procedure performed in the outpatient setting which takes 10 to 15 minutes.

**Step 1- Prepare the Area to be Biopsied**:

Initially perform the study visit-specific history, examination, and image documentation. Then explain the biopsy procedures and obtain informed consent to the biopsy (suspected CL lesion), or biopsies (suspected CL lesion and matched normal skin on the contralateral arm). The informed consent also includes the subsequent study follow up. Next, the areas of skin where the biopsies are to be performed are cleaned with an alcohol swab to ensure sterile conditions.

**Step 2 - Anesthetise the Skin:**

The skin that is going to be removed is anesthetised by injecting a solution of 1% lignocaine and adrenaline just under the epidermis (sub-epidermally) using a 2ml syringe and orange needle. The injection continues until a “bleb” or bubble has formed under the skin greater than 4mm in diameter. The injection will burn slightly (much like a bee sting) due to a pH difference between the skin and the solution. The slight burning will quickly subside and the site will become numb.

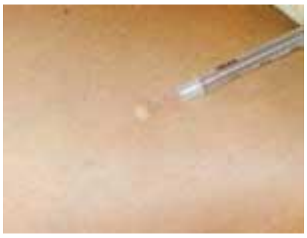

**Step 3 - Check for Numbness:**

After the local anaesthetic has been administered the areas to be biopsied should be checked to ensure that the skin is properly anesthetised. A needle is used to establish whether the skin has gone numb. Great care should be taken not to force the needle into the skin. The test site should be somewhere around the periphery of the area injected. Both of these precautions ensure a viable biopsy for diagnosis later. If the patient confirms no sensation, the biopsy can proceed. A pressure sensation is normal and expected but there should be no pain. If the area requires more anaesthetic a further injection (with a new syringe) is made until the skin is completely anaesthetised.

**Step 4 - Biopsy the Skin:**

Once the skin is anaesthetised, an ellipse is drawn around the lesion providing a margin of normal looking skin using a marker pen. The area to be excised will be approximately 16-20 mm long, and 7-8 mm wide. The use of a length of x 2-3 of the width, with the length in the direction of Lange’s lines, provides tension free closure and optimal healing and cosmesis. A size 15 blade is used, starting at one apex and following the arc of the ellipse, whilst holding the skin in traction. The process is repeated on the other side of the ellipse.

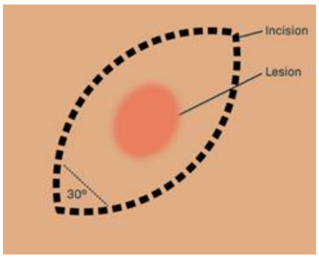


**Step 5 - Remove the Skin Biopsy:** Once the elliptical incision has been made it is normal for the area to bleed. Haemostasis is achieved by direct pressure. Using toothed forceps, the tissue is carefully elevated and dissected at the level of the subcutis with the scalpel. Care is taken to dissect the tissue along even planes. Haemostasis is again achieved with pressure, or light cautery if needed.  The biopsy is immediately placed in the vial containing Schneider's insect media, as below in Appendix 3.

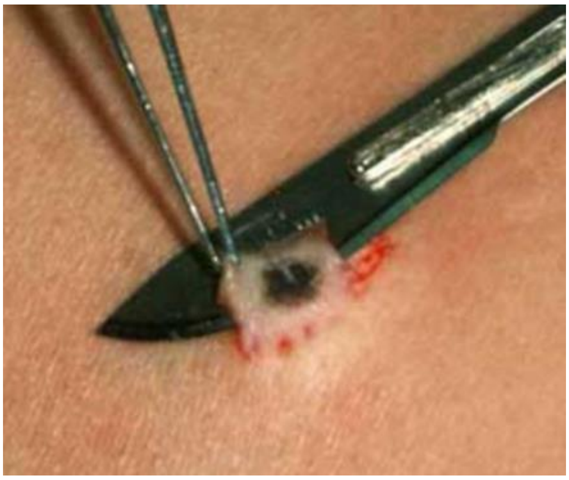

**Step 6 – Suture and Bandage the Biopsy Site:** Wound closure will be achieved with 3 or 4 simple interrupted sutures using 5-0 Ethilon. The biopsy site is then covered with a small adhesive dressing.

**Step 7 – Biopsy Site Care**: The biopsy site should be kept clean. The site should not be submerged in water (i.e. no swimming, hot tubs, baths etc) for 48 hours. In the long term, minimal scarring may occur. In a few cases the biopsy site may form a protrusion or bump but will heal normally

**Step 8 – Suture Removal**

If the sutures are still *in situ* at the 10 day follow up they are removed using gentle traction on one the 'ears' and carefully cutting the 'intra-dermal loop' on the skin side of the knot, and then gently pulling the whole suture clear.

**Appendix 3. Parasitological Confirmation SOP**

Parasitological confirmation will be determined using 3 different approaches, histology, culture and qPCR. All 3 approaches are performed on the skin biopsy.

**Step 1 – Skin Biopsy Collection:** Biopsy is collected as appendix 2, at step 6 the biopsy will be transferred to a vial containing Schneider's insect media and transferred to a biosafety cabinet.

**Step 2 – Skin Biopsy Preparation:** Remove the biopsy from vial using tweezers, place onto a glass slide and cut laterally in half using scalpel. Place one piece in a vial containing 4% paraformaldehyde, for histology*. Gently scrape across the surface of the remaining half collecting scrapings on glass slide, for culture. Place remaining biopsy piece in vial and immediately transfer to dry ice then -80°C, for qPCR.

**Histology**

**Step 1 – Tissue Preparation:** Tissue stored overnight at 4°C in 4% PFA. Transfer tissue to Leica tissue processor and embedding station for preparing FFPE tissue block.

**Step 2 – Slide Preparation:** 5µm sections will be cut from the FFPE tissue block.

**Step 3 – Haematoxylin and Eosin:** A slide will be deparaffinised and brought to water stained with Harris Haematoxylin followed by Eosin then dehydrated and mounted. Slides will be examined under a light microscope. Nuclei (including parasite) will be stained blue and cytoplasm and tissue structure will be stained pink.

**Step 4 – Immunostaining:** A slide will be deparaffinised and brought to water and antigen retrieval performed.

Then either a) or b)

a) An RNA probe specific for *Leishmania* and nuclear marker will be used. Slides will be examined by fluorescent microscopy.
b) Non-specific binding will be blocked using serum, an antibody for oligopeptidase B from *L.major* (OPB) applied followed by a fluorescently tagged secondary antibody and nuclei stain Yoyo-1.

Slides will be examined under a fluorescent microscope. Nuclei (including parasite) will be visible in one channel and parasite will be visible in the other channel.

**Culture**

**Step 1 – Limiting Dilution Preparation:** 100µl Schneider's insect media with 20% FCS will be put into 24 wells of a 96 well plate. 100µl Schneider's insect media with 20% FCS will be used to collect the biopsy scraping from the glass slide. Serial dilution will then be performed across the 24 wells. Plate will then be incubated at 26°C.

**Step 2 – Limiting Dilution Reading:** After 7 and 14 days the wells will be examined on an inverted microscope for parasite growth. The highest dilution with parasite growth will be noted at each time point.

**qPCR**

**Step 1 – DNA Extraction:** Biopsy portion will be weighed and extraction of total DNA will be performed using DNeasy blood and tissue kit (Qiagen) following the manufactures instructions. Tissue spiked with known parasite concentrations will used as a calibration curve.

**Step 2 – Parasite quantification:** SYBR green detection method will be used with primers targeting a leishmania kinetoplast minicircle DNA sequence. Results will be reported as parasites per mg tissue.

*(Further techniques as per our study objectives, not related to diagnosis, may require use of alternate media).

**Appendix 4. Punch biopsy SOP**

A punch biopsy of the skin is a very simple procedure performed in the outpatient setting which takes 10 to 15 minutes.

**Step 1- Prepare the Area to be Biopsied**:

Initially perform the study visit-specific history, examination, and image documentation. Then explain the biopsy procedures and obtain informed consent to the biopsy (suspected CL lesion), or biopsies (suspected CL lesion and matched normal skin on the contralateral arm). The informed consent also includes the subsequent study follow up. Next, the areas of skin where the biopsies are to be performed are cleaned with an alcohol swab to ensure sterile conditions.

**Step 2 - Anesthetise the Skin:**

The skin that is going to be removed is anesthetised by injecting a solution of 1% lignocaine and adrenaline just under the epidermis (sub-epidermally) using a 2ml syringe and orange needle. The injection continues until a “bleb” or bubble has formed under the skin greater than 4mm in diameter. The injection will burn slightly (much like a bee sting) due to a pH difference between the skin and the solution. The slight burning will quickly subside and the site will become numb.

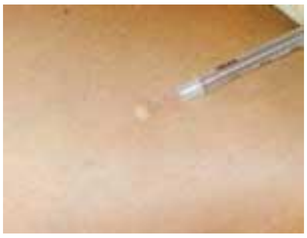

**Step 3 - Check for Numbness:**

After the local anaesthetic has been administered the areas to be biopsied should be checked to ensure that the skin is properly anesthetised. A needle is used to establish whether the skin has gone numb. Great care should be taken not to force the needle into the skin. The test site should be somewhere around the periphery of the area injected. Both of these precautions ensure a viable biopsy for diagnosis later. If the patient confirms no sensation, the biopsy can proceed. A pressure sensation is normal and expected but there should be no pain. If the area requires more anaesthetic a further injection (with a new syringe) is made until the skin is completely anaesthetised.

**Step 4 - Biopsy the Skin:**

Once the skin is anaesthetised, using a sterile disposable 4mm skin punch perpendicular to the skin, the clinician stretches the skin using the index finger and thumb providing traction is separate directions. Using the disposable punch the clinician then applies pressure and with a firm twisting action until the blade of the skin punch has pierced the epidermis of the skin. It is normal for the patient to experience some pressure sensation but no pain. By stretching the skin the defect is oval rather than round which results in a better cosmetic result when the area is sutured.

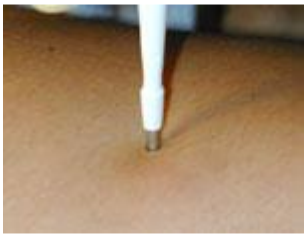

**Step 5 - Remove the Skin Punch:** After the blade has sufficiently “cored” or carved out a 4mm cylinder of skin the skin punch is removed. It is normal for the area to bleed after the punch is removed. Excess blood is wiped off with sterile gauze to expose the biopsy site.

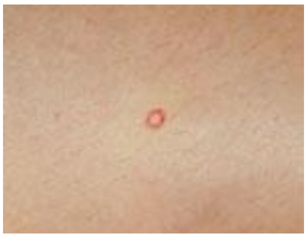

**Step 6 - Excise the Biopsy:** When the skin has been cored and cleared of excess blood, the next step is to remove the biopsy from the rest of the skin. Great care should be taken not to damage the epidermis by crushing it with forceps or by cutting it with a scalpel unnecessarily. The physician uses fine forceps or a skin hook to seize the dermis of the cored skin, pulls up the core to reveal dermis and subdermal fat, and uses a scalpel or scissors to snip the base of the lesion to cut the cored skin free. The biopsy is immediately placed in the vial containing Schneider’s insect media, as below in Appendix 3.

**Step 7 - Bandage Biopsy Site:** Once the full thickness punch biopsy has been removed from the skin there will usually be some degree of bleeding which should be absorbed with sterile gauze. Haemostasis and wound closure can be achieved with 2 simple interrupted sutures using 5-0 Ethilon. The biopsy sites are then covered with small adhesive dressings.

**Step 8 – Biopsy Site Care**: The biopsy site should be kept clean. The site should not be submerged in water (i.e. no swimming, hot tubs, baths etc) for 48 hours. In the long term, minimal scarring may occur. In a few cases the biopsy site may form a protrusion or bump but will heal normally

**Step 9 – Suture Removal**

If the sutures are still *in situ* at the 10 day follow up they are removed using gentle traction on one the ‘ears’ and carefully cutting the ‘intra-dermal loop’ on the skin side of the knot, and then gently pulling the whole suture clear.

**Appendix 5: GAD-7 and DLQI questionnaires**

GAD-7

| Over the last 2 weeks, how often have you been bothered by the following problems? | Not at all | Several days | More than half the days | Nearly every day |
|---|---|---|---|---|
| 1. Feeling nervous, anxious or on edge | 0 | 1 | 2 | 3 |
| 2. Not being able to stop or control worrying | 0 | 1 | 2 | 3 |
| 3. Worrying too much about different things | 0 | 1 | 2 | 3 |
| 4. Trouble relaxing | 0 | 1 | 2 | 3 |
| 5. Being so restless that it is hard to sit still | 0 | 1 | 2 | 3 |
| 6. Becoming easily annoyed or irritable | 0 | 1 | 2 | 3 |
| 7. Feeling afraid as if something awful might happen | 0 | 1 | 2 | 3 |

Total Score ____ = Add Columns ____ + ____ + ____

If you checked off any problems, how difficult have these problems made it for you to do your work, take care of things at home, or get along with other people?

| Not difficult at all | Somewhat difficult | Very difficult | Extremely difficult |
|---|---|---|---|
| ☐ | ☐ | ☐ | ☐ |

#### Dermatology Life Quality Index

**The aim of this questionnaire is to measure how much your skin problem has affected your life OVER THE LAST WEEK. Please tick (✓) one box for each question.**

| | | | | | |
|---|---|---|---|---|---|
| 1. | Over the last week, how **itchy**, **sore**, **painful** or **stinging** has your skin been? | Very much | ❒ | | |
| | | A lot | ❒ | | |
| | | A little | ❒ | | |
| | | Not at all | ❒ | | |
| 2. | Over the last week, how **embarrassed** or **self conscious** have you been because of your skin? | Very much | ❒ | | |
| | | A lot | ❒ | | |
| | | A little | ❒ | | |
| | | Not at all | ❒ | | |
| 3. | Over the last week, how much has your skin interfered with you going **shopping** or looking after your **home** or **garden**? | Very much | ❒ | | |
| | | A lot | ❒ | | |
| | | A little | ❒ | | |
| | | Not at all | ❒ | Not relevant | ❒ |
| 4. | Over the last week, how much has your skin influenced the **clothes** you wear? | Very much | ❒ | | |
| | | A lot | ❒ | | |
| | | A little | ❒ | | |
| | | Not at all | ❒ | Not relevant | ❒ |
| 5. | Over the last week, how much has your skin affected any **social** or **leisure** activities? | Very much | ❒ | | |
| | | A lot | ❒ | | |
| | | A little | ❒ | | |
| | | Not at all | ❒ | Not relevant | ❒ |
| 6. | Over the last week, how much has your skin made it difficult for you to do any **sport**? | Very much | ❒ | | |
| | | A lot | ❒ | | |
| | | A little | ❒ | | |
| | | Not at all | ❒ | Not relevant | ❒ |
| 7. | Over the last week, has your skin prevented you from **working** or **studying**? | Yes | ❒ | | |
| | | No | ❒ | Not relevant | ❒ |
| | If "No", over the last week how much has your skin been a problem at **work** or **studying**? | A lot | ❒ | | |
| | | A little | ❒ | | |
| | | Not at all | ❒ | | |
| 8. | Over the last week, how much has your skin created problems with your **partner** or any of your **close friends** or **relatives**? | Very much | ❒ | | |
| | | A lot | ❒ | | |
| | | A little | ❒ | | |
| | | Not at all | ❒ | Not relevant | ❒ |
| 9. | Over the last week, how much has your skin caused any **sexual difficulties**? | Very much | ❒ | | |
| | | A lot | ❒ | | |
| | | A little | ❒ | | |
| | | Not at all | ❒ | Not relevant | ❒ |
| 10. | Over the last week, how much of a problem has the **treatment** for your skin been, for example by making your home messy, or by taking up time? | Very much | ❒ | | |
| | | A lot | ❒ | | |
| | | A little | ❒ | | |
| | | Not at all | ❒ | Not relevant | ❒ |

#### Appendix 6: Source Data Definition

| Type of Data | Source Document |
|---|---|
| Informed consent | Paper copy in SSF |
| Relevant Medical History and Current Medical Conditions | CRF |
| Physical Examination and Observations | CRF |
| Concurrent medication | CRF |
| Clinical Study History | CRF |
| Diary Card | CRF |
| Fulfilment of eligibility criteria | CRF |
| Demographics | CRF |
| Clinical Laboratory Reports – Haematology, Biochemistry, HIV, | Printed report forms, kept in CRF |
| Time of study sand fly bite exposure in the clinical unit | CRF |
| Date/time of blood sampling | CRF |
| Date of visits/examinations | CRF |
| Adverse events | CRF |
| Protocol Deviations | CRF and file notes in SSF |
| Withdrawal | CRF |

### LEISH_Challenge:

#### A *Leishmania major* human challenge study

##### Participant Information Leaflet

For more information contact the project team at York:

##### Background

The University of York would like to invite you to take part in a research study, "A clinical study to develop a controlled human infection model using *Leishmania major*-infected sand flies".

Our research relates to an infection called leishmaniasis which mainly occurs in tropical countries. It affects millions of people and causes around 20,000 deaths across the world every year. There are different types of leishmaniasis around the world and some can be very serious. They affect the skin (cutaneous leishmaniasis) or the internal organs of the body (visceral leishmaniasis). Some of the milder forms will produce skin problems which will be localised, whilst other forms of leishmaniasis will cause widespread skin changes. The skin lesions (an abnormal change in the skin) of cutaneous leishmaniasis can be disfiguring if left untreated.

Our group is designing a type of research study, known as a controlled human infection model (CHIM), whereby human participants are deliberately exposed to infections. This approach of exposing humans to infectious diseases has been used for hundreds of years as a method for better understanding how diseases progress and for the evaluation of vaccines and treatments. Vaccine development for neglected tropical diseases has much to gain by the use of CHIMs, and this approach has already been used in developing vaccines for cholera, malaria, influenza and dengue fever. No vaccines are currently approved for leishmaniasis, a disease affecting millions each year. However, new candidate vaccines have emerged over the last few years, emphasising the need for a CHIM for leishmaniasis.

##### What is the purpose of the study?

Our objective is the development of a controlled human infection model of *Leishmania major* using sand fly transmission which is effective and safe. Leishmaniasis is caused by the Leishmania parasite and is transmitted by sand flies. The parasite is tiny and not visible to the naked eye, whereas the sand fly is visible but small and inconspicuous.

It has been shown that sand fly saliva is crucial for optimal infection. In addition, studies using mice injected with the *Leishmania* parasite have shown that using a needle to inoculate the parasite, rather than using sand flies, may be unreliable when testing vaccines. For these reasons, this study will be using sand flies to infect human participants in a controlled human infection model with Leishmaniasis.

In late 2019, we conducted an initial successful study using uninfected (disease-free) sand flies. Information from the original study will inform the way we work this time using **infected** sand flies.

The aim is to develop a model that we can use in the future to assess vaccines against Leishmaniasis. We will **not** be testing a vaccine in this study.

We will be using a species known as *Leishmania major* which does not cause severe disease but causes a localised lesion in the skin at the site of a sand fly bite. This can appear similar in appearance to a spot. This lesion can be treated with several known treatments for leishmaniasis, and in this study will be using a small biopsy to remove the lesion. We believe that usually no further treatment will be necessary, but if it is we may use a freezing treatment (cryotherapy). We will also ask your permission for an optional small biopsy of healthy skin from the opposite arm. This will help us to compare how the skin reacts to the *Leishmania* parasite.

Bear in mind that in addition to the clinical interventions and observations, we will also ask you to take part in a focus group with other participants which will be audio recorded with your consent.

Before agreeing to take part in this research, please read this information sheet carefully and let us know if anything is unclear or you would like further information.

#### Do I need to do anything beforehand?

Please take any regular medications ensuring that the study team are aware of these. If you have been started on any new medications it is extremely important to tell the study team as soon as possible before you attend. You may eat and drink as normal but a light diet is suggested.

Please wear loose clothes or a T-shirt for example, to ensure that the area of skin on your arms are easy to access and exposed. It is advised to bring a book or some reading material in case you have to wait for any period of time.

#### What happens if there is a problem?

If you feel at any stage that there has been a problem with your participation, you can discuss this matter, in confidence, with Professor Charles Lacey, the Chief Investigator, on: tel 01904 328879 or

#### Why have I been asked to take part?

We are advertising for volunteers to take part in this study. Volunteers must be men and women aged between 18 and 50 years old. Enrolled female participants must **not** be pregnant or breastfeeding and must be using an effective form of contraception. Volunteers must be healthy and **not** be at risk of serious infections and must **not** have any chronic skin conditions. Volunteers should **not** have travelled in the last 30 days to any area where leishmaniasis is present or for 30 consecutive days at any time in the past. There should be **no** history of significant reaction to insect bites. A full review of any possible reasons that could prevent you from taking part will be performed by the medical team if you express an interest in the study.

#### Where do the sand flies come from?

We are working with scientists in the Czech Republic who are insect experts (entomologists). They have been rearing sand flies for several years and are experts in developing sand flies for use in research. We have been working closely with these scientists who will provide us with the right kind of sand fly for our study. There will be up to 2 different types of sand flies used, both of which can transmit *Leishmania.*

#### Where does the Leishmania parasite come from?

Although there are several laboratories and parasite banks around the world that have parasites available for use in experiments, we wanted to know the precise origin of the parasites that we use to be able to prove their safety and effectiveness. Therefore, we have obtained fresh parasites from healthy individuals in Israel who developed a *Leishmania major* lesion which was not serious, healed well and from which they recovered fully. This parasite has then been tested by laboratories in Israel, Czech Republic, Germany and York to check some of the safety aspects, and to make sure it will be effective in our proposed model

#### What will happen if I agree to take part?

This study is designed to test the safety and effectiveness of the *Leishmania* human challenge protocol in healthy volunteers.

The number of study visits will depend on when a lesion develops as detailed in the diagram below. All visits will take place at Translational Research Facility, Q Block, Department of Biology, University of York, York, YO10 5DD.

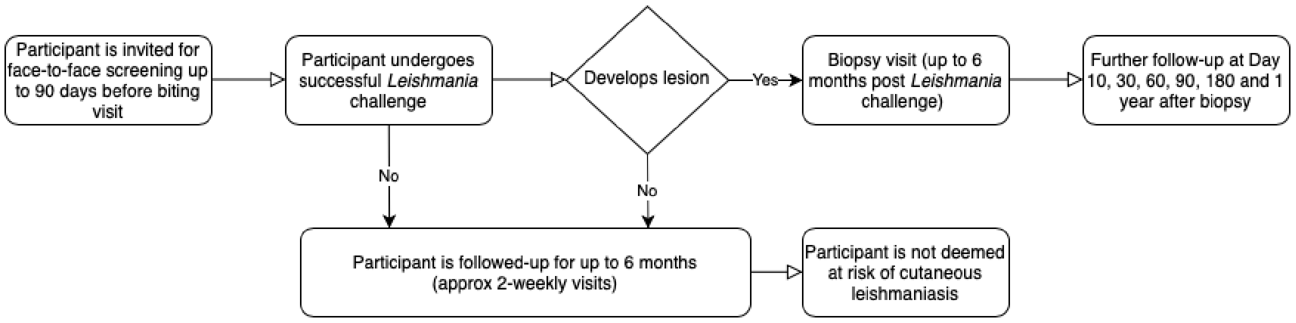


Participant wearing sand fly biting chamber. There are sand flies contained within the 'watch-like' chamber actively biting

Image available on request as per medRxiv requirements

- There are 3 phases, as shown above
    - Phase 1 - The screening visit for the study
    - Phase 2 - The leishmania challenge, and then the follow up until a leishmania lesion (spot/lump) develops (or otherwise, if no leishmania lesion develops)
    - Phase 3 - Then if the leishmania lesion (spot/lump) appears, the treatment and follow up phase
- In Phase 1, we will first ask you some screening questions by email and/or telephone contact, before inviting you for the formal screening visit:
    - **Study visit 1**: Screening including blood tests will be performed to assess suitability. This can take place up to 3 months before the main study. This will last an hour.
- Phase 2:
    - **Study visit 2**: The 2nd visit will be the leishmania challenge itself. Using a wrist watch-like biting chamber, 5 infected sand flies will be placed against your skin for 30 minutes. Photography and videography of the bite site will be performed with your permission. You will be observed for a period of time before being allowed to go home. A doctor and a nurse, or two doctors, and an insect specialist (entomologist) and other senior study investigators will be present throughout. This session will last around 3 hours. You will be asked if you agree to provide an optional video-recorded account of your experience.
    - **Study visits 3-5:** Follow-up visits will take place at approximately 4 days, 14 days and 28 days after the biting visit. These will be 30-minute visits for examination of the bite site and blood tests on days 14 & 28. It is expected that a cutaneous leishmaniasis lesion will develop at some point from 28 days onwards.
- After visit 3, participants will have ongoing follow-up until they develop a cutaneous leishmaniasis lesion. **Study visits 4-11**; these will take place at the following timepoints following sand fly biting: 2 weeks, 4 weeks, 6 weeks, 8 weeks, 10 weeks, 14 weeks, 18 weeks and 22 weeks. Study visits 6, 8, and 10 are planned to be 'virtual visits' by video calling for examination of the bite site. Study visits 5, 7, 9, and 11 will be 30-minute 'actual visits' for blood tests and examination of the bite site.
- If by the time of **visit 11** (22 weeks) no cutaneous leishmaniasis lesion has developed, then we will presume that no leishmania has been transmitted. You will be given a leaflet with details of the study team in case any issues or problems develop later. Your GP will also be informed at this stage, and no more study visits will be scheduled.
- Once a cutaneous leishmaniasis lesion is present at any study visit that is greater than or equal to 3mm in diameter, you will be followed-up in a **Biopsy visit**. This will involve a small biopsy to remove the lesion (see description in 'What are the potential risks of taking part?'). The biopsy visit will reset your schedule of follow-up visits as follows:-
- Following the biopsy visit, a further visit will take place at 10 days to check healing of the biopsy site.
- Further follow-up will then take place at 30, 60, 90, 180 days and the last study visit at 1 year following biopsy treatment. It is anticipated that some of those visits can be 'virtual' using video calling.
- We would also like volunteers to take part in a focus group. This will take place after the last study visit and it is anticipated that this will last between 2-3 hours and involve the participants that have taken part in the study. The focus group will be led by Prof Georgina Jones, a health psychologist, and provide the opportunity for you to share your experiences of participating in the study and any concerns you had, and help us to shape the design of the next stage of the research. The Focus Group will take place at the University of York, and

non-identifiable information will be recorded and will contribute to future publication and research.

Blood will be taken at Study visits 1, 4, 5, 7, 9, & 11. Blood will also be taken on the day of the biopsy, and also on days 30, 90 and 360 (last study visit) following biopsy. Approximately 50 ml of blood will be taken each time. There is a small risk of bruising following the blood tests. We will ask your permission to contact your GP to confirm your eligibility prior to starting the study.

We also want to try and measure if the research study causes you any stress or upset, and whether the bite site or cutaneous leishmania lesion causes you any or much discomfort or difficulties. To do this we will ask you to complete two brief questionnaires, known as GAD-7 & DLQI, throughout the study. We estimate these take 5-10 minutes total to complete and will be performed at Study Visits 1, 4, 5, 7, 9, & 11 Following the biopsy, these questionnaires will be recorded at every further visit.

#### What is a biopsy?

A biopsy of the skin is a very simple procedure performed in the outpatient setting which takes 10 to 15 minutes. A small sample of skin is removed and then examined by the study team. There are two possible ways in which this can be performed either an ‘excision’ biopsy in which a lesion is fully removed or an ‘incision’ in which a small sample of a skin is removed. The latter is often done with an instrument called a ‘punch’ device.

In both procedures the skin is cleaned to reduce risk of infection and then the skin is ‘anesthetised’ (numbing agent used to prevent discomfort during the procedure). A local anaesthetic is injected just underneath the skin to numb the area. This can cause a stinging sensation but will quickly subside and the area will become numb.

In an ‘excision’ biopsy, the area to be removed is drawn out on to the skin. In this study this is likely to be up to 2cm by up to 1cm in size (approximately the size of a jelly bean sweet). The skin is then removed with a small knife called a scalpel. 3-4 small stitches will then be used to bring the sides of the excision biopsy together to help with healing and a dressing put over this.

In a ‘punch’ biopsy a small pencil like device which looks like a miniature apple corer is used to remove a small sample of the skin. This is a much smaller version of a hole-punch device. 2-3 small stitches will then be used to bring the sides of the skin together to help with healing a dressing put over this.

Your doctor will discuss which of the above techniques will be used. You will be awake throughout and will be able to talk to the doctor throughout the procedure.

#### What is the purpose of the biopsy?

The excision biopsy will be used a treatment for cutaneous leishmaniasis, to stop the lesion growing further. This method has been used in other centres for this purpose. We will also be analysing this skin sample in the laboratory to look for evidence of Leishmania parasite and to perform other tests which will help in our study and future studies against leishmaniasis.

If you have agreed we may also perform a biopsy of healthy skin on the opposite arm to the *Leishmania* lesion. This is so we can compare what happens to an individual’s healthy skin versus infected skin following a sand fly bite. We will use the punch biopsy technique to do this to ensure that we take the smallest possible area of skin.

#### Who is organising and funding this study?

Funding has been provided by the UK Medical Research Council (Ref: MR/R014973/1)

#### What are the potential risks of taking part?

Volunteers may experience a 'pin-prick' sensation when bitten by sand flies. Common effects following sand fly bites (as with all insect bites) include some swelling, pain and redness at the bite site. This may take up to 48 hours to develop. Just as with any insect bite there is the potential for a skin infection which may require antibiotic treatment.

The species of sand fly that we are using has not been associated with any serious reaction in the past. Although hundreds of thousands of people are bitten by sand flies every day without any serious effects, we want to absolutely ensure your safety. For this reason a health check and blood tests taken prior to the study will help to identify volunteers who are more likely to suffer adverse effects and these individuals will be excluded from participating further (this includes a blood test to look specifically at sand fly bite reaction as well as other reactions).

To date, there have been no reported cases of anaphylactic (life-threatening) reactions to sand fly bites in the whole medical literature. However, there might still be a small theoretical risk of serious reaction to sand fly bite, just as with any insect bite. When a serious reaction occurs, although rare it usually presents with breathing problems and widespread skin rash. Within the very small number of people who develop a serious reaction, a small number of these may be at risk of death. We have appropriate treatment and a medical team on-site at all times in the very unlikely event that a serious reaction occurs.

We can tell quite accurately if sand fly biting has occurred, and in our previous study we were 100% successful. In the unlikely event that a participant does not develop any kind of bite or spot in the first four weeks, it is very likely that the sand flies did not transmit any parasites to the participant. If this occurs, the participant will only have regular follow-up till 28 days and then a final check at around 6 months to ensure that all is well.

We expect participants will develop a cutaneous leishmaniasis lesion at the sand fly biting site. This lesion can appear similar to a spot and we expect it to develop during the first 5 months after biting. There are no recorded cases of significant ill-effect from this in healthy individuals. The lesion will be allowed to grow up to 3mm in diameter (about one eighth of an inch). The sand flies are reared free from harmful infections. The sand flies will be infected with the *Leishmania major* parasite at the University of York. If left to grow without treatment, the lesion can develop into a sore known as an 'ulcer', although a number of individuals with this lesion have gone on to 'self-heal' without any treatment.

It is possible that after the biopsy there may still be some parasites left in the body near the site of infection. Normally these help to keep the immune system stimulated and provide protection against reinfection. This might be beneficial if the participant travels to a leishmaniasis endemic region and is exposed naturally. However, the presence of this small number of parasites makes it possible that immunosuppression (e.g. through an organ transplant, HIV or drugs that affect the immune system) could cause reactivation of the lesion. Although this can happen in other forms of leishmaniasis, we are unable to find any descriptions of this occurring with *Leishmania major* after successful treatment.

Just like with any medical procedure there are some risks involved with the biopsy. These are rare but are important to know about and include:
Pain
Infection
Bleeding
Incomplete excision of the lesion. This might mean a repeat biopsy is needed.
Bruising
Numbness at the scar site
Small scar (similar to a chicken pox scar)
Very rarely:-
A small bump might form at the scar site but if this occurs, it is likely to settle over time.
Wound breakdown and ulceration

As treatment, we intend to perform an 'excision biopsy' once a lesion develops. This will occur when it gets to about 3mm in diameter. This is a simple outpatient procedure which takes about 15 minutes. This procedure is performed in a sterile fashion, using local anaesthetic. The local anaesthetic can feel like a sharp scratch which will last a few seconds. Once the area is numb and no pain is felt, a small ellipse centred on the spot, approximately 18 mm x 7 mm (¾ x ¼ inch), will be removed using a scalpel. The long axis is oriented along the natural lines of the skin to get the best cosmetic result. A very small amount of blood may be visible immediately following the procedure. 2-3 small stitches are used at the biopsy site. For 2 days, contact with water just at this site should be avoided.

Some individuals may develop a very small scar at the site of the biopsy although this is likely to be minimal. Some individuals may notice a small bump at the site of the lesion. The stiches will be removed after 10 days.

If you feel unwell at any point during or after the study you will be supported by the research team and given any necessary medical attention. This will include a support line for advice if needed and admission to hospital, although this is thought to be very unlikely.

#### What precautions will you take to reduce the risk of COVID-19?

The study team involves doctors who are used to dealing with COVID-19 and wearing PPE (personal protective equipment). The team will adhere to all current UK Government guidelines. In addition all research will be approved by NHS and University research committees with a risk assessment carried out to mitigate any risks.

The study team will adhere to up-to-date clinical practice, and use of any necessary PPE. A dedicated entrance is used to enter the facility and there will be no contact with non-study team members. The study team is small and will likely comprise of 3 members all wearing PPE if deemed necessary by NHS practices. Good hand-hygiene practices will be employed throughout as is common practice in our clinical facility, and our facilities are cleaned regularly. Participants will be asked to wear a mask where necessary.

If you develop a fever, you must self-isolate in accordance with current government guidelines and it is feasible that some study visits can take place via video calling/phone. It must be noted that you may develop a fever following sand fly biting, although the risk of this is deemed to be very low and was not observed in our initial study.

Participants will be asked to undertake regular testing for SARS-CoV-2, if such facilities exist and are easily accessible (e.g. lateral flow testing), although this will not be mandatory.

#### Compensation for your involvement

Participants will be compensated for their time and for the inconvenience at approximately these rates:
Screening visit: £20
*Leishmania* challenge: visit £500
Follow-up visits: £40 per visit
Per completed participant-submitted photograph & diary card event: £5 (this will take place up to a maximum of once to twice weekly only in accordance with study requirements)
Biopsy visit: £500
Final visit: £100
Focus Group visit: £100
(Total up to approximately £2500 depending on involvement.)

#### What are the potential benefits of taking part?

The information from this study will help towards further understanding of sand fly bites and transmission of *Leishmania* parasite. This will contribute to developing a model for testing vaccines using infected sand fly bites. Ultimately this research will help in the effort to beat leishmaniasis.

You will also undergo health and wellbeing screening by the medical doctors who are involved in the study. This will involve a history, physical examination and blood tests. If there are any abnormal or unexpected findings from these screenings, this will be discussed with you by the study team and you will be referred to an appropriate medical speciality or your GP within the NHS.

Exposure to sand fly resulting in a cutaneous leishmaniasis lesion, may for some individuals protect against future leishmaniasis infection after further exposure e.g. if a participant travels to an area where leishmaniasis is present. This is a potential benefit but study investigators will be unable to quantify this.

#### Do I have to take part in the study?

No, participation is optional. If you do decide to take part, you will be given a copy of this information sheet for your records and will be asked to complete a participant consent form. If you change your mind at any point during the study (including during the focus group), you will be able to withdraw your participation without having to provide a reason. However given the nature of the study, and the need for ongoing follow-up, this will be discouraged after the leishmania challenge visit has taken place.

Any screening data taken prior to commencing the study will be destroyed if the participant is deemed not to be eligible to enter. Otherwise any samples and information obtained up until the point of withdrawal will be retained.

#### What will happen to my samples?

The blood samples will be tested at the Translational Research Facility, HYMS / Department of Biology, University of York and York Hospitals NHS Foundation Trust. This is where they will be studied by the research team. Samples will be tested in secure laboratory facilities accessible only by authorised research staff. Samples will be labelled with a unique ID number and the date only but no personal details, so they cannot be identifiable as having been donated by you. The samples will only be used in research projects that have been independently reviewed and approved by the

Department of Biology Ethics Committee. The samples will be stored for 12 months and may be used for additional future research with your permission. After this period, the samples will be destroyed.

#### What happens if there is a problem?

If you feel at any stage that there has been a problem with your participation, you can discuss this matter, in confidence, with Professor Charles Lacey, the Chief Investigator, on: tel 01904 328879 or

The University of York holds Public Liability ("negligent harm") and Clinical Trial ("non-negligent harm") insurance policies which apply to this study. If you can demonstrate that you experienced serious and enduring harm as a result of your participation in this study, you may be eligible to claim compensation without having to prove that University of York is at fault. If the injury was unrelated to the study, University of York will not be required to compensate you in this way. Your legal rights to claim compensation for injury where you can prove negligence are not affected. Please contact the Chief Investigator if you would like further information about the insurance arrangements which apply to the trial.

#### On what basis will you process my data (i.e. personal information)?

Under the General Data Protection Regulation (GDPR), the University has to identify a legal basis for processing personal data and, where appropriate, an additional condition for processing special category data. (Please see the additional document on data protection and personal information)

#### What happens next?

If you are interested in this study or if you have any questions, please contact one of the study team members (details given on the front of this leaflet). One of our study team will talk to you about what is involved and ask questions about your suitability for this study. The next step will then be to invite you to a screening assessment at our facilities at the University of York.

*This project: "Development of a human challenge model of Leishmania major infection as a tool for assessing vaccines against leishmaniasis" is funded by the Medical Research Council and Department for International Development (ref: MR/R014973/1)*

MRC Medical Research Council

Department for International Development

### LEISH_Challenge:

### A *Leishmania major* human challenge study

#### Participant Data Protection and Personal Data

##### On what basis will you process my data (i.e. personal information)?

Under the General Data Protection Regulation (GDPR), the University has to identify a legal basis for processing personal data and, where appropriate, an additional condition for processing special category data.

In line with our charter which states that we advance learning and knowledge by teaching and research, the University processes personal data for research purposes under Article 6 (1) (e) of the GDPR:

*Processing is necessary for the performance of a task carried out in the public interest*

Special category data is processed under Article 9 (2) (j):

*Processing is necessary for archiving purposes in the public interest, or scientific and historical research purposes or statistical purposes*

Research will only be undertaken where ethical approval has been obtained, where there is a clear public interest and where appropriate safeguards have been put in place to protect data.

In line with ethical expectations and in order to comply with common law duty of confidentiality, we will seek your consent to participate where appropriate. This consent will not, however, be our legal basis for processing your data under the GDPR.

##### How will you use my data?

Data will be processed for the purposes outlined in this information sheet.

##### Will you share my data with 3$^{rd}$ parties?

No. Data will be accessible to the project team only.

Anonymised data may be reused by the research team or other third parties for secondary research purposes.

##### How will you keep my data secure?

The University will put in place appropriate technical and organisational measures to protect your personal data and/or special category data. For the purposes of this project we will store data on dedicated servers and / or on the University of York central data store, which provided secure long term storage for data, including daily backups, according to the University of York Research Data Management Policy.

A study master file will be created to include the study protocol, original signed consent forms and ethics/governance documentation. It will also include a list of study group participants, their email address and their telephone number. The study master file will be stored at Department of Biology in a secure, fire and rodent-proof cabinet that is only accessible to authorised members of staff.

Information will be treated confidentiality and shared on a need-to-know basis only. The University is committed to the principle of data protection by design and default and will collect the minimum amount of data necessary for the project. In addition, we will anonymise or pseudonymise data wherever possible.

#### Will you transfer my data internationally?

No. Data will be held within the European Economic Area in full compliance with data protection legislation.

#### Will I be identified in any research outputs?

No.

#### How long will you keep my data?

Data will be retained in line with legal requirements or where there is a business need. Retention timeframes will be determined in line with the University's Records Retention Schedule.

#### What rights do I have in relation to my data?

Under the GDPR, you have a general right of access to your data, a right to rectification, erasure, restriction, objection or portability. You also have a right to withdrawal. Please note, not all rights apply where data is processed purely for research purposes. For further information see, https://www.york.ac.uk/records-management/generaldataprotectionregulation/individualsrights/

#### Questions or concerns

If you have any questions about this participant information sheet or concerns about how your data is being processed, please contact the study team in the first instance. If you are still dissatisfied, please contact the University's Acting Data Protection Officer at.

#### Right to complain

If you are unhappy with the way in which the University has handled your personal data, you have a right to complain to the Information Commissioner's Office. For information on reporting a concern to the Information Commissioner's Office, see www.ico.org.uk/concerns.

*This project: "Development of a human challenge model of Leishmania major infection as a tool for assessing vaccines against leishmaniasis" is funded by the Medical Research Council and Department for International Development (ref: MR/R014973/1)*

MRC Medical Research Council

Department for International Development
